# COVID-19, Race, and Redlining

**DOI:** 10.1101/2020.07.11.20148486

**Authors:** Graziella Bertocchi, Arcangelo Dimico

## Abstract

Discussion on the disproportionate impact of COVID-19 on African Americans has been at center stage since the outbreak of the epidemic in the United States. To present day, however, lack of race-disaggregated individual data has prevented a rigorous assessment of the extent of this phenomenon and the reasons why blacks may be particularly vulnerable to the disease. Using individual and georeferenced death data collected daily by the Cook County Medical Examiner, we provide first evidence that race does affect COVID-19 outcomes. The data confirm that in Cook County blacks are overrepresented in terms of COVID-19 related deaths since—as of June 16, 2020—they constitute 35 percent of the dead, so that they are dying at a rate 1.3 times higher than their population share.

Furthermore, by combining the spatial distribution of mortality with the 1930s redlining maps for the Chicago area, we obtain a block group level panel dataset of weekly deaths over the period January 1, 2020-June 16, 2020, over which we establish that, after the outbreak of the epidemic, historically lower-graded neighborhoods display a sharper increase in mortality, driven by blacks, while no pre-treatment differences are detected. Thus, we uncover a persistence influence of the racial segregation induced by the discriminatory lending practices of the 1930s, by way of a diminished resilience of the black population to the shock represented by the COVID-19 outbreak. A heterogeneity analysis reveals that the main channels of transmission are socioeconomic status and household composition, whose influence is magnified in combination with a higher black share.

**JEL Codes:** I14, J15, N32, N92, R38.

## 1 Introduction

Ever since when The Atlantic magazine (Kendi, 2020), on March 4, 2020, first launched a cry for attention to the disproportionate impact of COVID-19 on African Americans, the issue has taken center stage in the debate on the socioeconomic implications of the pandemic. Nevertheless, a race-disaggregated analysis of individual data has so far been lacking due to data unavailability.

The urgency of the racial issue and the need for data collection to account for ethnic and racial factors has been widely recognized also within the medical and epidemiological literature. Preliminary evaluations suggest that the high risk of COVID-19 death for minority ethnic groups can be explained by pre-existing health conditions, such as diabetes, obesity, hypertension, and asthma, that are more common among these groups, possibly because of genetic and biological factors. However, the emerging consensus is that the race differential in the prevalence of COVID-19 is also associated with socioeconomic correlates. As argued by Yancy (2020), a large share of the black population in the US lives in poor areas characterized by high unemployment, low housing quality, and unhealthy living conditions, making low socioeconomic status a critical risk factor. Furthermore, the higher prevalence of comorbidities in blacks is also itself associated with highly persistent socioeconomic factors. Other relevant health-related and behavioral risk factors, such as smoking, drinking, and drug abuse, are deeply ingrained in cultural norms that are also driven by social inequalities. African Americans suffer for further disadvantage in their ability to adhere to social distancing norms, as working from home, avoiding public transportation, and finding refuge in second homes away from crowded cities, are indeed privileges that are denied to the majority of them. Anecdotal evidence also suggests the possibility of a different response by health practitioners to black individuals showing COVID-19 symptoms. All the above considerations, as summarized by Yancy (2020) resonate with the long-standing debate on the racial disparities that are deeply entrenched in US history and point to a need to account for race in COVID-19 research and to investigate the role not only of biological factors but also of socioeconomic ones, while acknowledging that the latter may be rooted on inequalities that have been long neglected.

In this paper, we take advantage of an unexplored and extraordinarily detailed source of information on daily deaths from COVID-19 that includes race among a wide array of individual characteristics. The data are collected by the Medical Examiner’s Officer and made available by the Government of Cook County, Illinois, the county that hosts—among others—the City of Chicago.^1^

On the basis of the Cook County data we can provide, to start with, first evidence of how race affects COVID-19 individual outcomes. Furthermore, we can illustrate the correlates and dig into the historical determinants of such outcomes. The higher COVID-19 death toll paid by black Americans has been linked to the “redlining” policies introduced by the Home Owners Loan Corporation (HOLC) in the 1930s.^2^ These policies are believed to have favored the development of segregated neighborhoods plagued by unemployment, low housing quality, and unhealthy living conditions. In a public speech following the death of George Floyd, in the midst of the pandemic, former President Obama has also attributed the explosion of protests to the history of racial discrimination, including redlining.^3^ By combining the information on COVID-19 fatalities in Cook County with the redlining maps for the Chicago area, we can assess the explanatory power of the induced vulnerabilities that are rooted in history.

In more detail, the first contribution of the paper is to document how race affects COVID-19 mortality in Cook County, on the basis of individual data that also account for age, gender, and pre-existing health conditions. The analysis confirms that blacks are overrepresented in terms of COVID-19 related deaths since—as of June 16, 2020—they constitute 35 percent of the deaths against a black population share of only 27 percent in our sample. In other words, blacks are dying at a rate 1.3 times higher than their population share.

Second, by exploiting the fact that the Cook County Medical Examiner also provides the geographical coordinates of the home address of each individual that died from COVID-19, we combine the spatial distribution of mortality with the redlining maps for the Chicago area, to assess whether the higher vulnerability of blacks to the disease can indeed be found in socioeconomic inequalities rooted in history, rather than in biological conditions. Using cross-sectional information about individual deaths from COVID-19, we show that the probability that an individual who died from COVID-19 is black is much larger in lower graded areas, that is, those that were historically redlined and yellowlined, even after controlling for demographic and socioeconomic factors as well as pre-existing conditions. However, where the black population share is also controlled for, the influence of the HOLC policies is fully absorbed by that of the former, suggesting that the latter did induce segregation along racial lines. A warning is in order since the cross-sectional analysis, being based solely on information about those that died from COVID-19, is severely biased because of sample selection. This limitation motivates our next set of results.

Third, to establish a causal link between the vulnerabilities induced by historical redlining and the racial gaps in COVID-19 outcomes, and to disentangle socioeconomic factors from other determinants which normally correlate with African Americans, we rely on an event study analysis based on a weekly balanced panel of deaths from any cause that we assemble at census block group level over the period from January 1, 2020 to June 16, 2020. In this way, we can test for pre-treatment differences while controlling for block group fixed effects which should filter out the effect of invariant socioeconomic factors. Consistent with the cross-sectional evidence, to determine the treatment we refer to block groups that belong to HOLC areas graded either C or D, that is, either yellow or redlined. We find that, while no pre-treatment differences are detected, mortality in treated neighborhoods increases sharply after the epidemic shock, and the increase is driven by the death toll of African Americans.

We also carry out a heterogeneity analysis focused on the degree of vulnerability to shocks, that we measure using data provided by the Centers for Disease Control and Prevention (CDC). The analysis reveals that the channels through which historical redlining influences COVID-19 outcomes are socioeconomic characteristics, in particular personal income and the population share below poverty, and household composition, in particular the population shares of the elderly and of single parent households. Strikingly, however, the influence of these factors manifests itself only in combination with a higher black share. We complete our investigation with a battery of robustness checks, extensions, and falsification tests.

Overall, the evidence we collect points to a persistence influence of the racial segregation introduced by the discriminatory lending practices of the 1930s, that operates by way of an asymmetric effect of the epidemic shock, which is in turn channeled through a diminished resilience of African Americans. Far from being determined by genetic and biological factors, their vulnerability can be linked to socioeconomic status and household composition, as channels through which the legacy of the past manifests itself.

The paper is organized as follows. Section 2 presents related literature. Section 3 summarizes background information on historical redlining policies and Cook County. Section 4 describes the data. Section 5 presents cross-sectional evidence. Section 6 introduces an event study approach and the corresponding baseline results, while Section 7 is devoted to robustness checks and Section 8 to the heterogeneity analysis. In Section 9 we extend the investigation to the Hispanic population. Section 10 presents two falsification tests respectively based on deaths in the years 2017 to 2019 and Spanish flu deaths in 1918. Section 11 concludes. The Appendix contains additional figures and tables.

## 2 Related literature

We contribute to several streams of the literature. The first is the literature on the racial impact of COVID-19. The so far largest epidemiological study on the racial impact of COVID-19 has been performed in the UK based on the medical records of more than 17 million individuals (OpenSAFELY Collaborative, 2020, working on behalf of NHS England).^4^ The study confirms that pre-existing medical conditions such as diabetes or deprivation are linked to a higher likelihood of in-hospital death, but also that clinical risk factors alone cannot explain the observed disparities. Again with a focus on the UK, Bhala et al. (2020) also support structural, socioeconomic and environmental explanations, rather than biological ones, of the racial differences in COVID-19 susceptibility. Attention is called to the fact that ethnic/racial minorities hold highly-exposed jobs in health and social care, retail, and public transport, and to cultural habits including the approach to worship and the multigenerational family structure. On the basis of medical data from COVID-19 patients at a hospital in Louisiana, Price-Haywood et al. (2020) find that blacks were overrepresented among patients and fatalities, but did not show higher inhospital mortality than whites after controlling for differences in sociodemographic (i.e., insurance type and zip code of residence) and clinical characteristics on admission.

Despite the above mentioned growing body of contributions within medicine and epidemiology, within the economics field the literature on the racial impact of COVID-19 is still limited. Borjas (2020), Schmitt-Grohe et al. (2020), and Almagro and Orane-Hutchinson (2020) account for the racial dimension while looking at the demographic and socioeconomic correlates of the COVID-19 epidemics, with a focus on testing incidence and infections, but not deaths, across New York City zip codes. Across US counties, Desmet and Wacziarg (2020) find a positive correlation between the shares of African Americans and Hispanics and both the number of cases and the number of deaths, a finding which is confirmed by McLaren (2020) for African Americans’ deaths even after controlling for education, occupation, and commuting patterns. Using the 2017 wave of the Panel Study of Income Dynamics to examine the prevalence of specific health conditions, Wiemers et al. (2020) show evidence of large disparities across race-ethnicity and socioeconomic status in the prevalence of conditions which are associated with the risk of severe complications from COVID-19. Using CPS microdata on unemployment, Couch et al. (2020) show that African Americans were only slightly disproportionately impacted by COVID-19, while Latin Americans were hardly hit. With reference to the UK, Platt and Warwick (2020), Sa (2020), and White and Nafilyan (2020) report descriptive evidence on vulnerability factors, infections, and deaths, for ethnic minorities including Black Caribbean and Black African.^5^

Our second contribution is to research on the long-term influence of redlining policies, which have been the object of investigation not only within the field of economics but also within medicine, history, and law. Zenou and Boccard (2000), Appel and Nickerson (2016), Krimmel (2018), Aaronson et al. (2017), Mitchell and Franco (2018), and Anders (2019) respectively look at the effects on unemployment, home prices, homeownership, racial segregation, inequality, and crime. Krieger et al. (2020a, 2020b) and Nardone et al. (2020) respectively associate historical redlining with higher risk of cancer, preterm birth, and asthma, suggesting that this discriminatory practice might be contributing to racial and ethnic disparities.^6^ Jackson (1980, 1985), Hillier (2003, 2005), and Greer (2012, 2014) provide historical accounts of the activities of the HOLC and its influence on American cities. Schill and Wachter (1995) and Nier (1999) offer a legal interpretation of redlining and the induced spatial bias of federal housing law and policy.^7^

A third research stream that is relevant to our approach has looked at other pandemics. The long-term determinants of the HIV/AIDS pandemic, with special attention to women, who represent within Africa the most vulnerable group, have been studied by Anderson (2018), who links the higher female HIV rates to the tradition of common law, while Bertocchi and Dimico (2019), Loper (2019), and Cage and Rueda (2020) respectively refer to the slave trade and the associated diffusion of polygyny, the practice of matrilinearity in ancestral societies, and the influence of the Christian missions established during the colonial period. The long-term economic, social and cultural consequences of the “Spanish” influenza have been studied by Almond (2006), Karlsson et al. (2014), Lin and Liu (2014), Helgertz and Bengtsson (2019), Aassve et al. (2020), and Guimbeau et al. (2020), while Richardson and McBride (2009), Voitglander and Voth (2012), Jedwab et al. (2016, 2019), and Alfani and Murphy (2017) have studied the Black Death.

## 3 Historical background

### 3.1 Redlining

The Home Owners Loan Corporation (HOLC) was created in June 1933 by the US Congress, in the aftermath of the Great Depression and within the first 100 days of the Roosevelt administration, as part of a key package of New Deal policies aimed at rescuing the housing and banking sectors through actions on the mortgage lending market. In the general effort to revive the economy, housing policies were viewed as critical and were therefore assigned a major role. The task of the HOLC was to refinance mortgages in default to prevent foreclosures, as a response to the banking sector turmoil and the drastic fall in home loans and ownership (Harriss, 1951). In 1934 the National Housing Act established the Federal Housing Administration (FHA) to reinforce previous measures and boost the market for single-family homes. With the goal of improving the accuracy of real-estate appraisal and in turn standardizing the process of mortgage lending, credit worthiness assessment, and mortgage support assignment, in 1935 the HOLC was asked to create “Residential Security Maps” of 239 cities that ranked areas on the basis of default risk. The ranking encompassed four levels. The safest areas, mostly consisting of newly-build suburban neighborhoods, were labelled as “Best”, assigned to Type A, and outlined in color green. “Still Desirable” areas were assigned to Type B and outlined in blue. The next two levels included “Definitely Declining” areas, assigned to Type C and outlined in yellow, and “Hazardous” areas assigned to Type D and outlined in red. Because of the color used to highlight to the worst-assessed neighborhoods, those that ended up being de facto denied any mortgage financing, the process came to be known as “redlining”.

The HOLC rankings were based on meticulous assessments and recording of neighborhood characteristics including population growth, class and occupation of the inhabitants, and block-by-block quality of the buildings (type, size, construction material, age, need for repair, occupancy rate, owner-occupancy rate, past and predicted property prices, rents, and sales and rental demand trends). Notably, the share of foreign and black families and the degree to which the neighborhood was deemed to be “infiltrated” were also accurately recorded. Figure A1 in the Appendix reports the area description drafted by the HOLC in 1939 for a D-rated neighborhood in South Chicago, summarily described as *“A 100 per cent negro development. (*…*) A blighted section”*.

To assemble the mapping, the HOLC trained thousands of home appraisers and, in the process, set standards for the development of a new approach to mortgage lending, which were adopted and further refined in the Underwriting Manuals compiled by the FHA (1938). The task for which the HOLC was created was undoubtedly fulfilled, as the agency had a major impact on the subsequent expansion of real estate investments. As documented by Harriss (1951), between 1933 and 1935 the HOLC received almost 1.9 million applications for home mortgage refinancing.^8^ Out of the 54 percent of them that were accepted, the majority involved one- or two-family homes of modest size and value and borrowers of relatively limited income. In the New York region, 44 percent of the properties whose purchase was supported with a loan were located in neighborhoods described as “native white” and 42 percent in “native white and foreign.” The fact that only 1 percent of the applications covered properties in neighborhoods described as “native Negro” is attributed by Harriss to the low percentage of applicants from such areas, which *“doubtless reflects the fact that most Negroes (*…*) lived in rented quarters and did not, therefore, fall within the limits of the HOLC programs*.”

The direct and indeed intended consequences of redlining were to channel credit and investment away from poorer areas and toward more affluent ones. As a result, the slums deteriorated even farther. Over time, the practice is widely believed to have contributed to the exacerbation and persistence of initial inequalities (Douglas Commission, 1968). After the Second World War, racial segregation further intensified with the “white flight” from the inner cities to the suburbs (Boustan, 2011). It was only with the Fair Housing Act of 1968, a provision of the Civil Rights Act, that housing segregation was outlawed, while specific legislation to establish fair lending practices was only enacted in the 1970s with the Equal Credit Opportunity Act (1974) and the Community Reinvestment Act (1977).

Throughout the process, the HOLC maps were deliberately hidden from public view, even though they may have been shared selectively with realtors and lenders (Greer, 2012, 2014). The existence of the maps emerged later and became the subject of investigation of the National Commission on Urban Problems (Douglas Commission, 1968), created by President Johnson in 1965 *“to study building codes, housing codes, zoning, local and Federal tax policies and development standards”* in order to *“provide knowledge that would be useful in dealing with slums, urban growth, sprawl and blight, and to insure decent and durable housing.”* But it was only much later that Jackson (1980, 1985), an urban historian, discovered the HOLC Residential Security Maps in the National Archives, documenting what he describes as a system designed to apply *“notions of ethnic and racial worth to real-estate appraising on an unprecedented scale.”* His discovery spurred a renewed research effort aimed at identifying redlining as a key factor in perpetuating racial disparities that are still observed up to the present day.

### 3.2 Cook County, Illinois

With over five million residents in 2019, Cook County is the most populous county in Illinois and the second-most-populous county in the US after Los Angeles County, California. Over 40 percent of all residents of Illinois live in Cook County. The largest of the county’s 135 municipalities is the City of Chicago—the third-most-populous US city—followed by the City of Evanston. Overall, Cook County is highly urbanized and densely populated. According to the United States Census Bureau,^9^ in 2019 non-Hispanic whites, Hispanics, African Americans, and Asians were the most represented racial and ethnic groups, respectively with 42.0, 25.6, 23.8, and 7.9 percent of the population,^10^ A 21.6 percent share of the population was under age 18 and 15.1 percent was above 65. A 13.8 fraction of the population, higher than the national average, was below the poverty line. Health status disparities between black and white populations widened in Chicago between 1990 and 2005 (Orsi et al., 2010). Politically, the county is heavily Democratic-leaning, with a 73.9 share of the votes being cast for the Democratic Party in the 2016 presidential elections.

According to the Johns Hopkins University & Medicine Coronavirus Resource Center,^11^ as of June 16, 2020 Cook County ranked third among US counties, after Kings and Queens in New York, in terms of COVID-19 related deaths, with over 4,000 deaths, and first in terms of confirmed cases, with over 80,000 cases. A strict stay-at-home order was issued by Illinois Governor Pritzker on March 20 (effective March 21), four days after the first COVID-19 related death, when the death toll was still limited to five.

Turning to the urban history of Cook County in the aftermath of the Great Depression, in his 1933 dissertation on the evolution of land values in Chicago Hoyt (1933)—before joining the FHA in 1934 as Principal Housing Economist—provided a ranking of races and nationalities with respect to their beneficial effect upon land values. While acknowledging that such an effect may have been reflecting racial prejudice, he ranked Anglo-Saxons and Northern Europeans at the top and Negroes, followed by Mexicans, at the bottom. He also produced a map of Chicago (Figure 1, left panel) reporting the areas occupied by predominant groups among the most recent immigrant waves, namely Negro, Italian, Polish, Jewish, and Czechoslovakian newcomers. As the figure shows, blacks were concentrated in the so-called “Black Belt” on Chicago’s South Side, where they were forced to settle from the beginning of the Great Migration, facing squalid housing conditions and extremely high population densities (Greer, 2014).

**Figure 1:**
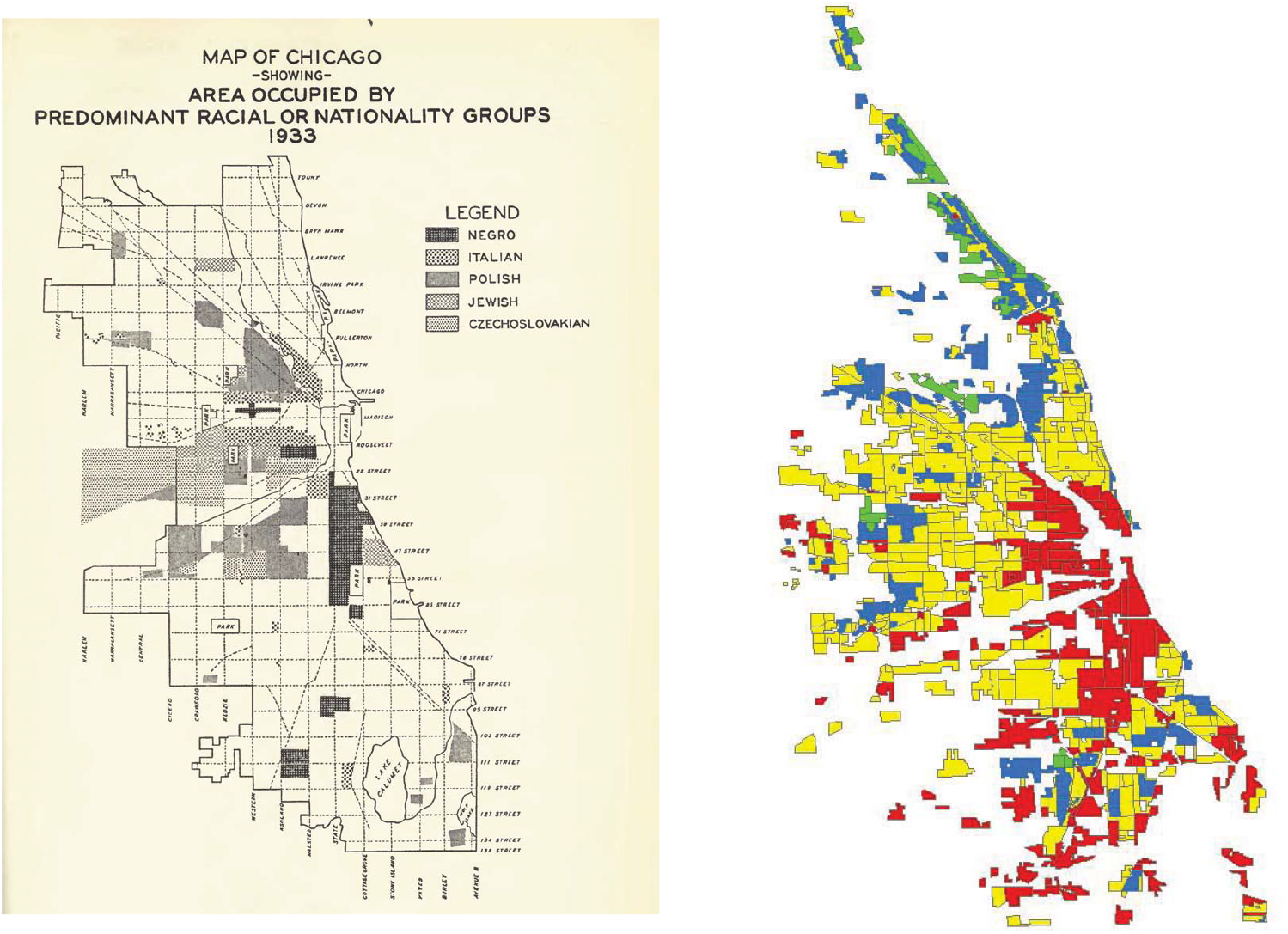
Historical Maps of the Chicago Area. *Note:* The figure shows, on the left panel, the “Map of Chicago Showing Area Occupied by Predominant Racial or Nationality Groups, 1933” (Hoyt, 1933) and, on the right panel, the HOLC maps for the Chicago area (Nelson et al., 2020), with green, blue, yellow, and red denoting respectively grade A, B, C, and D neighborhoods.

By 1940, a large portion of Cook County was mapped by the HOLC. On the right panel, Figure 1 shows the HOLC areas of the Chicago area as rendered in the Mapping Inequality: Redlining in New Deal America 1935-1940 dataset by the University of Richmond.^12^ As in the other American cities, the geography of redlining had a clear racial connotation. The figure reveals that the same areas inhabited by blacks in the Hoyt map were assigned the lowest grade and highlighted in red. Indeed those blocks were characterized by houses lacking basic amenities such as access to water and heating (Greer, 2014). Krimmel (2018) estimates that, by 1940, 98 percent of the relatively small share of blacks living in the city, about 8 percent, were redlined.

The programs that were implemented by the agency were welcome with wide support from the Chicago white press, and were originally only weakly opposed by the black press, including the Chicago Defender. Nevertheless, the disinvestment induced by the HOLC policies had consequences that extended well past the Second World War, with white families easily obtaining mortgage insurance to move to the suburbs, and black ones being relegated in blighted neighborhoods where the only financing opportunity consisted in exploitative installment land contracts (Greer, 2012, 2014). As a result, as of 1967, in the words of the Douglas Commission (1968) Chicago was hosting the most notorious slums in the country.

## 4 Data

The Cook County Medical Examiner’s Officer^13^ has been reporting individual COVID-19 related deaths daily since March 16, 2020, the date of the first fatality recorded in the county.^14^ The declared goal of the initiative is to provide direct access to critical facts that can allow to identify communities that are most severely impacted by the virus. We employ data recorded up to June 16, that we downloaded on June 18.

The Medical Examiner’s Office reports those deaths that are under its jurisdiction, including among others those due to diseases constituting a threat to public health.^15^ The information on COVID-19 fatalities coincides with that provided by the Johns Hopkins University & Medicine Coronavirus Resource Center.^16^

The race, gender, age, comorbidities, and residence (home address, city, zip code, and geographical coordinates) of each dead individual are also provided by the Medical Examiner.^17^ Overall, 4,325 individuals—1,491 of whom black (that is, 34.47 percent)—have died of COVID-19 in Cook County between March 16, 2020 and June 16, 2020. However, geographical coordinates are missing for 704 individuals. This leaves us with a sample of 3,621 deaths—1280 (35.35 percent) of blacks—recorded over the over 14-week span.^18^

Figure 2 plots the number of COVID-19 deaths in our sample for each day, separately for blacks and all other races combined. From March 16 to April 9 the daily number of blacks dying from COVID-19 is above the number for all other races, although the share of the black population is only about one fourth of the total population. By April 9, the cumulative share of blacks who have died from COVID-19 represents almost 58 percent of the total COVID-19 deaths. The daily number of deaths among blacks keeps increasing until mid-April and then starts decreasing at a slow pace. The number of deaths among other races, on the other hand, keeps increasing until mid-May. By May 16, the cumulative share of black COVID-19 deaths is down to about 39 percent, to decrease further to 35.3 percent by June 16. In other words, cumulatively to June 16, blacks in our sample are dying at a rate 1.3 times higher than their 27 percent sample share in the population.

**Figure 2:**
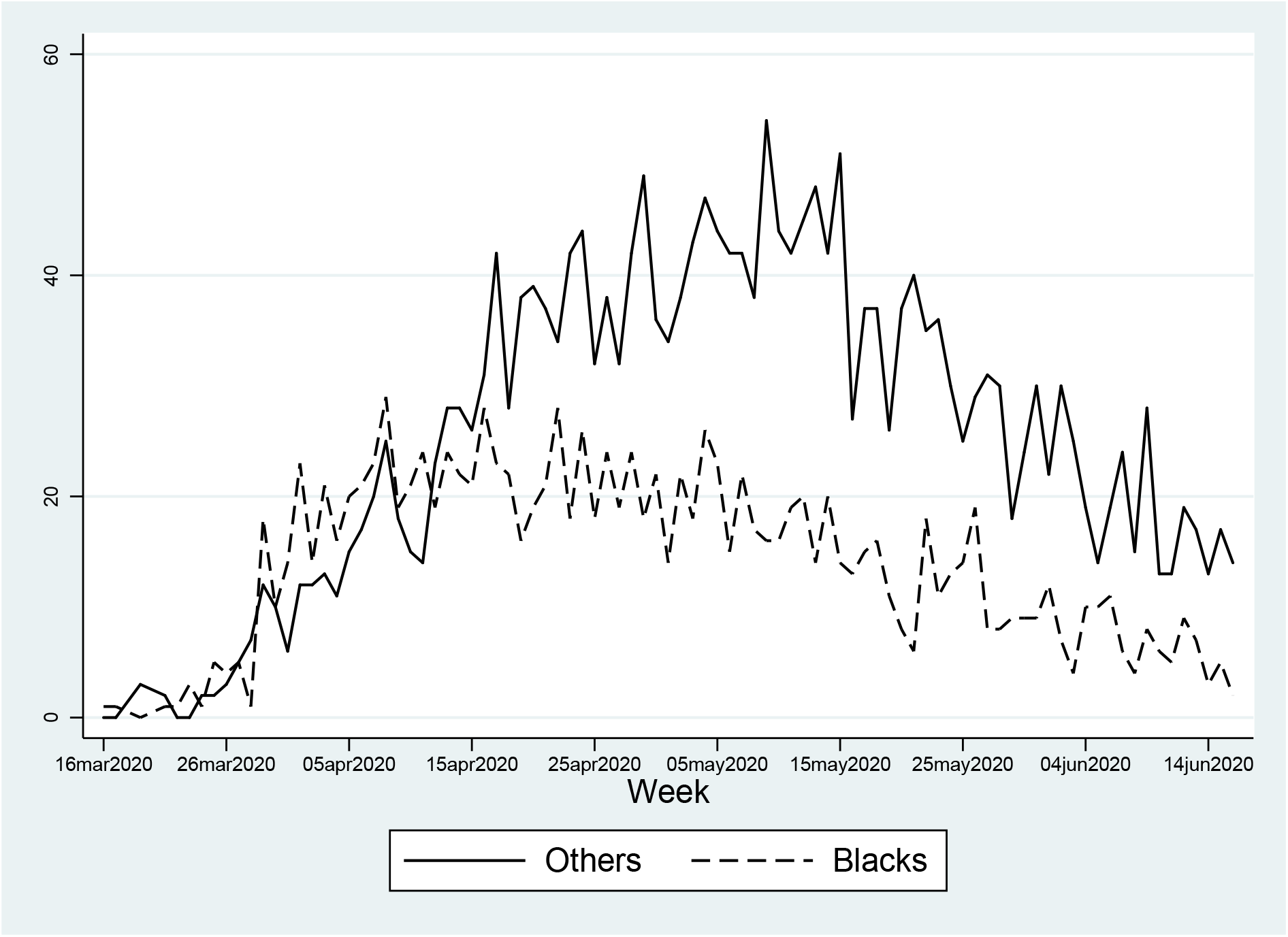
COVID-19 Deaths, by Race - Cook County, March 16-June 16, 2020. *Note:* The figure reports the number of COVID-19 related deaths by day, separately for blacks and all other races combined.

The above data clearly show that blacks are overrepresented in terms of COVID-19 deaths. Furthermore, the data document that blacks likely became infected, and eventually died, before the rest of the population, with a consequent decline in the share of cumulative black deaths as the epidemic followed its course—a trend that has been overlooked by media and public bodies reports.^19^

Figure A2 shows the distribution of deaths for blacks and all other races combined, by age group.^20^ Blacks display a much larger number of deaths in their 60s and 70s, and a lower number past their 80s. Nevertheless, the age distribution below age 60 is quite similar. Overall, the graph confirms a large number of fatalities within the elderly population, a phenomenon which has been largely documented. Figure A3 shows the breakdown by gender among the two groups. Compared to other groups, blacks have a much higher probability of death among women (almost 48 percent against 39 percent).^21^

We extract information on comorbidities by generating a set of 14 dummy variables that take value one (and zero otherwise) when an individual who died from COVID-19 was affected by diabetes and/or asthma, liver disease, cancer, hypertension, kidney disease, obesity, respiratory diseases, neuro-cardiac diseases, neuro-respiratory diseases, asplenia, immunodeficiency, transplant, and heart diseases.^22^ Figure A4 shows the distribution of deaths by comorbidity and race. Diabetes and hypertension are by far the two most common comorbidities. For both, blacks are more likely to suffer from them than the other races combined.

We spatially merge the death data from the Medical Examiner with census block group boundary files^23^ and with the redlining maps produced by HOLC and georeferenced by the University of Richmond.^24^ Figure 3 shows the result of the spatial merge. Each individual COVID-19 death is mapped into a specific block group and HOLC area using the georeferenced home address of the deceased. The map highlights block group boundaries, while HOLC areas are identified by the colour, with green, blue, yellow, and red denoting respectively grade A, B, C, and D.

**Figure 3:**
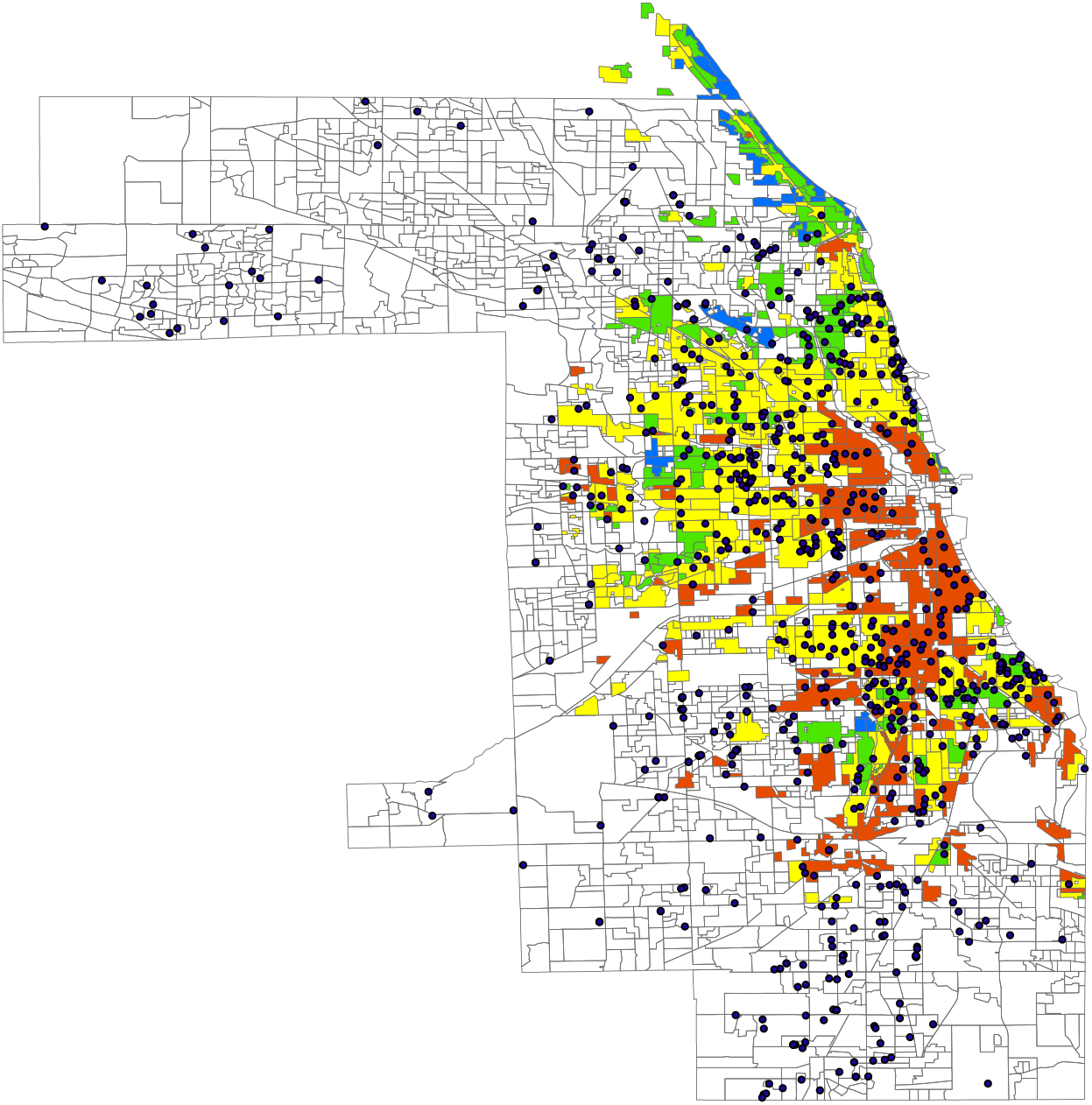
Cook County Map and Total COVID-19 Deaths, March 16-June 16, 2020. *Note:* The map reports census block group boundaries and HOLC areas, with green, blue, yellow, and red denoting respectively grade A, B, C, and D neighborhoods.

From the Cook County Government^25^ we also obtain census tract level data on socioe-conomic characteristics (i.e., age, education, personal income, unemployment, population, and racial groups), averaged over the period 2014-2018. We match these data using for each block group the information available for the corresponding tract.

Figure A5 illustrates the share of black deaths by HOLC grade, only for census tracts where a COVID-19 death has been reported. In D-ranked neighborhoods blacks represent the highest share of the dead, while no black death is reported in those ranked A. Figure A6 illustrates the share of blacks in the population by HOLC grade. With the only exception of A-ranked neighborhoods, the share of the black population is lower than the corresponding share of black deaths shown in Figure A5. This preliminary evidence supports a role for redlining in determining segregation.

We complement the dataset with distance (measured in degrees) from a hospital and a nursing home.^26^ We interpret the latter as a proxy for the likelihood that a COVID-19 death occurred in a nursing home.^27^

Lastly, for the heterogeneity analysis in Sub-section 8.2, we employ the CDC’s Social Vulnerability Index (SVI) dataset,^28^ created by the Geospatial Research, Analysis & Services Program (GRASP) run by the Agency for Toxic Substances and Disease Registry (ATSDR) with the scope of helping emergency response planners and public health officials in the face of a hazardous event, such as a natural disaster (e.g., a tornado or epidemic) or a human-made event (e.g., an oil spill). Social vulnerability refers to those factors that may affect the resilience of a geographical area to such an event. The source of the data used to obtain these indices is the American Community Survey, that provides averages for geographic areas over the period 2014-2018 at a census tract level. The SVI dataset include 15 characteristics, grouped into four indices: Socioeconomic Status (comprising income, poverty, employment, and education variables), Household Composition/Disability (comprising age, single parenting, and disability variables), Minority Status/Language (comprising race, ethnicity, and English language proficiency variables), and Housing/Transportation (comprising housing structure, crowding, and vehicle access variables). A general measure of social vulnerability, which summarizes the above four indices, is also provided.^29^

Tables A1 and A2 in the Appendix reports variable definitions and sources, for the cross section and the panel respectively.

## 5 Cross-sectional evidence

We start our analysis by exploiting cross-sectional information about individual deaths from COVID-19 occurring in Cook County between March 16, 2020 and June 16, 2020. Summary statistics for the cross-sectional sample are reported in Table A3.

A preliminary warning is in order, since the sample is clearly self-selected, as it includes only individuals who have died of COVID-19 and therefore exhibiting specific characteristics, which are precisely those that tend to be more prevalent among blacks.

Keeping in mind the sample selection problem afflicting the cross-sectional sample, in order to assess whether residence in a historically redlined area is a predictor of the probability that an individual that dies from COVID-19 is black, we employ as the outcome variable a dummy taking value one if the individual that died from COVID-19 in Cook County between March 16 and June 16 is reported to be black, and zero otherwise. Table A3 shows that the probability of a black death is 35.3 percent.

Table A3 also indicates, for each HOLC area, the probability that a COVID-19 death has occurred in that area. For instance, the probability that a COVID-19 death has occurred in A-graded neighborhoods is 0.2 percent.^30^ Mean age is between 60 and 70 and 42 percent of the dead are females. Among pre-existing conditions, the disease with highest prevalence is hypertension, that affects 52 percent of the sample, followed by diabetes with nearly 41 percent.

The empirical model aims at exploiting the cross-sectional variation in the mortality of blacks across HOLC areas. Formally, we estimate the following model:

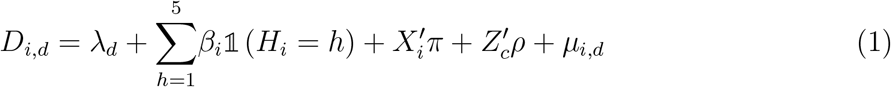

where

*D*_*i,d*_ is a dummy taking value one if individual *i* that died from COVID-19 in day *d* is black (and zero otherwise); *λ*_*d*_ represent day fixed effects that are meant to capture the daily variation in the number of deaths; 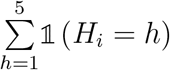 are a full set of dummies denoting the four HOLC-graded areas, from A to D, plus the ungraded area, where individual *i* used to reside; the vector 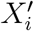 includes a set of individual characteristics (age group, gender, comorbidities, and distance from the closest hospital and nursing home); the vector 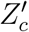 includes a set of socioeconomic characteristics at tract level (the logarithmic of population and the shares of the population of age 18-64, over 65, without a high school diploma, and black); *µ*_*i,d*_ is the error term which we cluster at day level.

Table 1 reports OLS estimates for eight variants of Equation 1. In Model 1 we only control for HOLC (the omitted area is the ungraded area, No HOLC) and day fixed effects. In Model 2 we add demographic information on age and gender, in Model 3 comorbidities, in Model 4 population, in Model 5 the shares of the population aged 18-64 and over 65, in Model 6 the share of the population without a high school diploma, and in Model 7 distance from hospital and nursing home. In all models, the fact that an individual that died from COVID-19 used to live in HOLC areas D and C is positively and significantly associated with the probability that the individual is black, while the opposite is true for area A. In other words, black mortality is much larger in low-graded areas. To be noticed is that the effect is statistically equally significant for areas C and D, even though the size of the coefficient is larger for D.^31^ In terms of magnitudes, relative to the sample mean (35.3), the probability that an individual that died from COVID-19 is black is over 24 percent larger in area C and 26 percent larger in area D. However, when in Model 8 we also add the share of blacks in the census tract, having been a resident in such areas is no longer a predictor of the dead individual’s racial group, suggesting that indeed historical redlining induced segregation along racial lines, that fully absorbs the influence of the former.

**Table 1:** Black COVID-19 Death, Cross-Sectional Results - Cook County, March 16-June 16, 2020.

|  | Black COVID-19 Death |  |  |  |  |  |  |  |
| --- | --- | --- | --- | --- | --- | --- | --- | --- |
|  | (1) | (2) | (3) | (4) | (5) | (6) | (7) | (8) |
| HOLC A | -0.3808***<br>(0.0640) | -0.3652***<br>(0.0720) | -0.3244***<br>(0.0757) | -0.3261***<br>(0.0800) | -0.3247***<br>(0.0811) | -0.3636***<br>(0.0798) | -0.3514***<br>(0.0751) | -0.1250***<br>(0.0424) |
| HOLC B | 0.0389<br>(0.0329) | 0.0360<br>(0.0334) | 0.0399<br>(0.0327) | 0.0373<br>(0.0323) | 0.0355<br>(0.0327) | 0.0303<br>(0.0322) | 0.0072<br>(0.0316) | 0.0169<br>(0.0250) |
| HOLC C | 0.0800***<br>(0.0177) | 0.0817***<br>(0.0185) | 0.0830***<br>(0.0181) | 0.0677***<br>(0.0175) | 0.0716***<br>(0.0202) | 0.1176***<br>(0.0216) | 0.0848***<br>(0.0234) | -0.0146<br>(0.0182) |
| HOLC D | 0.1752***<br>(0.0233) | 0.1769***<br>(0.0238) | 0.1745***<br>(0.0231) | 0.0833***<br>(0.0255) | 0.0852***<br>(0.0266) | 0.1232***<br>(0.0281) | 0.0922***<br>(0.0297) | 0.0106<br>(0.0222) |
| Age Groups | No | Yes | Yes | Yes | Yes | Yes | Yes | Yes |
| Female | No | Yes | Yes | Yes | Yes | No | Yes | Yes |
| Comorbidities | No | No | Yes | Yes | Yes | Yes | Yes | Yes |
| Population (log) | No | No | No | Yes | Yes | Yes | Yes | Yes |
| Share Aged 18-64 | No | No | No | No | Yes | Yes | Yes | Yes |
| Share Aged 65+ | No | No | No | No | Yes | Yes | Yes | Yes |
| Share With No High School | No | No | No | No | No | Yes | Yes | Yes |
| Distance From Hospital | No | No | No | No | No | No | Yes | Yes |
| Distance From Nursing Home | No | No | No | No | No | No | Yes | Yes |
| Black Share | No | No | No | No | No | No | No | Yes |
| Day FE | Yes | Yes | Yes | Yes | Yes | Yes | Yes | Yes |
| Adj.R-squared | 0.056 | 0.069 | 0.090 | 0.111 | 0.111 | 0.132 | 0.144 | 0.475 |
| Observations | 3618 | 3616 | 3616 | 3616 | 3616 | 3616 | 3616 | 3616 |

*Note:* The dependent variable is a dummy variable that takes value one if an individual who died from COVID-19 is black, an zero otherwise. The omitted area is the one ungraded by the HOLC. Robust standard errors clustered at a day level in parentheses: \*\*\* p<0.01, \*\* p<0.05, \* p<0.1.

It is instructive to report how the dependent variable is affected by other covariates (all coefficients are reported in Table A4). For instance, even in the full specification where we control for the black share, its likelihood is higher for females, which means that, within the black population, women have a relatively higher chance to die from COVID-19, relative to the chance they have within the total population. Furthermore, pre-existing conditions, in particular hypertension and kidney and respiratory diseases among those with high prevalence, exert a significantly positive effect on the dependent variable. Distance from hospital decreases the probability of a black death although it becomes insignificant once we control for the share of blacks. Distance from a nursing home, that is, a lower probability that a death has occurred in a nursing home, increases the probability of a black death but, once race is accounted for, the sign of the association is reversed, which can be attributed to a negative correlation between the presence of nursing homes and the black share of a neighborhood. Nevertheless, neither comorbidities nor the other demographic and socioeconomic factors fully absorb the influence of redlining, at least until the black share is accounted for.

Overall, the cross-sectional results confirm that yellow and redlined areas are associated with a higher incidence of COVID-19 deaths among blacks, and that the difference between redlined and yellowlined areas is blurred. However, since blacks are concentrated in the areas where mortality is higher, the cross-sectional analysis solely based on information about those that died from COVID-19 is severely biased because of sample selection. This limitation motivates the event study approach we introduce in the next section.

## 6 An event study approach

### 6.1 Empirical strategy

The cross-sectional approach only provides simple correlations with no causal implications because of issues related to sample selection and omitted variable bias, which may confound pre-existing differences between areas with and without COVID-19 mortality. To reduce such biases, we assemble a weekly balanced panel at block group level over the period from January 1, 2020 to June 16, 2020.^32^ For these 24 weeks, we collect information on all types of deaths, that is, from COVID-19 and other causes under the jurisdiction of the Cook County Medical Examiner’s Officer. Between January 1 and June 16, 6,753 deaths—of which 5,492 after March 16^33^—are reported, each associated with home residence, geographical coordinates, and individual characteristics.^34^ Again we map individual deaths into census block groups and HOLC areas and then we aggregate them at a census block group and by week. Therefore, for each block group-week, we gather information on reported number of deaths (if any) for a given block group in any of the 24 weeks from January 1 to June 16. Figure 4 illustrates the spatial merge. The data is then merged with the previously described socioeconomic characteristics provided at census tract level by the Cook County Government and with proxies for social vulnerability to shocks from the CDC.

**Figure 4:**
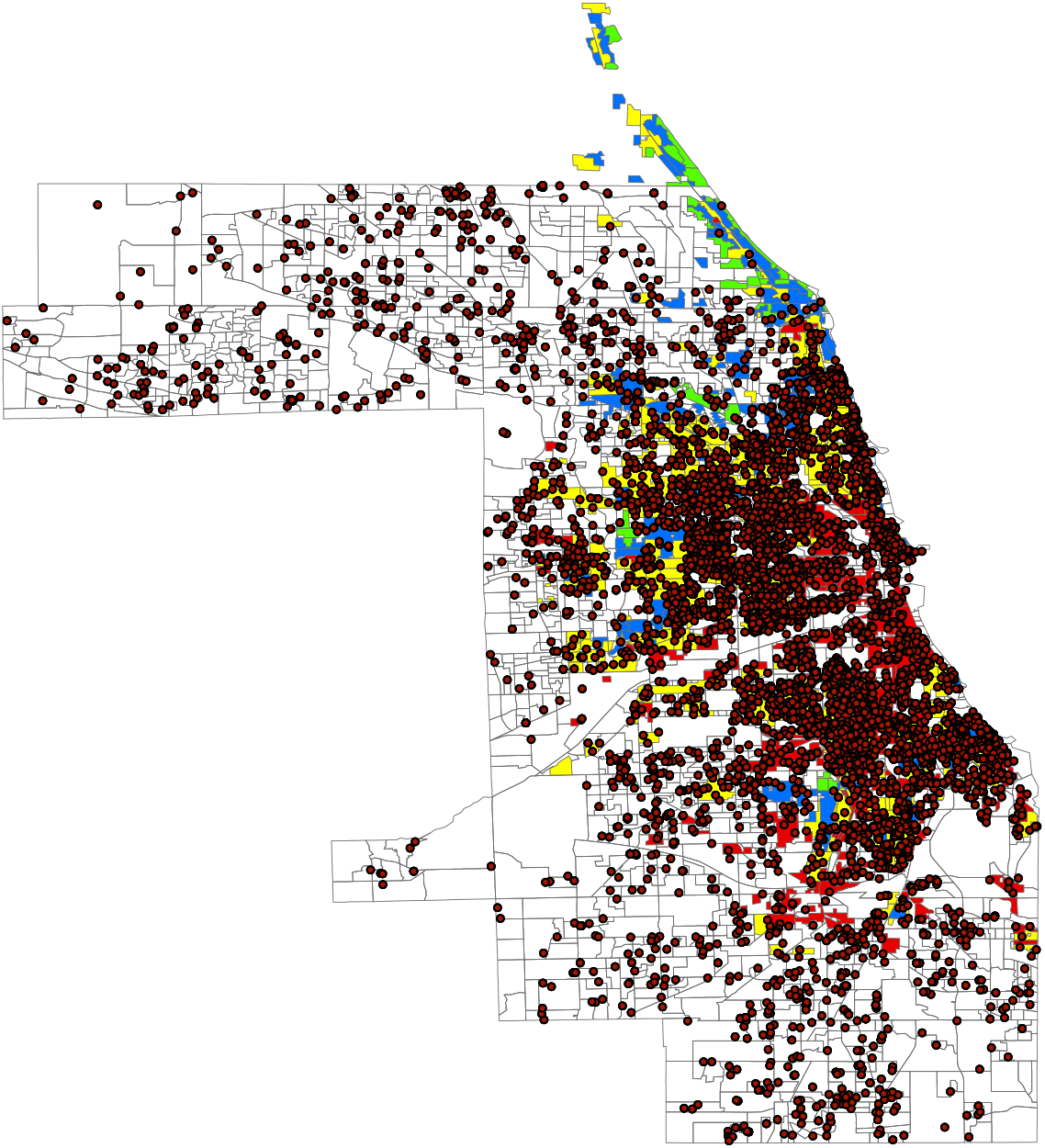
Cook County Map and Total Deaths, January 1-June 16, 2020. *Note:* The map reports census block group boundaries and HOLC areas, with green, blue, yellow, and red denoting respectively grade A, B, C, and D neighborhoods.

Aggregating at a block group level has clear advantages in terms of identification, since it allows to test for pre-treatment differences while controlling for block group fixed effects which should filter out the effect of all socioeconomic factors which do not change over this 24-week period. However, the aggregation implies that a given block group may overlap multiple HOLC neighborhoods, possibly assigned to different grades. Figure A7 zooms into the map and displays as an example four block groups, each including neighborhoods that belong to three different HOLC types. In each block group, two neighborhoods are graded C and outlined in yellow, and one is ungraded (the white one). The distribution of HOLC neighborhoods by block group is shown in Figure A8. Almost 80 percent of the block groups in our sample include no more than two HOLC neighborhoods (potentially with the same HOLC grade) and almost 95 percent of the block groups include no more than three, with the rest of the block groups including up to ten different HOLC neighborhoods. Thus, in order to determine treated block groups, we will focus on those for which the majority of the surface area falls into HOLC areas graded either C or D. This is consistent with the cross-sectional evidence, according to which redlined and yellowlined areas exert a similar effect on the probability that an individual that died from COVID-19 is black, and also with the evidence reported by

Aaronson et al. (2017), who stress the relevance of yellowlining for racial segregation. The treatment therefore will capture whether resilience to external shocks, namely, COVID-19, is affected by the historical policies which have favored racial segregation and economic discrimination.

Figure 5 plots the mortality rate from any cause, separately for blacks and all other races combined, for each week in the sample.^35^ From the beginning of the sample period, the mortality rate is higher for blacks. For the first 10 weeks, before the epidemic outbreak, average mortality for blacks fluctuates between 0.04 and 0.05 deaths per thousand versus around 0.02 per thousand for other races. Starting from the eleventh week (i.e., the week of March 13, that is the week when the first COVID-19 death is recorded on March 16), mortality soars among both groups, but much more steeply so for blacks. Both curves seem to be reaching a plateau by week 18 and start converging toward to pre-COVID mortality rates by week 24.

**Figure 5:**
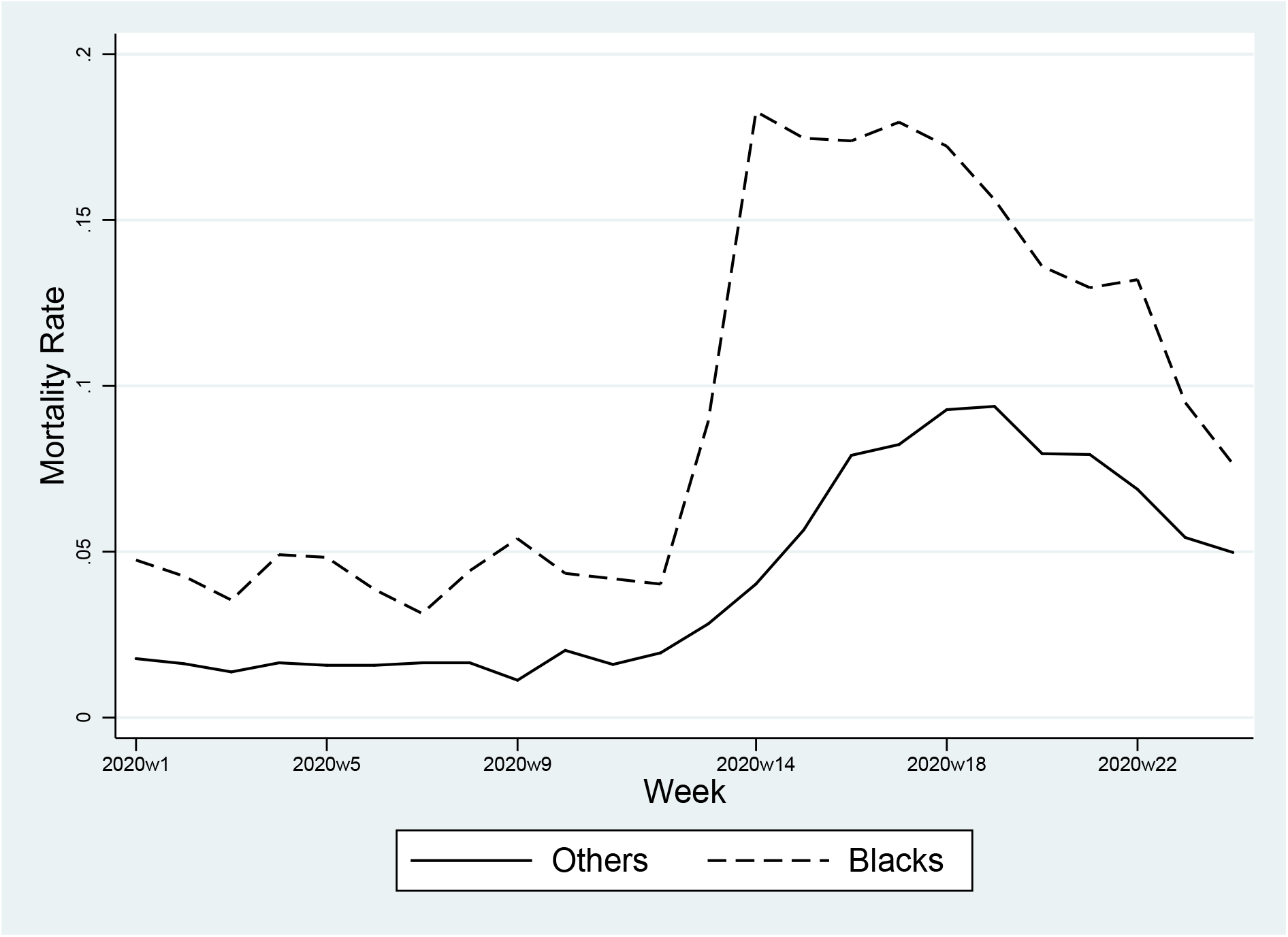
Mortality Rate, by Race - Cook County, January 1-June 16, 2020. *Note:* The figure reports mortality rates from any cause of death by week, separately for blacks and all other races combined.

Summary statistics for the panel dataset, including socioeconomic covariates, are reported in Table A5. On average, for each block group-week in the sample, 0.070 deaths—0.029 (41.4 percent) of blacks—are recorded during the period under examination. The probability of a death in a given block group-week (measured as a dummy variable) is 5.7 percent, while the probability of a black death is 2.5 percent. Treated block groups (i.e., those predominantly graded C or D) represent almost 62 percent of the sample. Social vulnerability indices represent percentile ranking of geographical areas depending on 15 categories and are bounded between 0 and 1, with 1 representing the maximal level of vulnerability (the last percentile). A socioeconomic vulnerability of 0.54 therefore denotes that the average census tract within Cook County falls approximately within the 50th percentile of the distribution of vulnerability.

Figure A9 summarizes a number of demographic and economic characteristics by HOLC grade. Personal income in red and yellowlined neighborhoods is well below $40,000 and even lower in the latter. A similar pattern emerges for the share of population with no high school diploma, which is highest in C-graded neighborhoods. The share of the black population is highest in redlined neighborhoods (close to 40 percent), while the rate of black mortality peaks in yellowlined ones, with an average of two deaths per thousand of blacks. However, in reading mortality statistics one must consider the fact that the denominator (the black population) is not constant across the four areas. Figure A10 plots the weekly mortality rate for blacks and the other races by HOLC grade. Mortality increases with the outbreak of the COVID-19 epidemic in all areas. The spikes we observe for black mortality in A-graded neighborhoods are due to the very small share of the black population. In neighborhoods belonging to the other three areas, black mortality is always higher than the one for other races and particularly high in yellowlined areas, although one must consider that the black population in yellowlined areas is much smaller than in redlined ones, as shown in the previous figure.

Our goal is to capture the impact of the asymmetric shock introduced by COVID-19 on historically segregated areas, that is, whether in majority C and D block groups, after treatment initiation, deaths deviate more from those recorded in the pre-treatment period. Thus, we estimate variants of the model below:

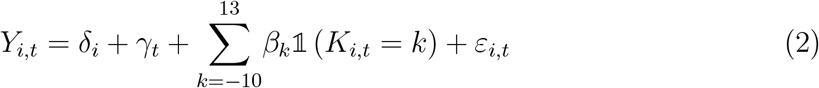

where *Y*_*i,t*_ represents the number of deaths in block group *i* and week *t*;^36^ *δ*_*i*_ and *γ*_*t*_ are block group and week fixed effects meant to control for block group characteristics which are fixed over the 24-week period and to capture natural fluctuations in mortality as well as policies that affect the county uniformly (namely, the lockdown); 𝕝 (*K*_*i,t*_ = *k*) denote treated blocks for *k*_*i,t*_ = *−*10, *−*9, …, 13 periods (i.e., weeks) before and after the treatment kicks in, with the *β*_*k*_s for *k <* 0 corresponding to pre-treatment effects and the *β*_*k*_s for *k ≥* 0 corresponding to the dynamic effects *k* periods relative to the event. We omit the period before the treatment kicks in, i.e., week 10. The error term, *ε*_*i,t*_, is clustered at block group level.

The event study approach outlined in Equation 2 allows to alleviate some of the shortcomings affecting the cross-sectional analysis. The inclusion of the pre-treatment periods allows to test for the parallel trend assumption and therefore for potential secular differences between treated and control groups, as well as the occurrence of self-selection, that may lead to different rates of disease transmission between treated and untreated group. Issues related to sample selection will also be ruled out, since the sample includes the universe of the block groups in the county. Learning and adaptation to the treatment before it kicks in, as it occurs with staggered treatment, is also unlikely given that the treatment period is constant. The only potential source of bias is therefore likely to be related to measurement error, since not all the deaths that occur in Cook County are reported to the Medical Examiner. Measurement error may be more severe in areas with higher mortality but, since this is likely to cause an attenuation bias, the resulting estimator would produce more conservative estimates.

### 6.2 Baseline results

As shown in Figure 1, Cook County was only partially mapped by the HOLC in the 1930s, since the Corporation focused on cities with a population in 1930 above 40,000. It is therefore possible that those neighborhoods which were mapped had completely different characteristics from those which were not, and that these differences in pre-existing conditions may be somewhat correlated with the treatment. To minimize the possibility of biases arising from comparing areas that were already different, we focus on block groups within municipalities which were mapped by the HOLC.^37^

Figure 6 illustrates the dynamic treatment effect on the number of deaths for blacks (Panel a) and all other races (Panel b). The coefficients are least-squares estimates of the *β*_*k*_s and vertical lines represent 95 percent confidence intervals based on standard errors clustered at at block group level. Before the COVID-19 outbreak, the average number of deaths for blacks and all other races in treated block groups is not significantly different from the average number of deaths in non-treated ones. However, after the epidemic shock, mortality in treated block groups increases quite sharply and this is particularly evident among African Americans, for whom the number of deaths in treated block groups increases up to 0.05 per week, an over 17 percent increase with respect to the average number of black deaths by week and by block group. The effect starts picking up in period 4 (toward mid April, when overall mortality sharply increases). Four weeks after the outbreak, the number of deaths among African Americans increases by almost 0.03 per week and continues rising for the next few weeks before it starts fading away toward the end of the 24-week period. By contrast, deaths for other races are not significantly affected by the treatment.^38^

**Figure 6:**
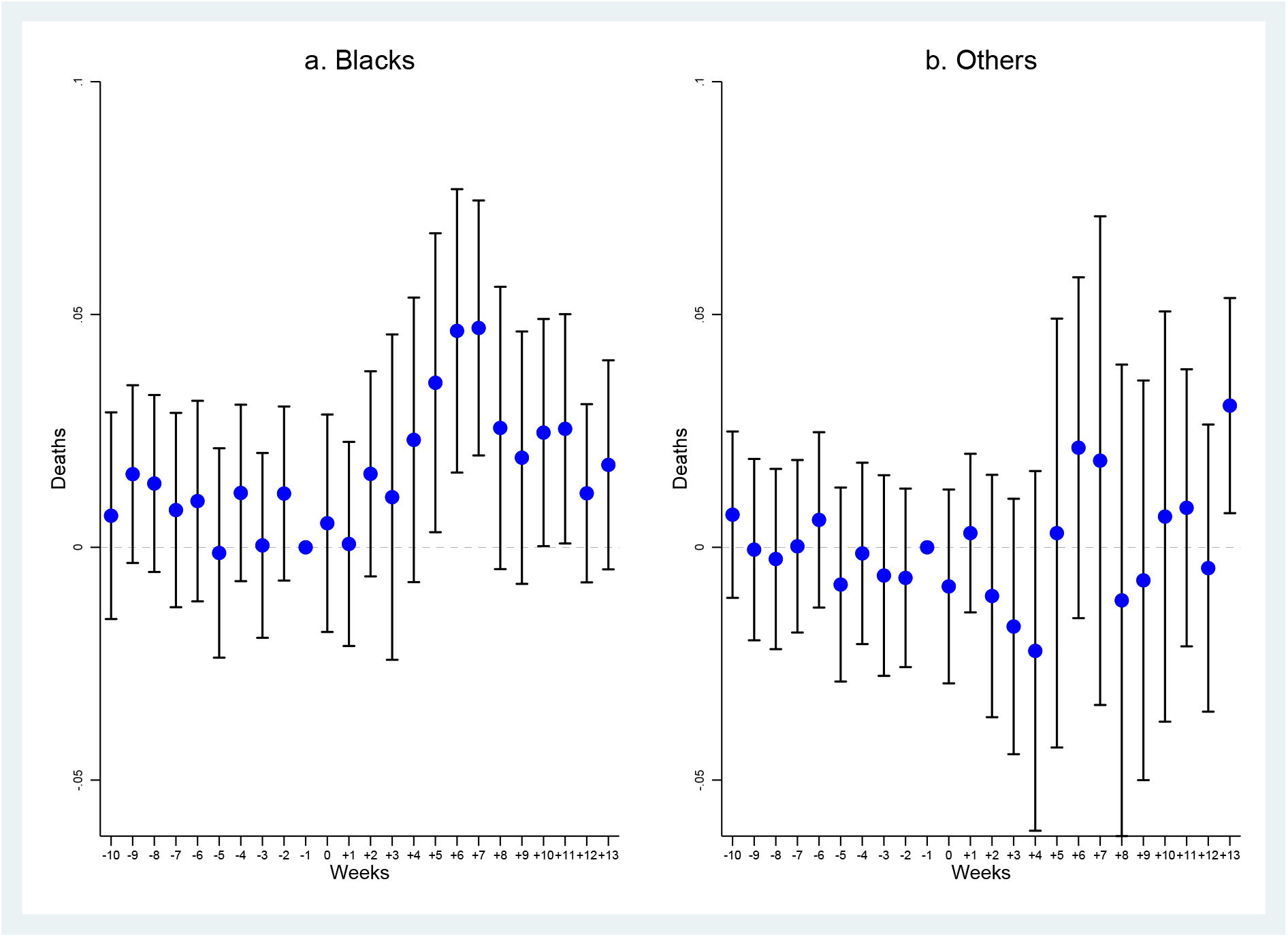
Dynamic Effect of the Treatment on Deaths, by Race - Cook County, January 1-June 16, 2020. *Note:* The dependent variable is number of deaths, of blacks (Panel a) and of other groups (Panel b). The coefficients are least-squares estimates of the *β*_*k*_s. Block group and week fixed effects are included. Vertical lines represent 95 percent confidence intervals based on standard errors clustered at at block group level. The omitted period *k* = *−*1, i.e., week 10.

To quantify the overall effect of the treatment, in Table A6 we estimate the Average Treatment Effect on the Treated (ATET) for blacks and the other races using a simple-difference-in differences method. Overall, we find that the number of black deaths in treated block groups after the epidemic shock (relative to non-treated blocks) increases by 0.015, while the effect for the other races is not statically significant (and negative). If compared to the average number of deaths for the untreated sample in the post-treatment period (i.e., 0.025), the implied magnitude of the estimated effect is an increase in the number of black deaths by almost 60 percent.

## 7 Robustness

### 7.1 Allowing for differential trends

The evidence presented in the previous section reveals an asymmetric effect of the epidemic shock and suggests that the relative resilience to it depends on the level of historical racial segregation induced by loan market discrimination as a result of the HOLC Residential Security Maps. An alternative explanation for the positive effect we detect for the treatment points to potential differential trends which may have placed treated and untreated neighborhoods on different trajectories that then surfaced only when the shock struck. To control for such differential trends, we use tract level socioeconomic and demographic controls, that we interact with week dummies in order to provide the time variation we aim at exploiting. The result of this exercise is reported in Figure A13 for black deaths. In Panel a we interact with week dummies the share of the population aged 65 and above. In Panel b we further add interactions with the shares of Chinese speakers and Hispanics. In Panel c we keep adding differential trends depending on unemployment and income. In the last three plots we insert interactions with the share with no high school diploma (Panel d), the share of blacks (Panel e), and distance from hospital and nursing home (Panel f). Differential trends in income and unemployment partially offset the effect of the treatment, but we can still observe for it a sizeable (although diminished) effect on the number of deaths.

### 7.2 Robustness to aggregation

As mentioned above, aggregating at a block group level allows us to tighten the identification of the effects. However, the definition of the treatment may be blurred by the fact that a block group can overlap multiple HOLC areas (as in the example shown in Figure A7). To explore the extent of issues that may be raising due to our aggregation approach, we perform a sensitivity test and split the sample between block groups including a single HOLC neighborhood and block groups including up to two, up to three, and up to four HOLC neighborhoods. Block groups that only include a single HOLC neighborhood are not affected by treatment definition issues due to aggregation, since each of them falls entirely within a given HOLC category. As a result, comparing results obtained for these block groups with those for block groups that include up to two, three and four neighborhoods will allow us to understand whether aggregation is truly a problem. Reassuringly, the effect of the treatment does not change sensibly whenever we include blocks which include multiple HOLC neighborhoods (Figure A14).

## 8 Heterogeneity

### 8.1 Heterogeneity by HOLC grade

In order to evaluate factors that are potentially correlated with the partitioning of cities by the HOLC and may explain the effect we found, this section goes on to investigate how the effect of the treatment varies with specific characteristics. We start with a heterogeneity analysis by HOLC grade. Specifically, we consider sub-samples of neighborhoods, defined on the basis of their HOLC ranking.

Results are shown in Figure 7. In Panel a we focus only on block groups for which the majority of the surface area falls in either a grade A or a grade D area.^39^ In other words, we compare the best and the worst neighborhoods (according to the HOLC grading scheme). In Panel b we compare the second best neighborhoods (HOLC grade B) with the worst (grade D). In Panel c we compare the best neighborhoods (grade A) with the second worst (grade C) and in Panel d the second best (grade B) with the second worst (grade C). The effect of the treatment is much larger when we compare the worst and the second worst neighborhoods with the best (respectively Panels a and c), and becomes significant from period 2 in Panel a and from period 3 in Panel c. However, even when we compare the worst and the second worst neighborhoods with the second best (respectively Panel b and Panel d), we still observe a quite significant effect of the treatment for several weeks.

**Figure 7:**
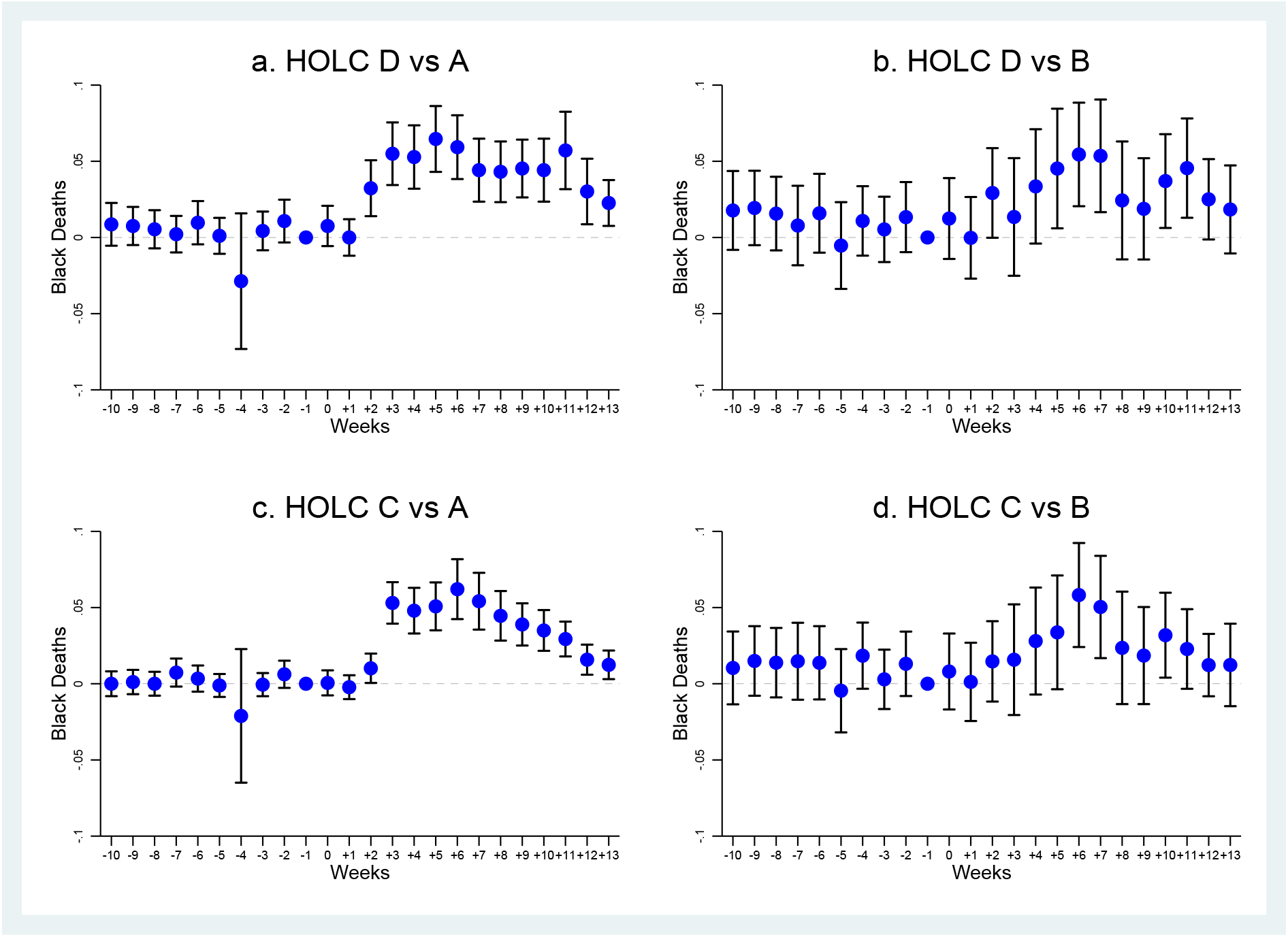
Heterogeneity by HOLC Grade - Cook County, January 1-June 16, 2020. *Note:* The dependent variable is number of black deaths. The coefficients are least-squares estimates of the *β*_*k*_s over samples of block groups for which the majority of the surface area falls in either a grade A or a grade D area (Panel a), in either a grade B or a grade D area (Panel b), in either a grade A or a grade C area (Panel c), and in either a grade B or a grade C area (Panel d). Block group and week fixed effects are included. Vertical lines represent 95 percent confidence intervals based on standard errors clustered at at block group level. The omitted period *k* = *−*1, i.e., week 10.

### 8.2 Heterogeneity by social vulnerability

The social vulnerability indices collected at a census tract level by the CDC allow us to carry out a heterogeneity analysis depending on the level of vulnerability to shocks of specific areas. Thus, in this section we group neighborhoods on the basis of the four distinct dimensions of social vulnerability, as well as some of their components.

Figure 8 shows heterogeneity results by socioeconomic status vulnerability, an index that comprises four components, reflecting percentile scores for personal income, poverty, unemployment, and education. In Panels a and b we split the sample between block groups with a value of the socioeconomic vulnerability index below and above the median. In the next two panels (Panels c and d) we split block groups below median socioeconomic vulnerability between those with a population black share respectively below and above 25 percent.^40^ In the two bottom panels (Panels e and f) we repeat the same exercise for block groups with above median socioeconomic vulnerability. Unsurprisingly, the effect of the treatment is much larger (and significant) in block groups with above median socioeconomic vulnerability (Panel b). The estimated dynamic treatment effect in the sample of block groups below median is nearly zero and not significant (at a 5 percent level), despite the fact that standard errors are exceptionally small. When we split the two sub-samples between those with a share of blacks below (Panels c and e) and above (Panels d and f) 25 percent, some noteworthy differences emerge. The treatment effect is much larger in neighborhoods with a black share above 25 percent, which is not surprising given that the black share is this sub-sample is larger. However, among these neighborhoods, the treatment effect changes quite significantly depending on the level of socioeconomic vulnerability (Panel d vs Panel f). For areas below median vulnerability (Panel d), the post-treatment effect of being red or yellowlined is not that different from the pre-treatment, and in any case not significantly so at a 5 percent level. When, on the other hand, we look at the sample above median vulnerability, that is, at neighborhoods with worse performances, we observe quite a strong treatment effect, which resembles the one we found over the whole sample. These dissimilarities point to a much stronger impact of socioeconomic determinants, rather than genetic and biological one, of black mortality. In other words, neighborhoods with the same black share exhibit different effects of the treatment, and therefore different level of resilience to external shocks, depending on the level of socioeconomic vulnerability.

**Figure 8:**
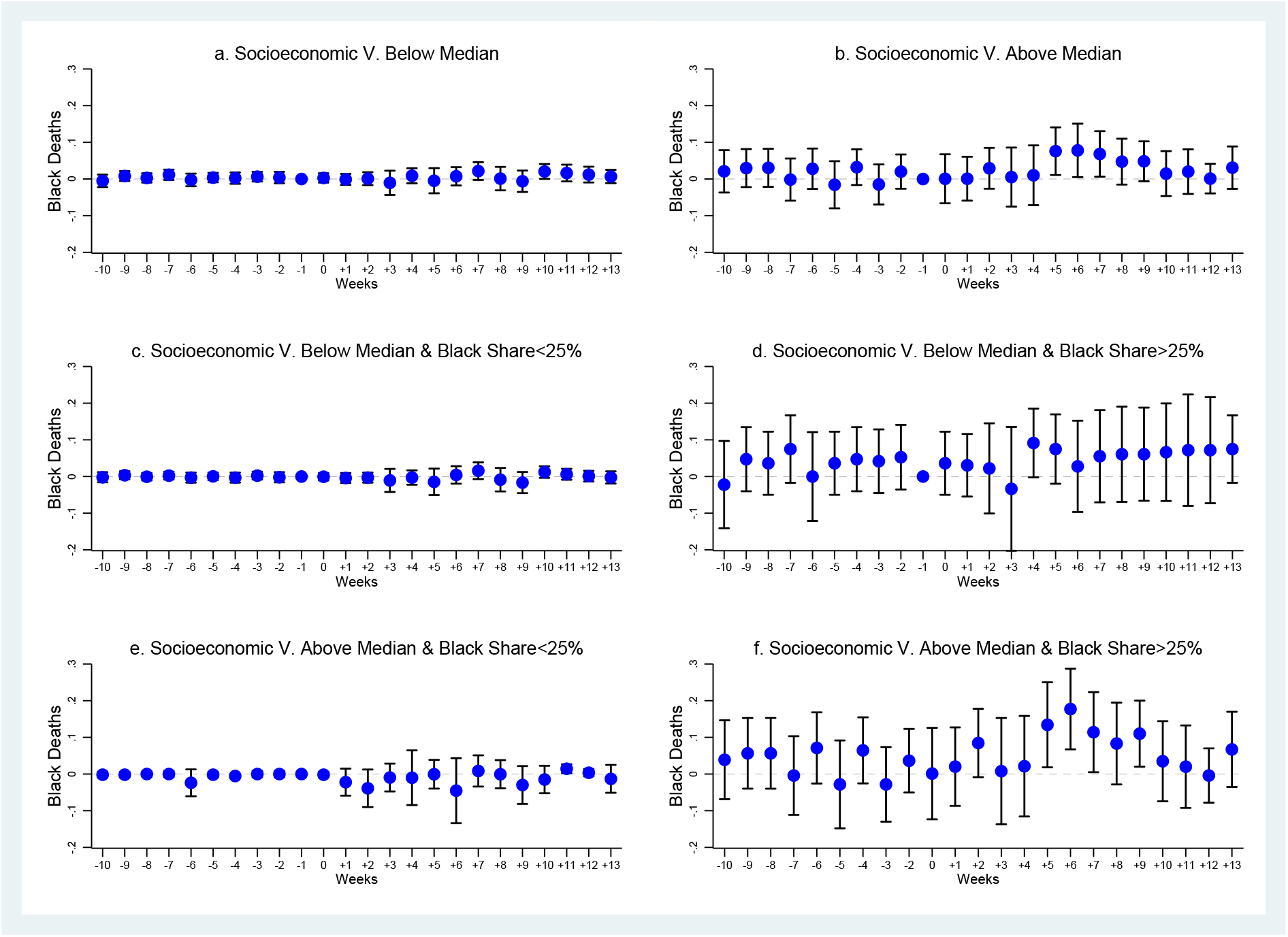
Heterogeneity by Socioeconomic Status Vulnerability - Cook County, January 1-June 16, 2020. *Note:* The dependent variable is number of black deaths. The coefficients are least-squares estimates of the *β*_*k*_s over samples of block groups with socioeconomic status vulnerability below median (Panel a), above median (Panel b), below median and with black share below 25 percent (Panel c), below median and with black share above 25 percent (Panel d), above median and with black share below 25 percent (Panel e), and above median and with black share above 25 percent (Panel f). Block group and week fixed effects are included. Vertical lines represent 95 percent confidence intervals based on standard errors clustered at at block group level. The omitted period *k* = *−*1, i.e., week 10.

To better understand which component of the socioeconomic vulnerability index drives our findings, in Figures A15-A18 we replicate the same analysis focusing, one by one, on its four components, that is personal income, poverty, unemployment, and education. Overall, the effect of the treatment is much stronger in neighborhoods that underperform in all four dimensions and at the same time exhibit a black share above 25 percent. However, when we focus on neighborhoods with a black share above 25 percent, the difference between the worst and the best performing neighborhoods is especially striking for the income and poverty dimensions. The treatment effect for neighborhoods below median income (Figure A15, Panel d) is large and significant, while above median income (Panel f) the treatment effect is not significant in most post-treatment periods. The same occurs for poverty (Figure A16), with a relatively small (and significant in one period only) average treatment effect in the post-treatment period for neighborhoods with a population share below the poverty line smaller than the median (Panel d), and a sizeable effect for those with a larger one (Panel f).

In Figure 9 we repeat the same analysis with a focus on the household composition vulnerability index, which includes four components, that is the shares of the population over 65, below 17, older than 5 with disabilities, and of single parents. Again the effect is much more marked when we focus on block groups with above median household vulnerability and a black share above 25 percent (Panels d and f). Once again, to understand which component of the index drives the effect, in Figure A19 we split the sample between block groups with a share of population aged 65+ below and above the median, while in Figure A19 we do so according to the share of single parents. As expected, aging is quite an important factor. Consistent with the pattern reported for blacks in Figure A2, fatalities among the elderly are more numerous when we look at the sample with a black share above 25 percent (Panels d and f). For the share of single parents in Figure A20, the difference in the estimated effect between Panel d and Panel f is sizeable. In other words, the effect of yellow and redlining is most detrimental when a high share of single parents is combined with a high black share.

**Figure 9:**
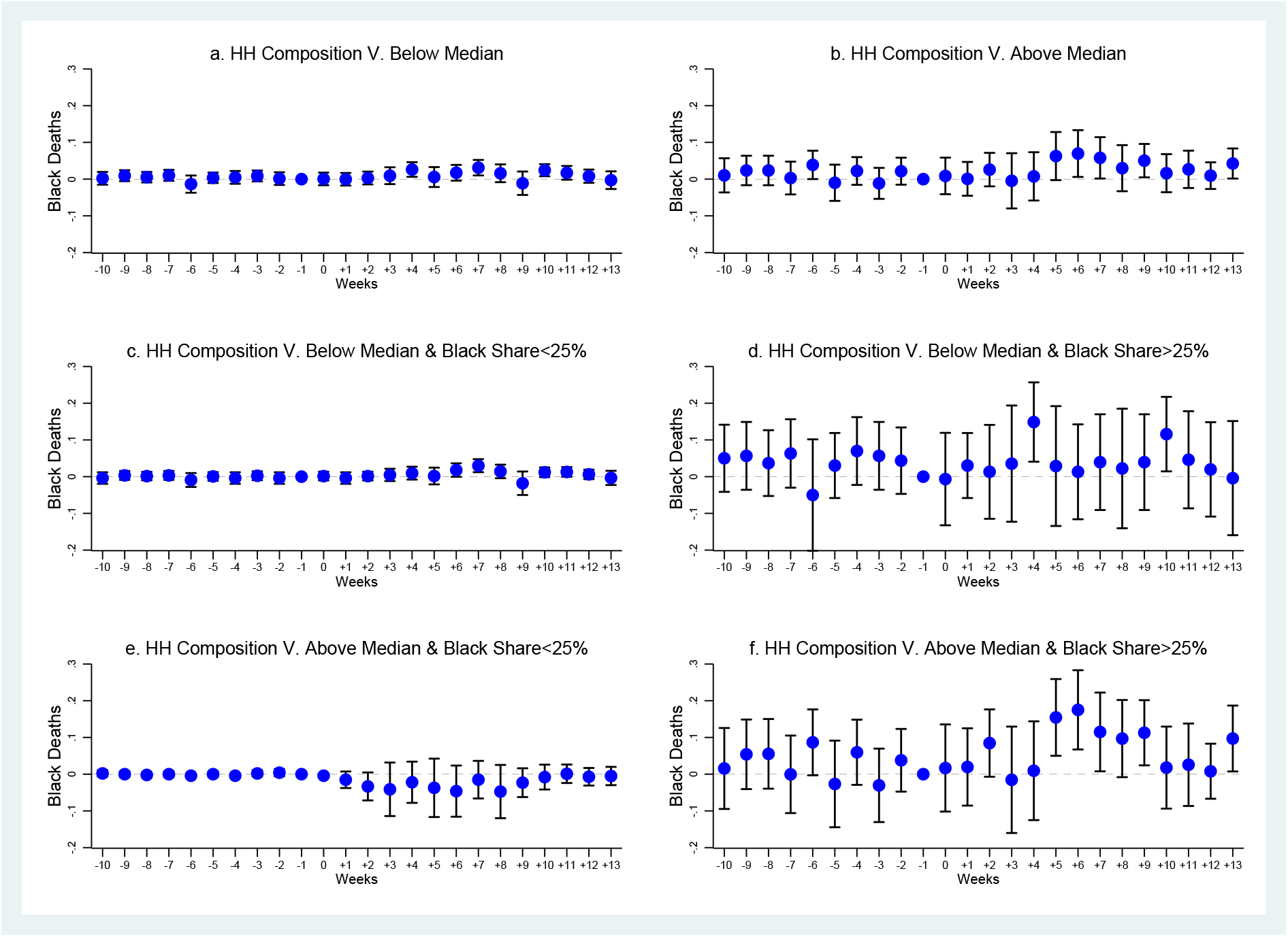
Heterogeneity by Household Composition Vulnerability - Cook County, January 1-June 16, 2020. *Note:* The dependent variable is number of black deaths. The coefficients are least-squares estimates of the *β*_*k*_s over samples of block groups with household composition vulnerability below median (Panel a), above median (Panel b), below median and with black share below 25 percent (Panel c), below median and with black share above 25 percent (Panel d), above median and with black share below 25 percent (Panel e), and above median and with black share above 25 percent (Panel f). Block group and week fixed effects are included. Vertical lines represent 95 percent confidence intervals based on standard errors clustered at at block group level. The omitted period *k* = *−*1, i.e., week 10.

In Figure 10 we repeat the heterogeneity analysis by vulnerability depending on minority status (the index also comprises a measure of English language proficiency) and in Figure 11 by housing/transportation vulnerability (the index comprises percentages of multi-unit structures, mobile homes, a measure of crowding based on the presence of more people than rooms, households with no vehicle, and households in group quarters). Differences in the treatment depending on vulnerability along these two dimensions, and combined with the black share, are still detectable but less pronounced than those we found for socioeconomic status and household composition vulnerability.

**Figure 10:**
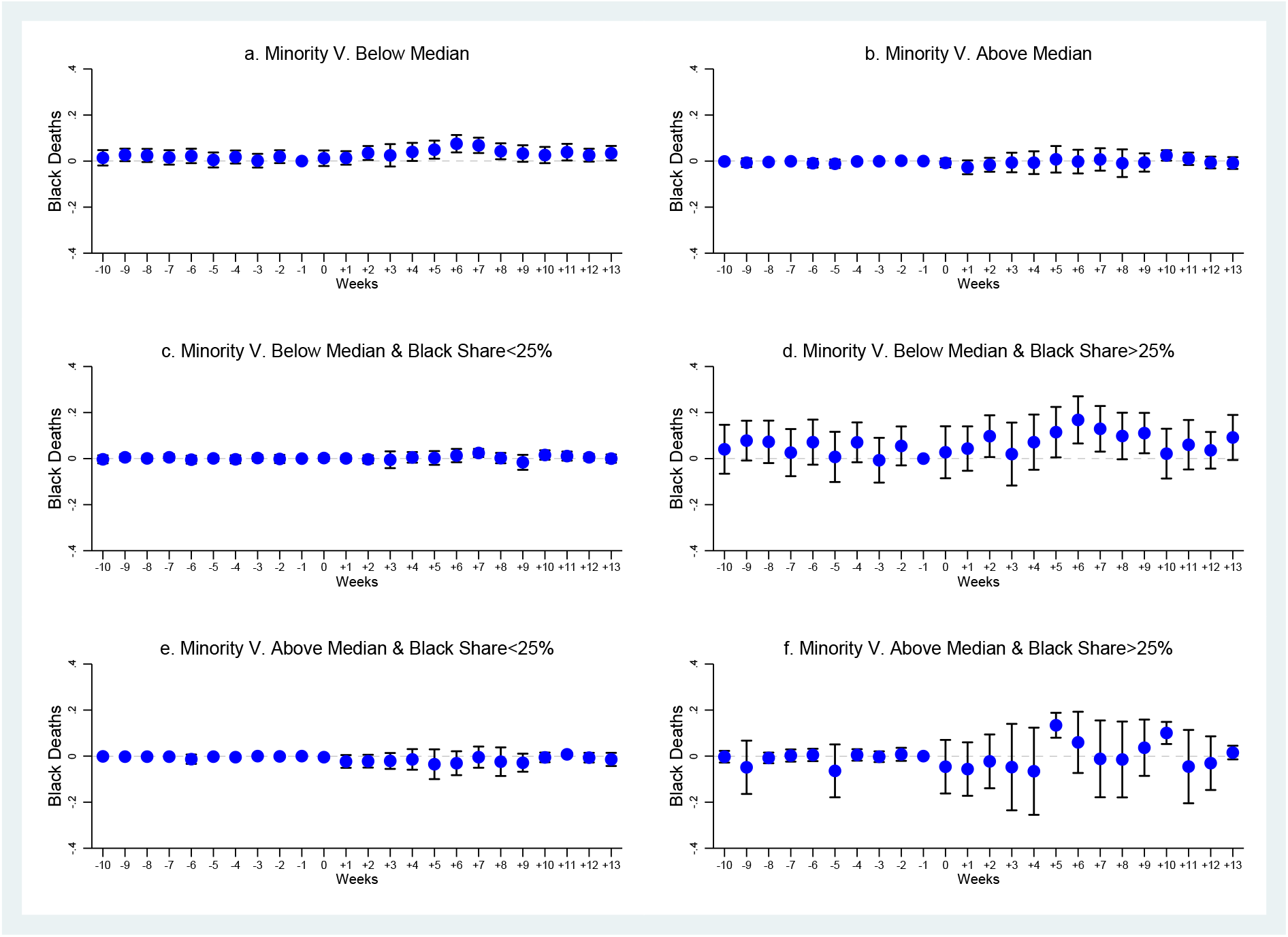
Heterogeneity by Minority Status Vulnerability - Cook County, January 1-June 16, 2020. *Note:* The dependent variable is number of black deaths. The coefficients are least-squares estimates of the *β*_*k*_s over samples of block groups with minority status vulnerability below median (Panel a), above median (Panel b), below median and with black share below 25 percent (Panel c), below median and with black share above 25 percent (Panel d), above median and with black share below 25 percent (Panel e), and above median and with black share above 25 percent (Panel f). Block group and week fixed effects are included. Vertical lines represent 95 percent confidence intervals based on standard errors clustered at at block group level. The omitted period *k* = *−*1, i.e., week 10.

**Figure 11:**
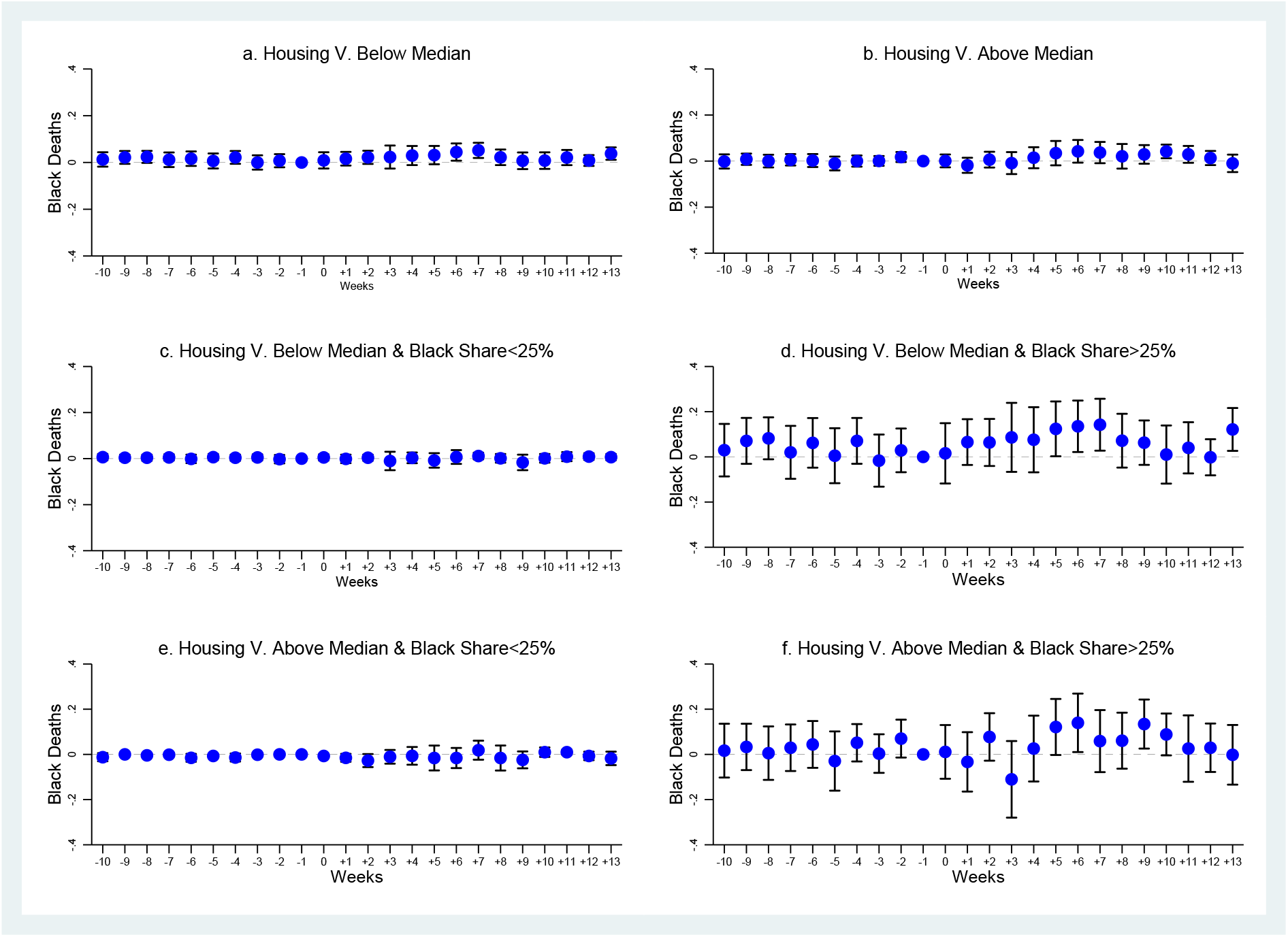
Heterogeneity by Housing/Transportation Vulnerability - Cook County, January 1-June 16, 2020. *Note:* The dependent variable is number of black deaths. The coefficients are least-squares estimates of the *β*_*k*_s over samples of block groups with housing/transportation vulnerability below median (Panel a), above median (Panel b), below median and with black share below 25 percent (Panel c), below median and with black share above 25 percent (Panel d), above median and with black share below 25 percent (Panel e), and above median and with black share above 25 percent (Panel f). Block group and week fixed effects are included. Vertical lines represent 95 percent confidence intervals based on standard errors clustered at at block group level. The omitted period *k* = *−*1, i.e., week 10.

### 8.3 Heterogeneity by nursing home location

Lastly, the high levels of mortality recorded in nursing homes have been stressed both by the media and the medical literature. To explore the relevance of this potential channel, using the information on nursing home location available from Medicare^41^ we generate the minimal centroid distance of a block group from a nursing home and we replicate the heterogeneity analysis focusing on block groups located within 0.009 degrees (i.e., 1 km) from a nursing home. Our aim is to capture the probability that a recorded death has occurred in a nursing home. Figure 12 shows that the treatment effect is much stronger in neighborhoods further away from nursing homes and that this is especially true when the black share is above 25 percent. Thus, we can confidently exclude potential concerns related to the possibility that we could have captured the effect of deaths in nursing homes.

**Figure 12:**
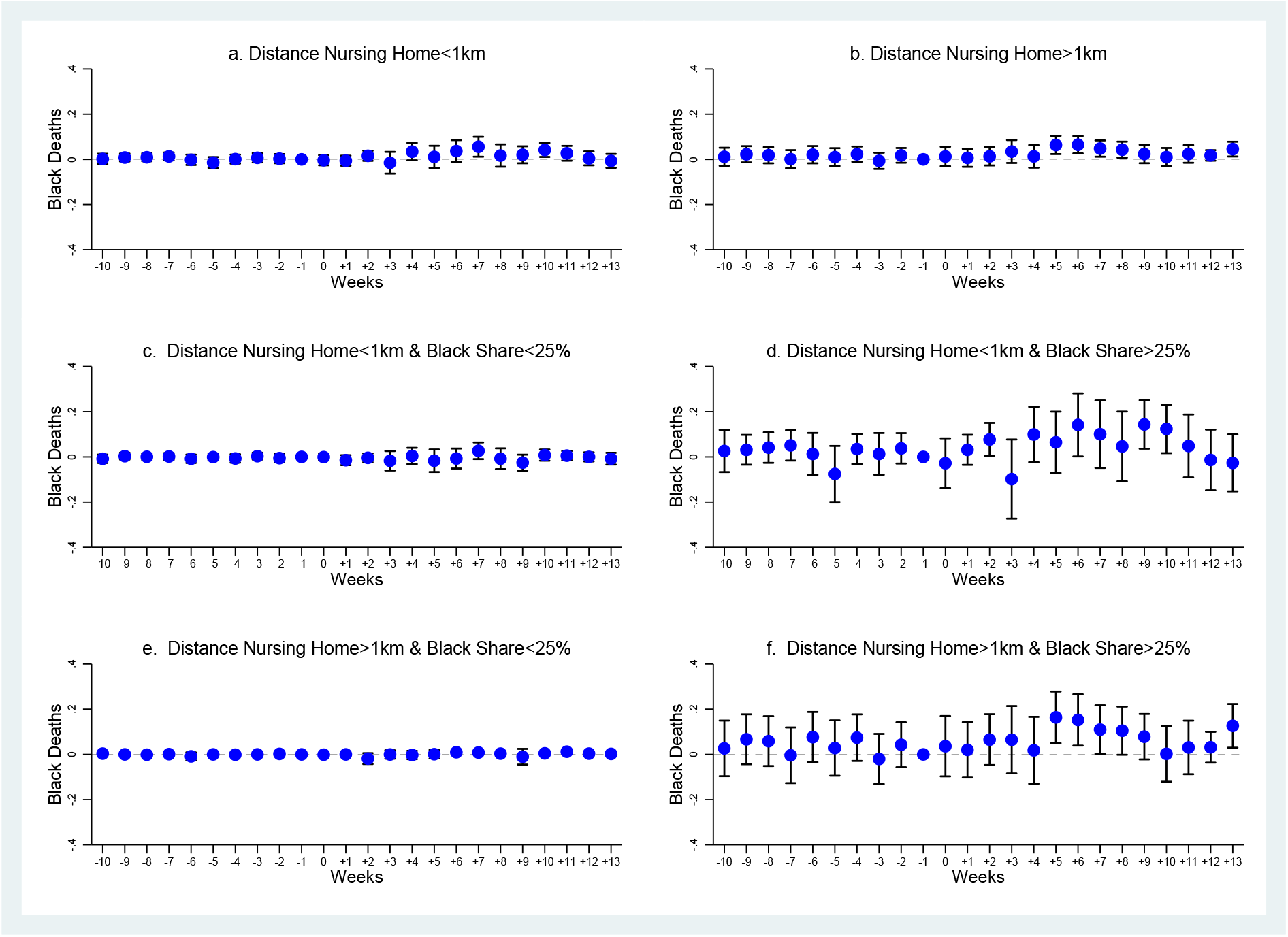
Heterogeneity by Distance from Nursing Home - Cook County, January 1-June 16, 2020. *Note:* The dependent variable is number of black deaths. The coefficients are least-squares estimates of the *β*_*k*_s over samples of block groups with distance from nursing home below 1 km (Panel a), above 1 km (Panel b), below 1 km and with black share below 25 percent (Panel c), below 1 km and with black share above 25 percent (Panel d), above median and with black share below 25 percent (Panel e), and above median and with black share above 25 percent (Panel f). Block group and week fixed effects are included. Vertical lines represent 95 percent confidence intervals based on standard errors clustered at at block group level. The omitted period *k* = *−*1, i.e., week 10.

## 9 The Hispanic population

While our main focus so far has been on how the African American population has been hit by COVID-19, the Hispanic population has also been the subject of concern, both in the media and the medical literature.^42^ Therefore, in this section, we extend the previous analysis of COVID-19 outcomes to the white Hispanic population of Cook County.^43^

The history of Latino immigration to Cook County starts at least as early as in the period 1916-1928, when a steady and large flow of Mexicans moved to Chicago to find work in the railroad and steel industries. Another wave took place in the period 1942-1964. Their settlement pattern was similar to that of blacks and they were also historically affected by segregation and redlining (Betancur, 1996). In fact Hoyt (1933) placed Mexicans last, after blacks, in his ranking of the influence of ethnic groups on property values.

Figure A21 is a replica of Figure 2 that also reports Hispanic COVID-19 deaths, as well as blacks and the remaining groups combined, from March 16 to June 16. Even though the number of Hispanic deaths does increase in the initial weeks, it stays below the number of black deaths. Figure A22 is a replica of Figure 5 that plots the Hispanic overall mortality rate from January 1 to June 16. Strikingly, before the epidemic, the mortality rate for Hispanic is smaller than that of blacks and even of other groups combined. It does increase with the COVID-19 outbreak, but remains relatively contained if compared to other groups.

The fact that Hispanics exhibit lower levels of mortality than the rest of the population is actually a well-known fact, that has been referred to as the “Latino paradox”, as Hispanics tend to display relatively favorable health outcomes despite their low socioeconomic status (Markides and Coreil, 1986; Abraido-Lanza et al., 1999).

Nevertheless, when in Figure A23 we replicate Figure 6 with the baseline event study, including results for Hispanics, we find that they are also affected by the treatment, even though the effect kicks in later, and lasts longer, if compared to blacks. This confirms that HOLC practices carry long-term implications, in terms of the induced resilience to the epidemic shock, also for Hispanics.

## 10 Falsification tests

### 10.1 From 2017 to 2019

There are still additional potential threats to identification. To test the extent of such threats, in this section we will carry out two sorts of falsification tests.

First, it might be the case that the treatment captures endemic annual trends in mortality that would have anyway occurred and that have nothing to do with the COVID-19 epidemic. To test the extent of such threat to identification, we carry out falsification tests using the number of deaths in the corresponding time frame for the three years prior to the epidemic, that is, from January 1 and June 16 in 2017, 2018, and 2019. The goal is to gauge the possibility that the treatment is capturing annual trends in mortality related to the diffusion of other diseases (e.g., the flu).^44^ By replicating the analysis over the previous years, we shall be able to ascertain that we are not merely capturing a yearly trend having nothing to do with the COVID-19 epidemic.

Figure 13 shows that there is no effect of the treatment on black or other deaths in either of the three previous years over the same time frame, which confirms that the effect we found is attributable to the 2020 COVID-19 epidemic.

**Figure 13:**
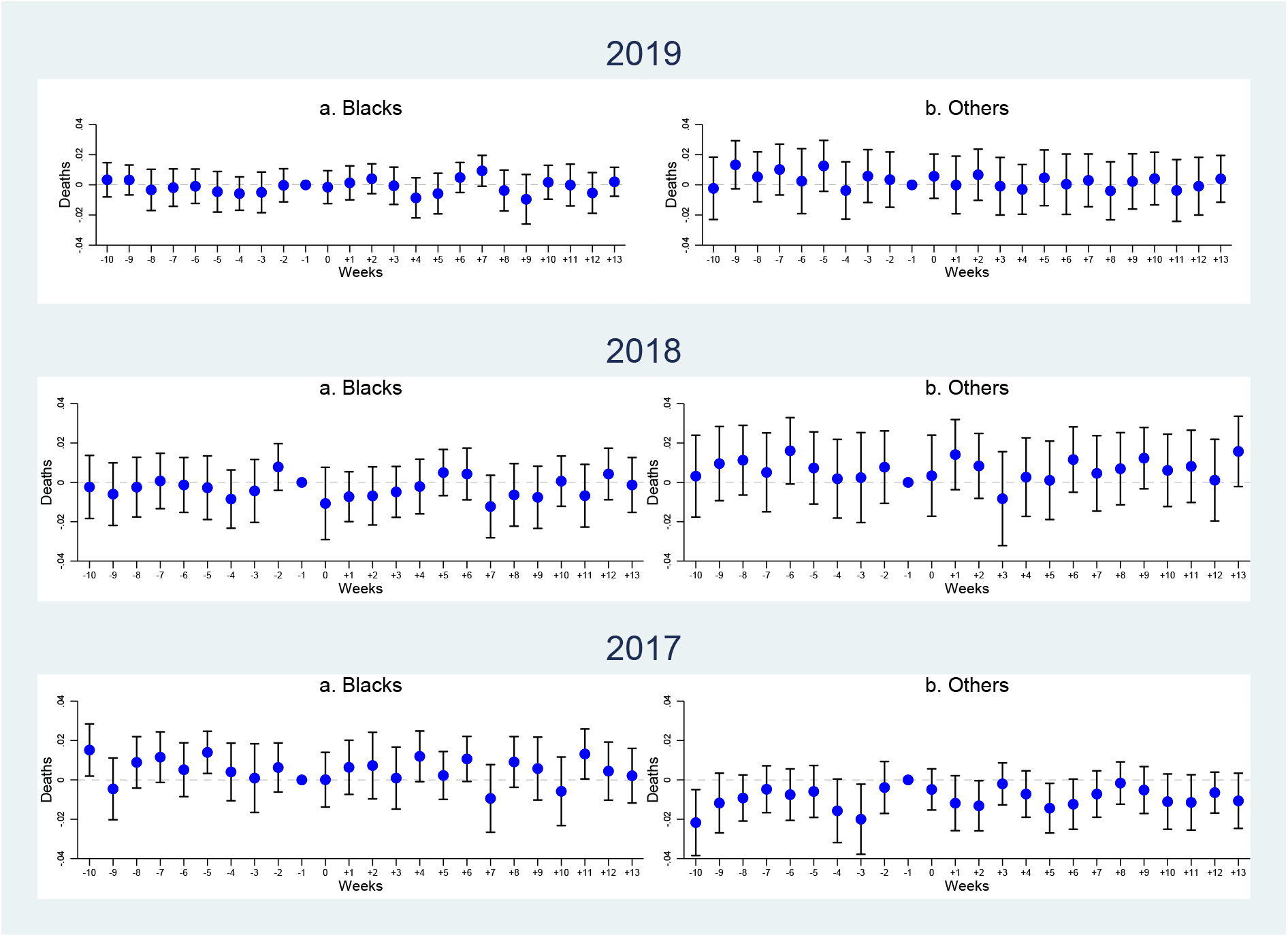
Dynamic Effect of the Treatment on Deaths, by Race - Cook County, January 1-June 16, 2019, 2018, and 2017. *Note:* The dependent variable is number of deaths, of blacks (Panels a) and of other groups (Panels b), in the period from January 1 to June 16, in 2019 (top panels), 2018 (middle panels), and 2017 (bottom panels). The coefficients are least-squares estimates of the *β*_*k*_s. Block group and week fixed effects are included. Vertical lines represent 95 percent confidence intervals based on standard errors clustered at at block group level. The omitted period *k* = *−*1, i.e., week 10.

### 10.2 The Spanish flu

A further threat to identification is due to the fact the transmission rate of diseases (not mortality) may have always been larger in redlined neighborhoods, so that the 1930s rankings may have merely formalized existing conditions. In other words, it may be the case that low-graded neighborhoods may have already been subject to higher transmission of viral diseases, even prior to redlining. To make sure that our results are not affected by this kind of threat, we focus on Spanish flu mortality.

By 1918, Chicago had already experienced the first wave of the Great Migration, with the black population rising from about 34,000 to 92,000 between 1910 and 1920, and a parallel increase of racial segregation (Tuttle, 1970). Chicago was badly hit by the Spanish flu, but blacks were actually hit less harshly than whites, as in the rest of the country, a fact that is still largely unexplained (Gamble, 2010; Okland and Mamelund, 2019). From our perspective, the fact that blacks were already segregated, by and large, in the same areas of the city, while at the same time they were somewhat protected from the disease, suggests that those areas were not per se more unhealthy.

In order to perform a falsification test, we use census tract level data on Spanish flu mortality in Chicago in 1918 to test whether the transmission of epidemics in low-graded areas was already higher before they were assessed by the HOLC. Data are provided by the Infectious Disease Dynamics Group.^45^ The dataset contains census tract location and week of epidemic of 8,031 influenza and pneumonia deaths as well as sociodemographic data (including population density, the illiteracy rate, the home ownership rate, and the unemployment rate) for 496 census tracts within the City of Chicago. However, race-disaggregated information is not reported.^46^

Table 2 reports four variants of a model where the number of Spanish flu deaths in 1918 is the dependent variable. In Model 1, where we only control for majority C or D neighborhoods and week fixed effect, we find a marginally significant effect of the control variable on deaths. However, when in the following models we add (the log of) population density, illiteracy, home ownership, and unemployment, no residual influence remains. Thus, reassuringly, we find no evidence that the HOLC areas that we found to be associated with higher COVID-19 mortality were subject to higher infections rates prior to the implementation of the redlining policies of the 1930s.

**Table 2:** Spanish Flu - Chicago, 1918.

|  | Deaths From Spanish Flu |  |  |  |
| --- | --- | --- | --- | --- |
|  | (1) | (2) | (3) | (4) |
| Majority C & D | 0.3020*<br>(0.1588) | 0.1039<br>(0.1525) | 0.1257<br>(0.1498) | 0.1070<br>(0.1479) |
| Population Density (log) | Yes | Yes | Yes | Yes |
| Illiteracy Rate | No | Yes | Yes | Yes |
| Home Ownership Rate | No | No | Yes | Yes |
| Unemployment Rate | No | No | No | Yes |
| Week FE | Yes | Yes | Yes | Yes |
| Adj.R-squared | 0.252 | 0.281 | 0.284 | 0.287 |
| Observations | 3416 | 3416 | 3416 | 3416 |

*Note:* The dependent variable is the number of deaths from Spanish flu in Chicago in 1918. Robust standard errors clustered at a census tract level in parentheses: \*\*\* $p < 0.01$ , \*\* $p < 0.05$ , \* $p < 0.1$ .

## 11 Conclusion

Not only the United States is registering the worldwide highest number of fatalities from the COVID-19 pandemic but, within the country, the death toll on African Americans has been disproportionately large. Up to now, however, lack of race-disaggregated data has prevented a rigorous assessment of this phenomenon and of its determinants.

Using so far unexploited individual and georeferenced death data collected by the Cook County Medical Examiner, we provide first evidence that race does affect COVID-19 outcomes. The data confirm that in Cook County blacks are overrepresented in terms of COVID-19 related deaths since—cumulatively in the period that goes from the outbreak of the epidemic on March 16, 2020 until June 16, 2020—they constitute 35 percent of the dead, which implies that they have been dying at a rate 1.3 times higher than their population share.

Furthermore, by combining the spatial distribution of mortality with the redlining maps for the Chicago area, we obtain a block group level panel dataset of weekly deaths from all causes, over the period January 1, 2020-June 16, 2020, to which we apply an event study design, where the treated neighborhoods are those that were historically either yellow or redlined and treatment initiation coincides with the outbreak of the COVID-19 epidemic. We show that, while no pre-treatment differences are detected, after the outbreak of the epidemic on March 16, 2020 historically low-graded neighborhoods display a sharper increase in mortality, which is driven by blacks. Thus, we uncover a persistence influence of the racial segregation induced by the discriminatory lending practices introduced in the 1930s.

We also establish that this influence runs by way of an asymmetric effect of the epidemic shock, which is in turn channeled through a diminished resilience of the black population to the shock represented by the COVID-19 outbreak. Far from being determined by genetic and biological factors, such vulnerability can be linked to socioeconomic status and household composition, as the likely channels through which the legacy of the past manifests itself.

To conclude, one of the stylized facts emerging from this paper, and that deserves further attention, is that not only blacks are disproportionately hit by COVID-19, but also that they started to succumb to it earlier than other groups. Several explanations may be behind this phenomenon. On the one hand, it is possible that blacks become infected as much as the rest of the population, but they experience a faster progression through the stages of the disease, because of pre-existing medical conditions and/or access to health care. It may also be the case that blacks were more exposed from the beginning of the outbreak, because of their occupations and living conditions, so that once stay-at-home orders where issued they benefitted from them more thoroughly. The age and gender composition of each racial group also may also play a role.^47^ The fact remains that the evolution of the epidemiological curve reveals for the US an extraordinary degree of racial and ethnic segregation, with different groups displaying profoundly distinct patterns even in the timing of their exposure to the epidemic.

## Data Availability

Data are publicly available.

## APPENDIX

**Figure A1:**
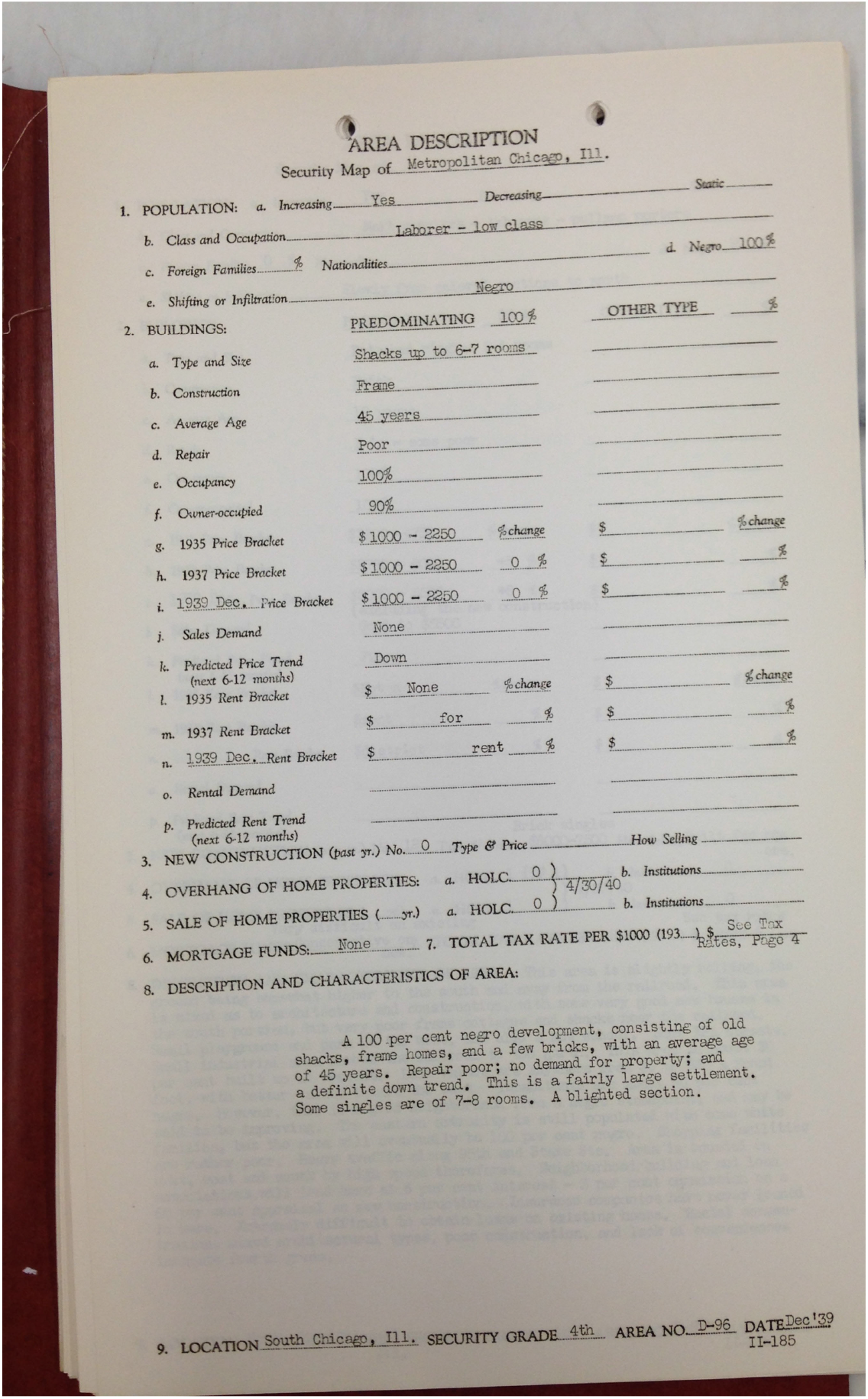
HOLC Area Description File for Area D96, Chicago Metropolitan Area, 1939.

**Figure A2:**
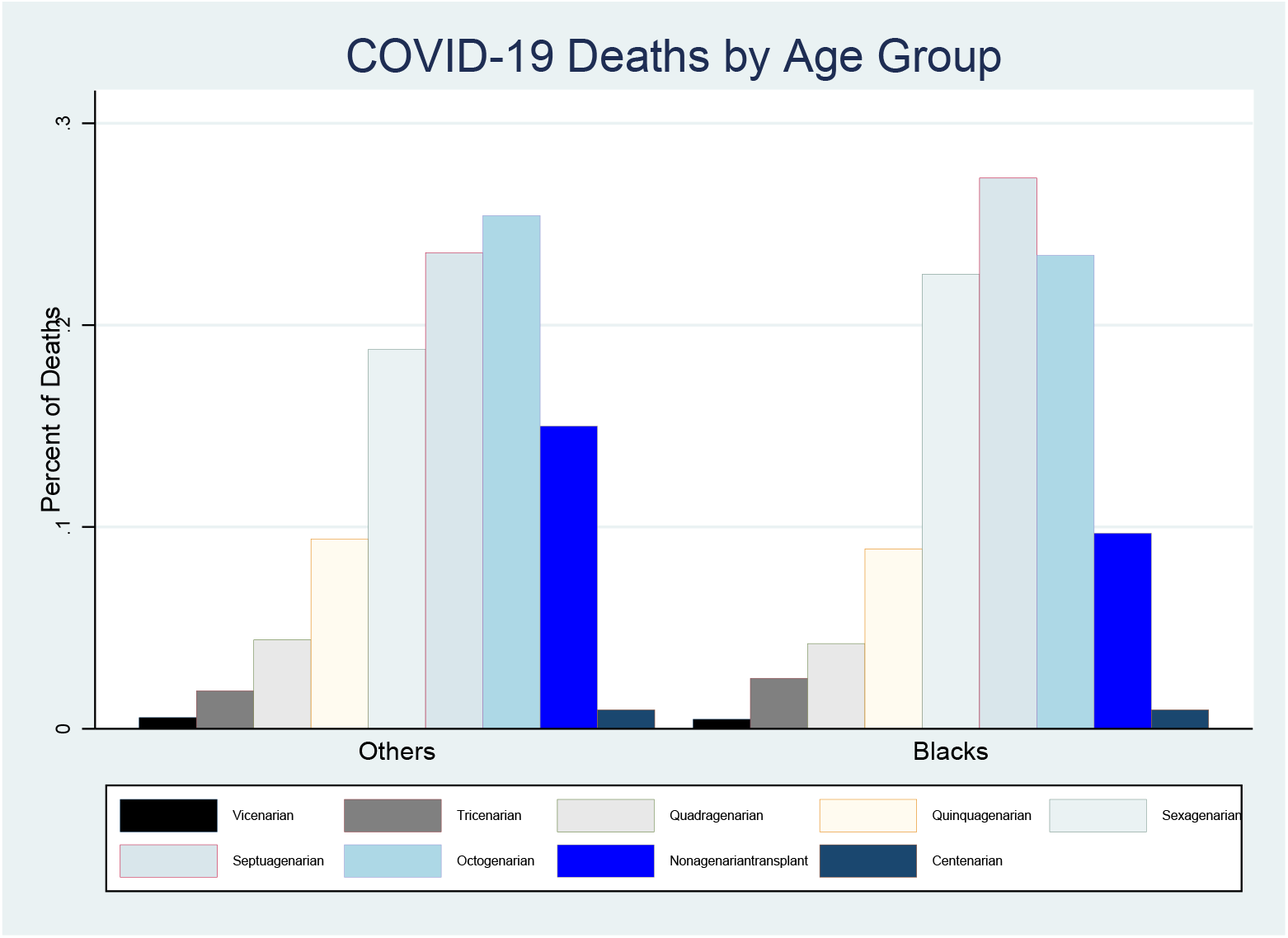
COVID-19 Deaths, by Age Group - Cook County, March 16-June 16, 2020.

**Figure A3:**
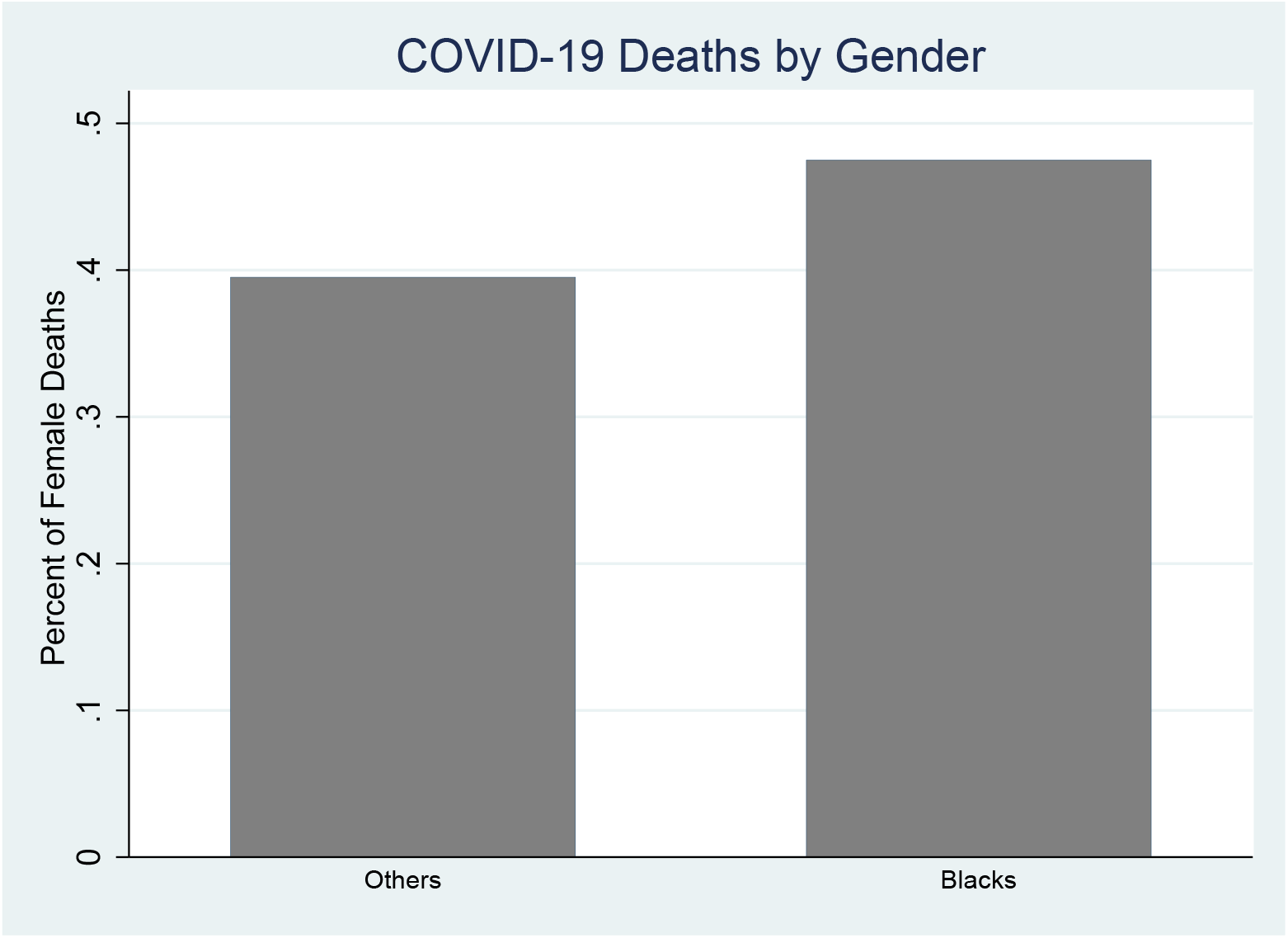
COVID-19 Deaths, by Gender - Cook County, March 16-June 16, 2020.

**Figure A4:**
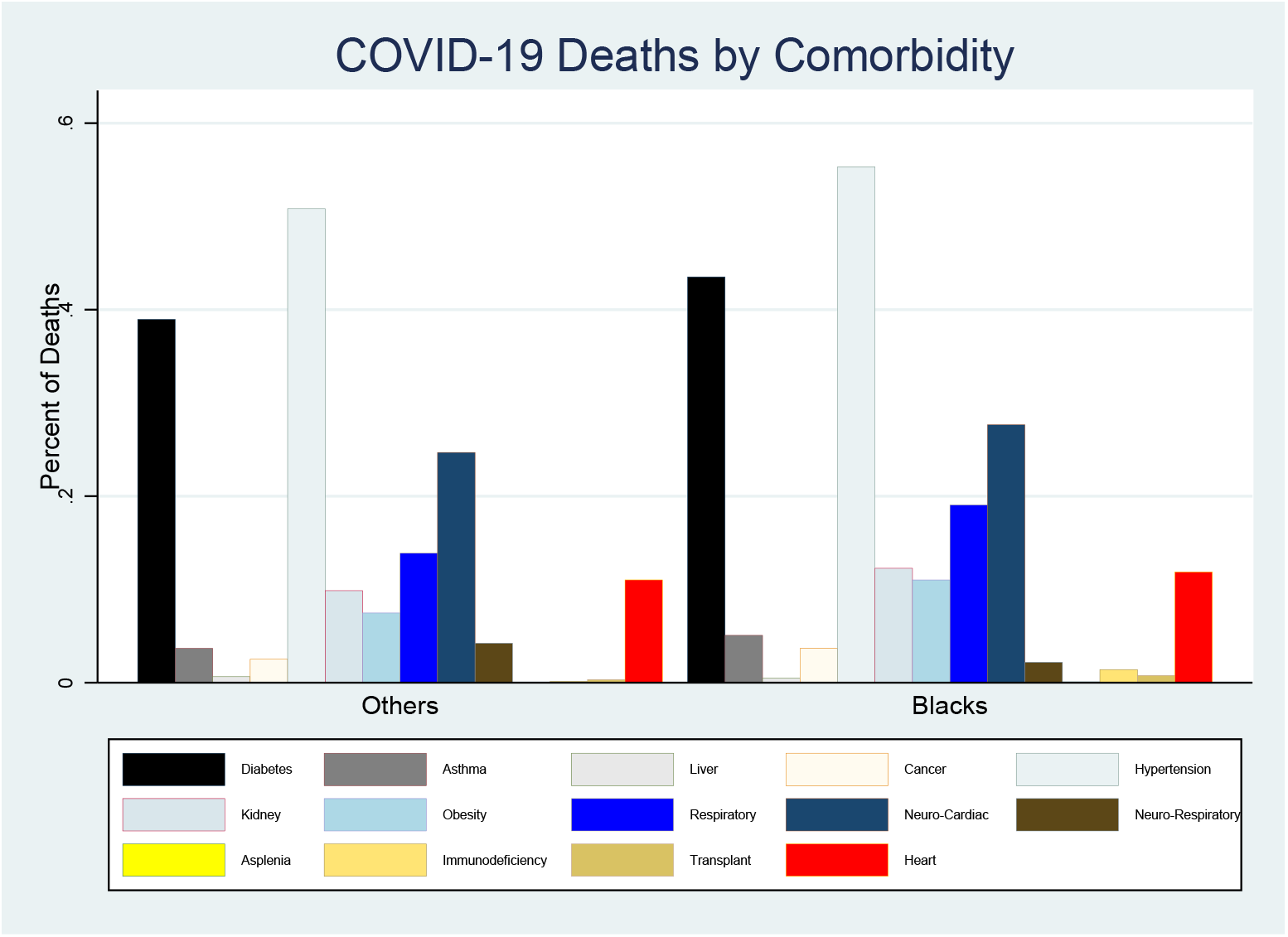
COVID-19 Deaths, by Comorbidity - Cook County, March 16-June 16, 2020.

**Figure A5:**
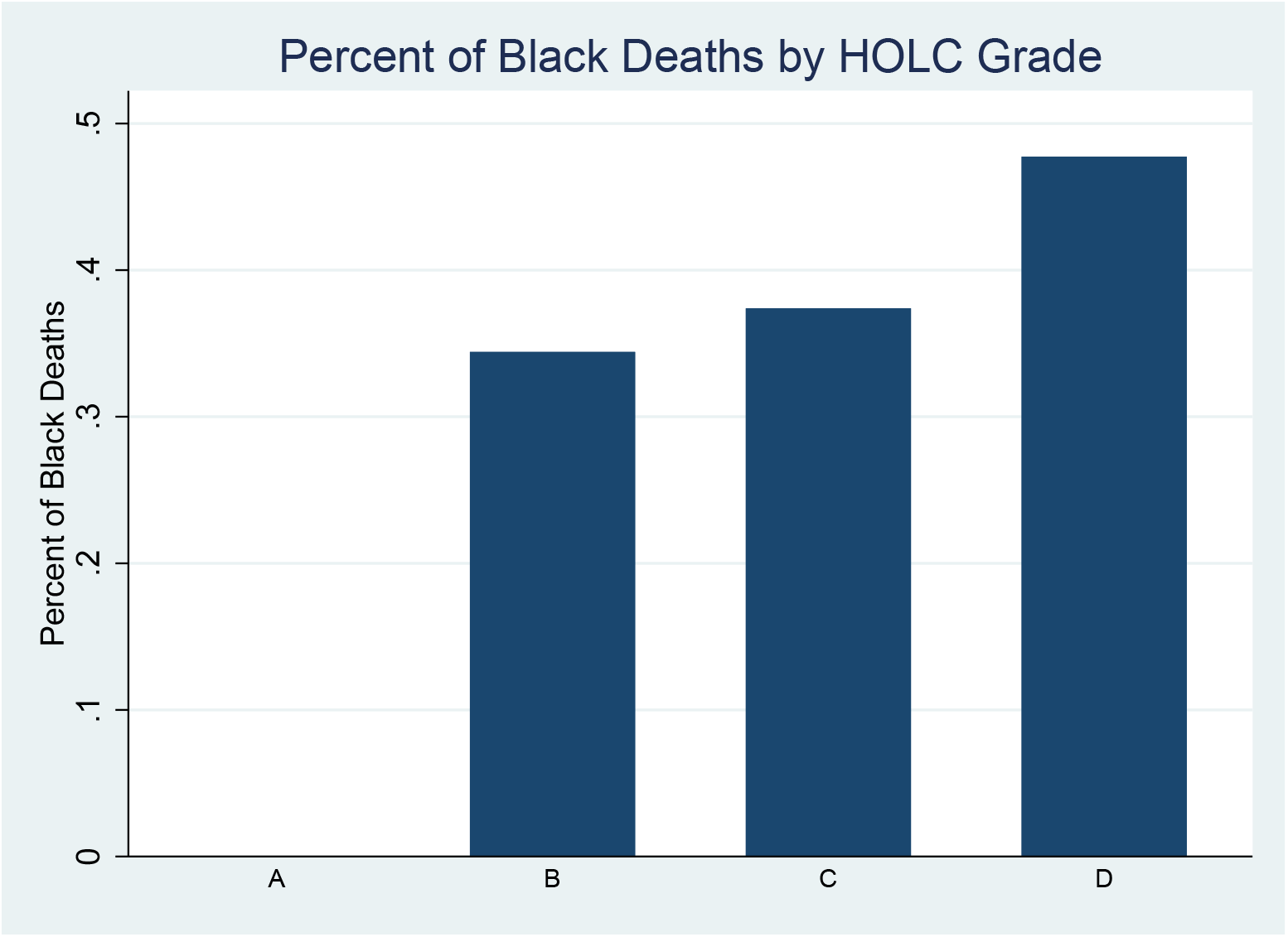
Share of Black COVID-19 Deaths, by HOLC Grade - Cook County, March 16-June 16, 2020.

**Figure A6:**
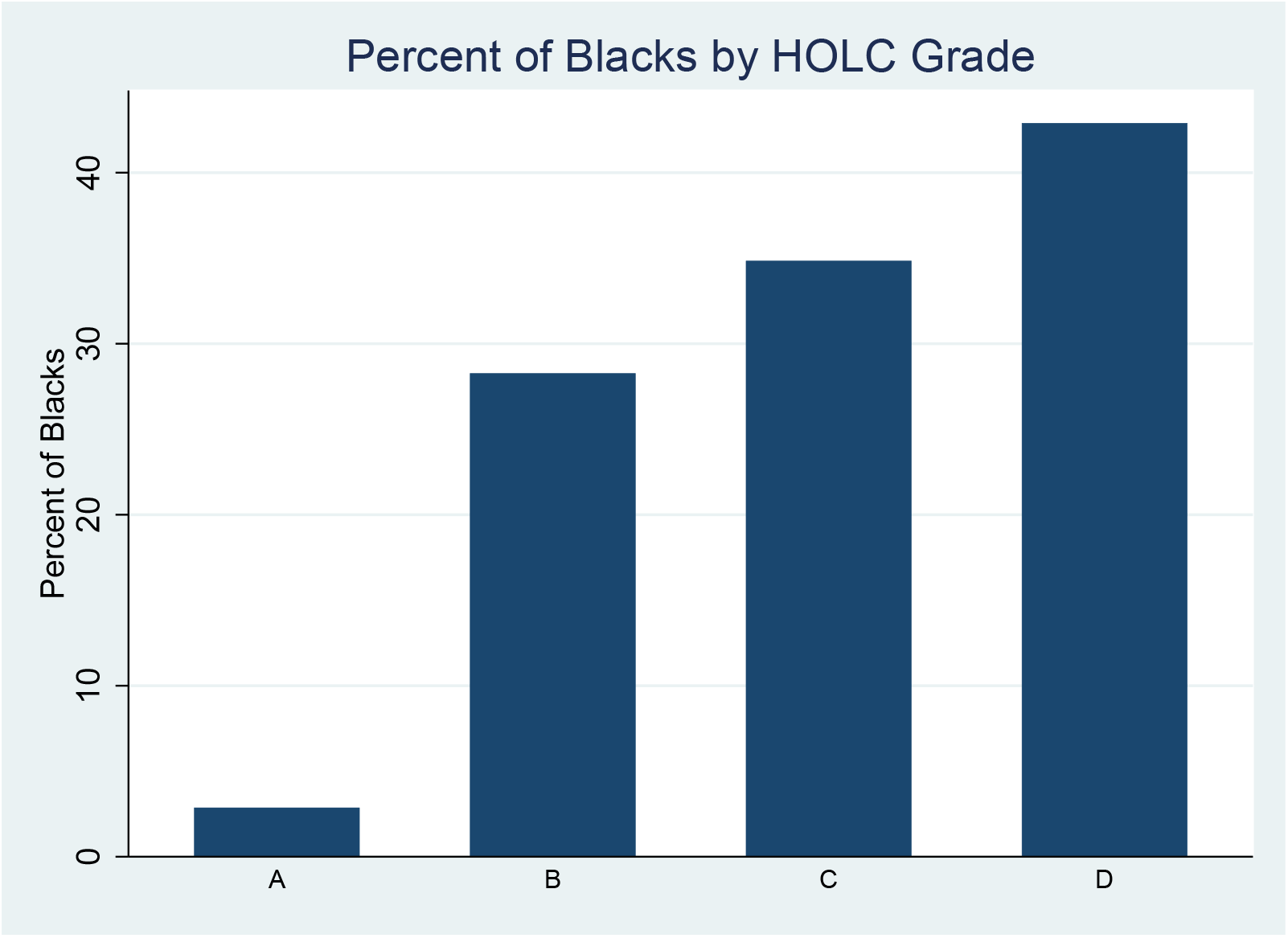
Black Population Share, by HOLC Grade - Cook County, 2010 Census.

**Figure A7:**
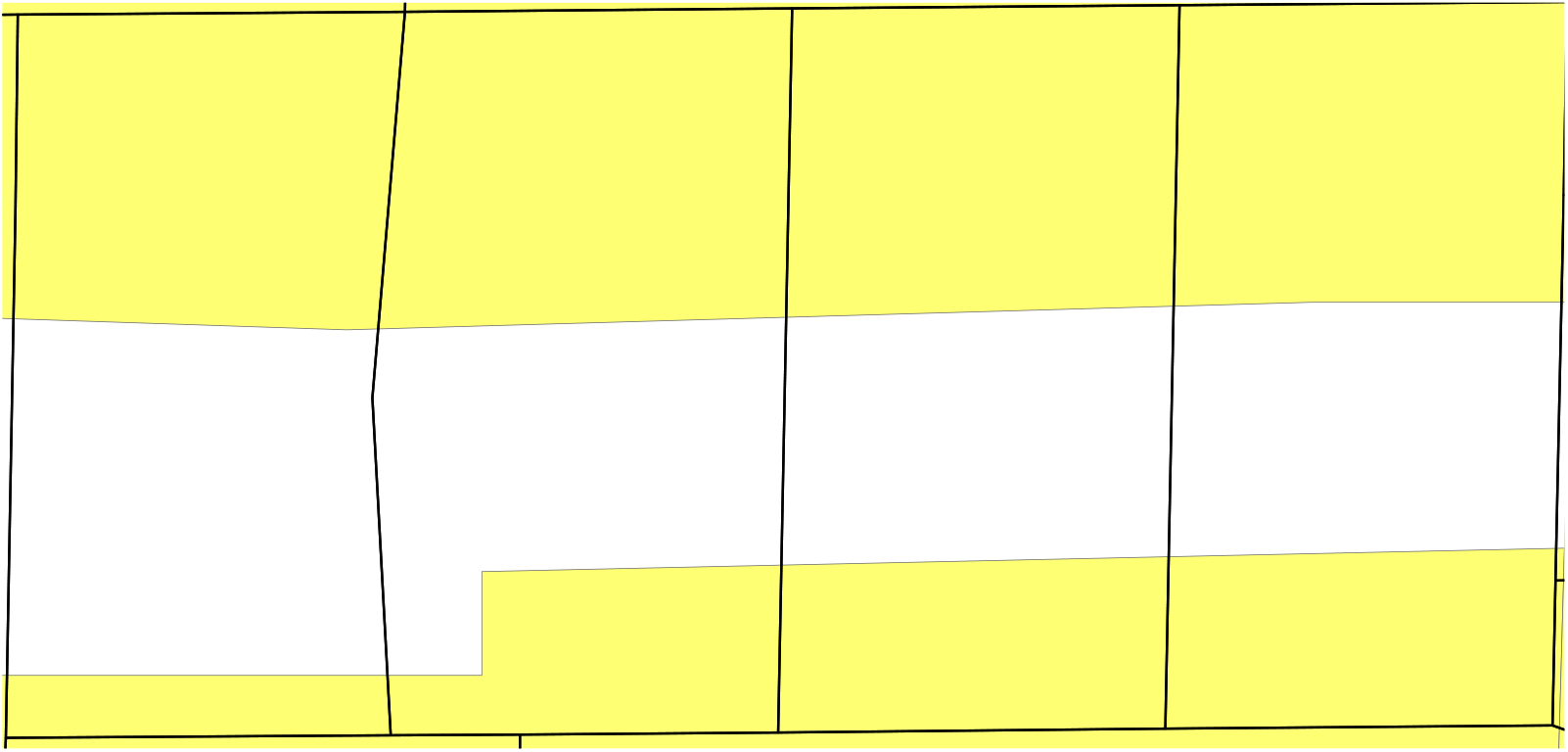
HOLC Neighborhoods by Block Group. *Note:* The figure reports an example of four block groups, each partitioned into three neighborhoods, two of which belonging to two different HOLC areas, both graded C (in yellow) and one ungraded (in white).

**Figure A8:**
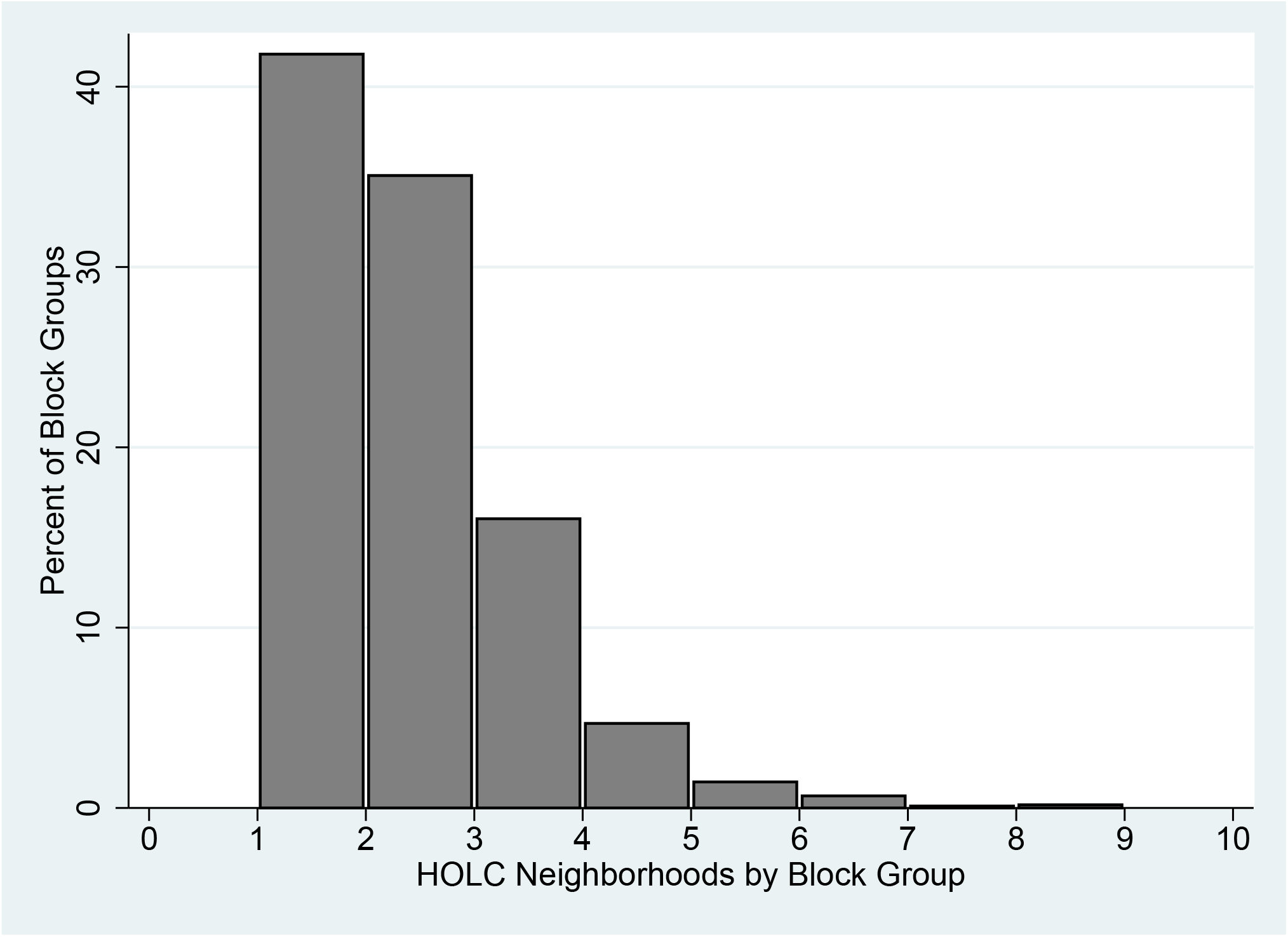
Distribution of HOLC Neighborhoods by Block Group. *Note:* The Figure Ahows that over 40 percent of the block groups in the sample include only one HOLC neighborhood (potentially with the same HOLC grade), while around 35 percent include two, and so on, with a small percentage including up to ten.

**Figure A9:**
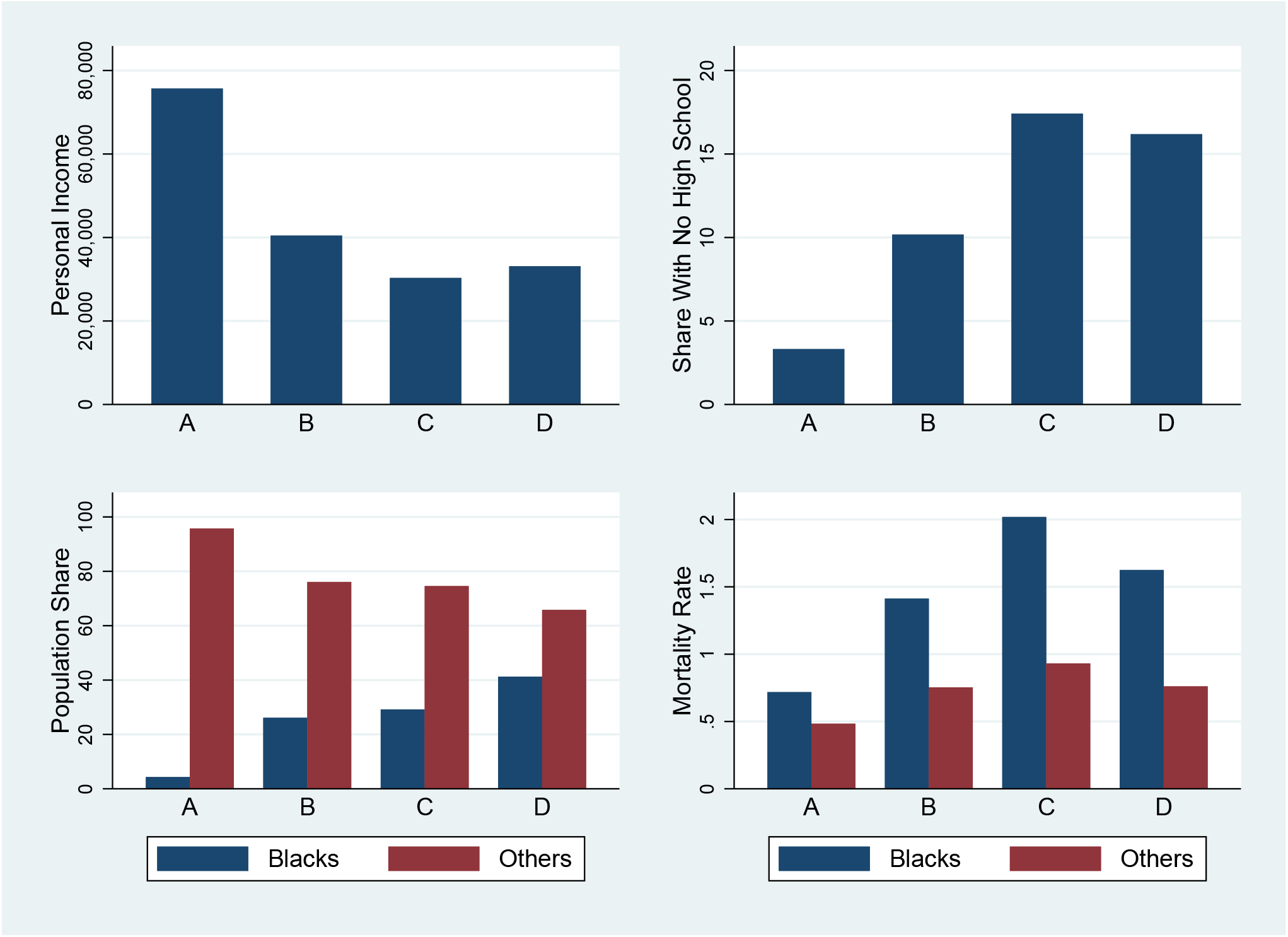
Socioeconomic and Demographic Characteristics by HOLC Grade. *Note:* Personal income, share with no high school, population share by race, and mortality rate by race, by HOLC area.

**Figure A10:**
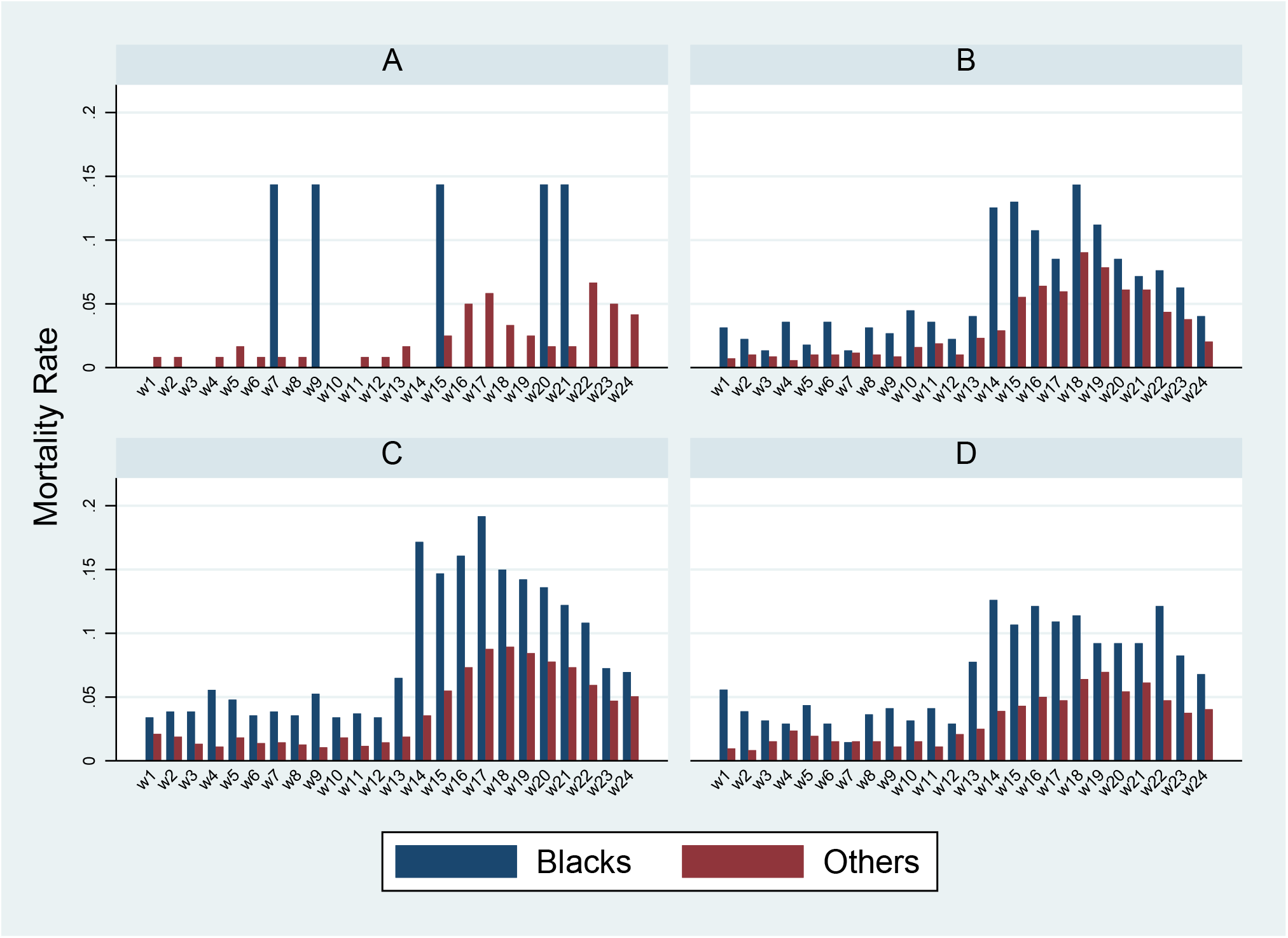
Mortality Rate, by Race, Week, and HOLC Grade - Cook County, March 16-June 16, 2020.

**Figure A11:**
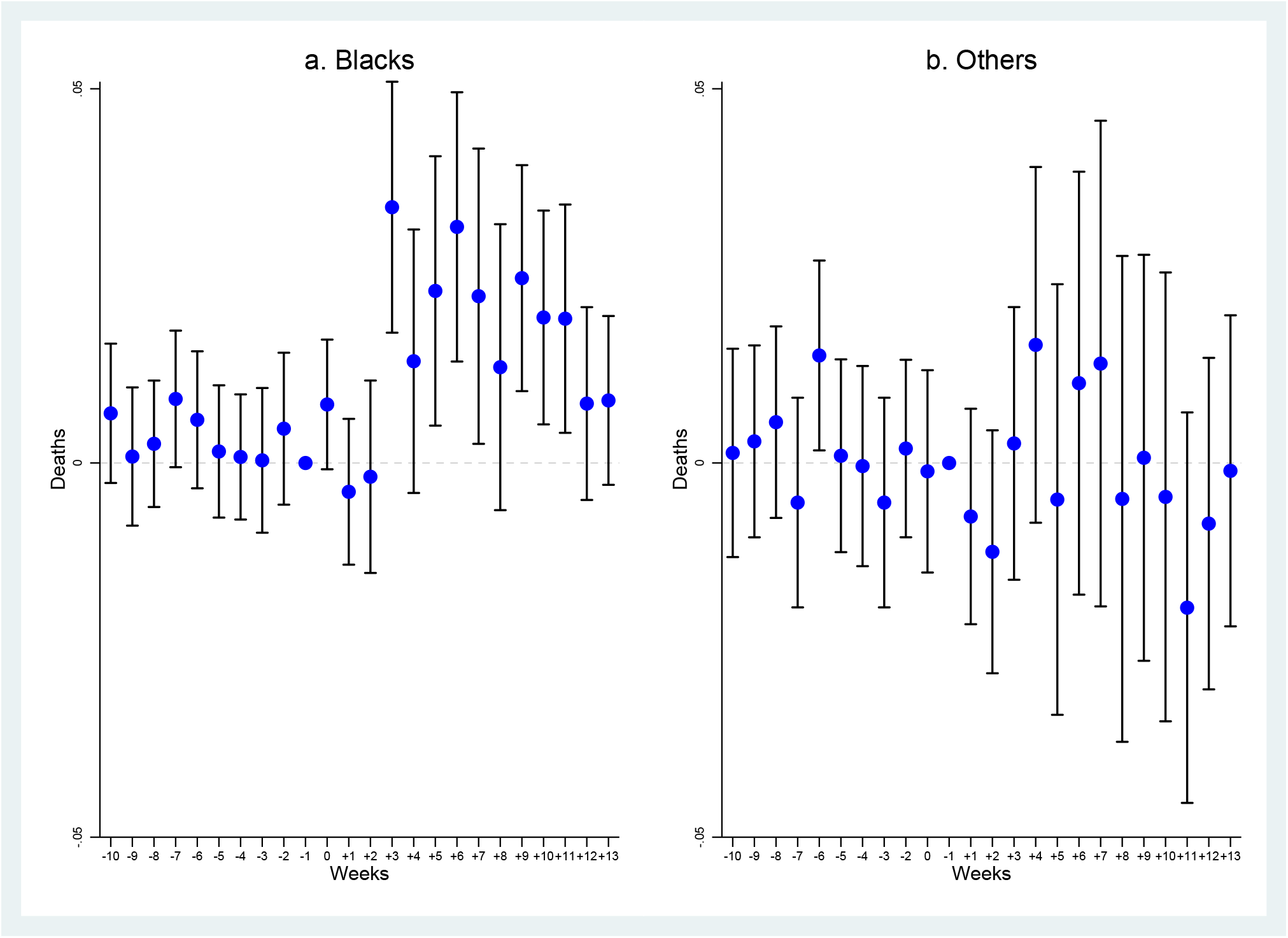
Including Block Groups Ungraded by the HOLCD - Cook County, January 1-June 16, 2020. *Note:* The dependent variable is number of deaths, of blacks (Panel a) and of other groups (Panel b). The sample includes block groups that were not graded by the HOLC. The coefficients are least-squares estimates of the *β*_*k*_s in regressions sequentially including additional interactions between week dummies and the variables indicated for each panel. Block group and week fixed effects are included. Vertical lines represent 95 percent confidence intervals based on standard errors clustered at at block group level. The omitted period *k* = *−*1, i.e., week 10.

**Figure A12:**
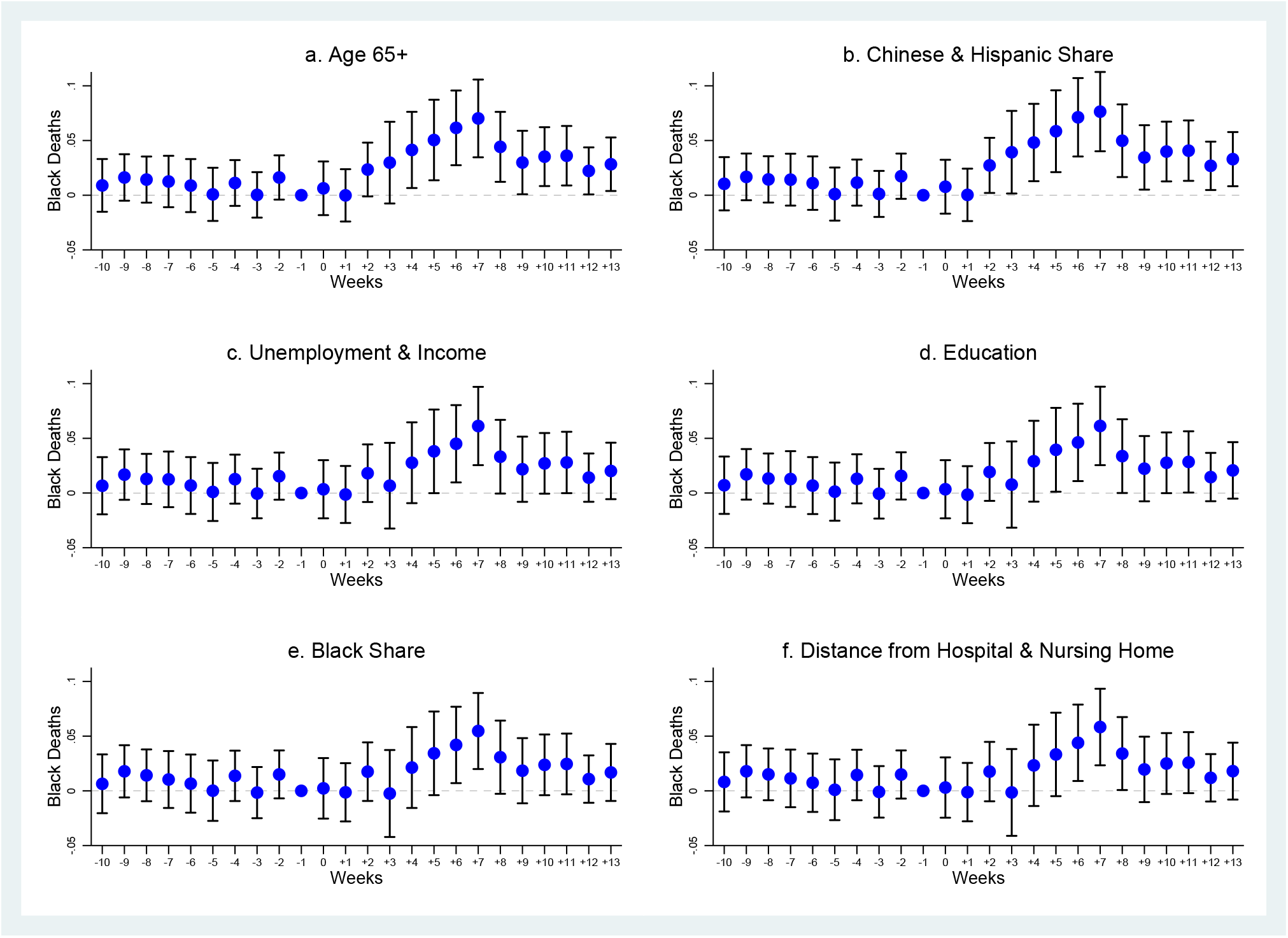
Dummy Death as Alternative Dependent Variable - Cook County, January 1-June 16, 2020. *Note:* The dependent variable is a dummy variable taking value one if a death, of blacks (Panel a) and of other groups (Panel b), occurred in a block group-week, and zero otherwise. The coefficients are least-squares estimates of the *β*_*k*_s in regressions sequentially including additional interactions between week dummies and the variables indicated for each panel. Block group and week fixed effects are included. Vertical lines represent 95 percent confidence intervals based on standard errors clustered at at block group level. The omitted period *k* = *−*1, i.e., week 10.

**Figure A13:**
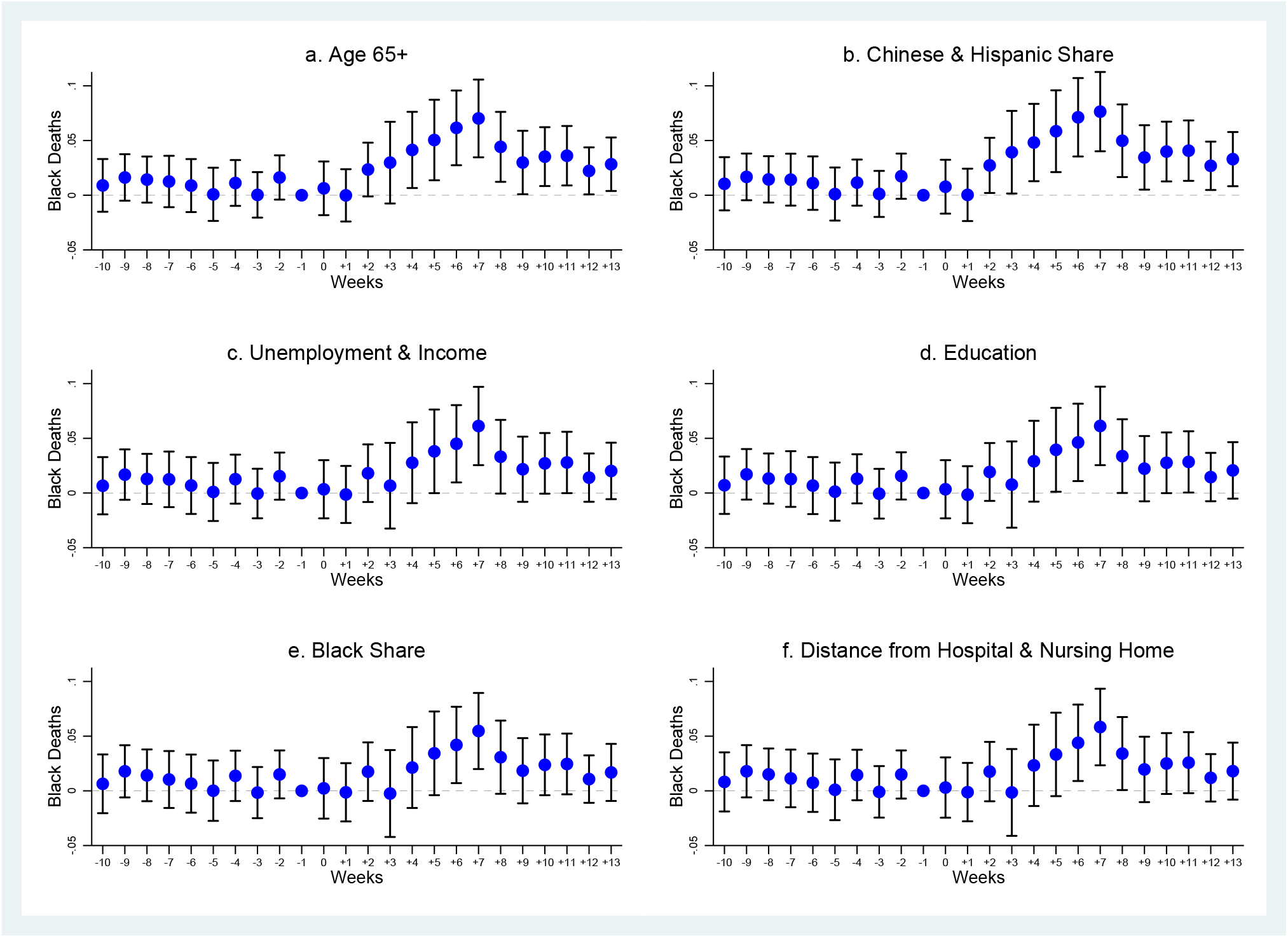
Allowing for Differential Trends - Cook County, January 1-June 16, 2020. *Note:* The dependent variable is number of black deaths. The coefficients are least-squares estimates of the *β*_*k*_s in regressions sequentially including additional interactions between week dummies and the variables indicated for each panel. Block group and week fixed effects are included. Vertical lines represent 95 percent confidence intervals based on standard errors clustered at at block group level. The omitted period *k* = *−*1, i.e., week 10.

**Figure A14:**
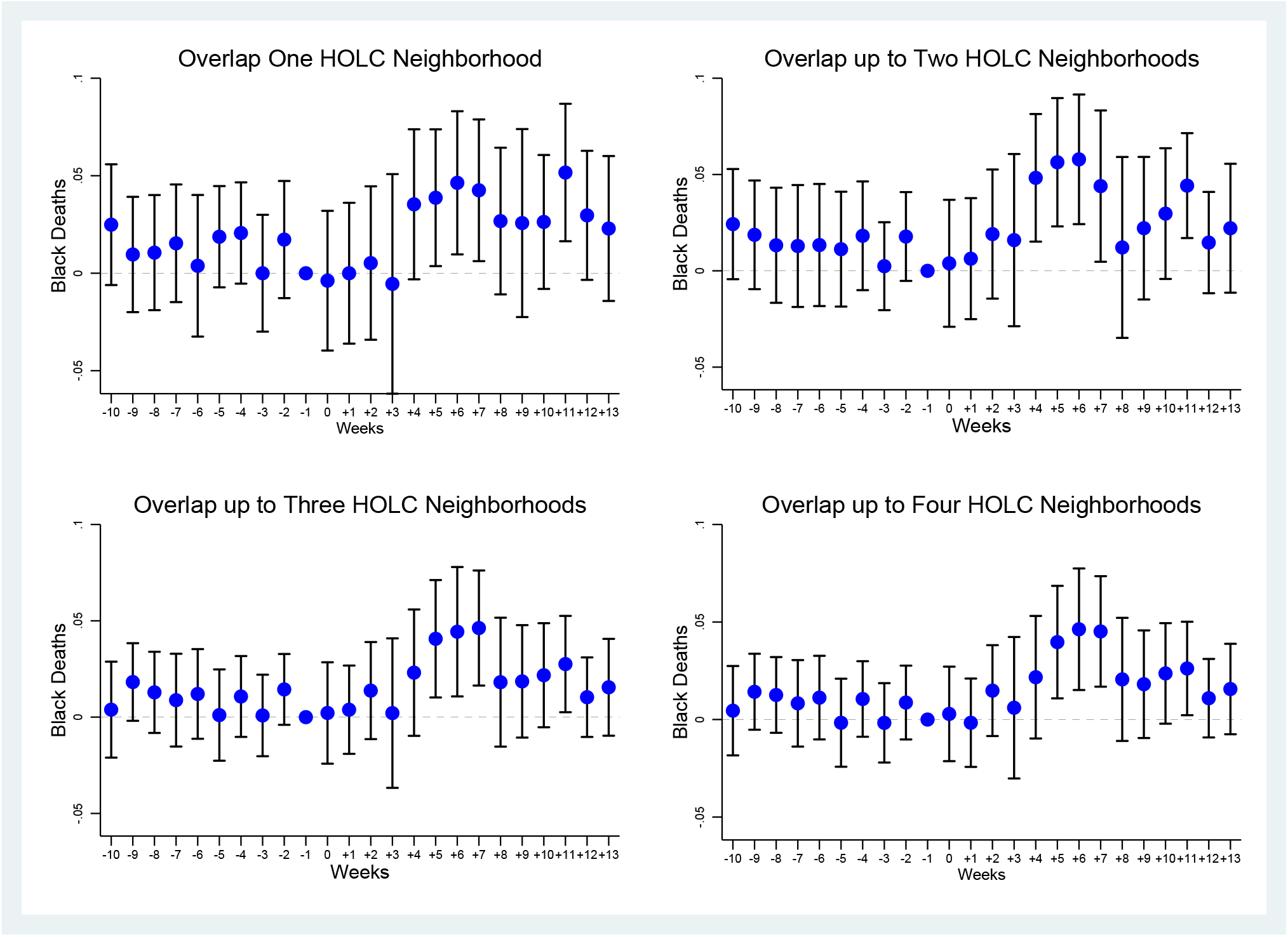
Robustness to Aggregation - Cook County, January 1-June 16, 2020. *Note:* The dependent variable is number of black deaths. The coefficients are least-squares estimates of the *β*_*k*_s over samples of block groups including one, up to two, up to three, and up to four HOLC neighborhoods. Block group and week fixed effects are included. Vertical lines represent 95 percent confidence intervals based on standard errors clustered at at block group level. The omitted period *k* = *−*1, i.e., week 10.

**Figure A15:**
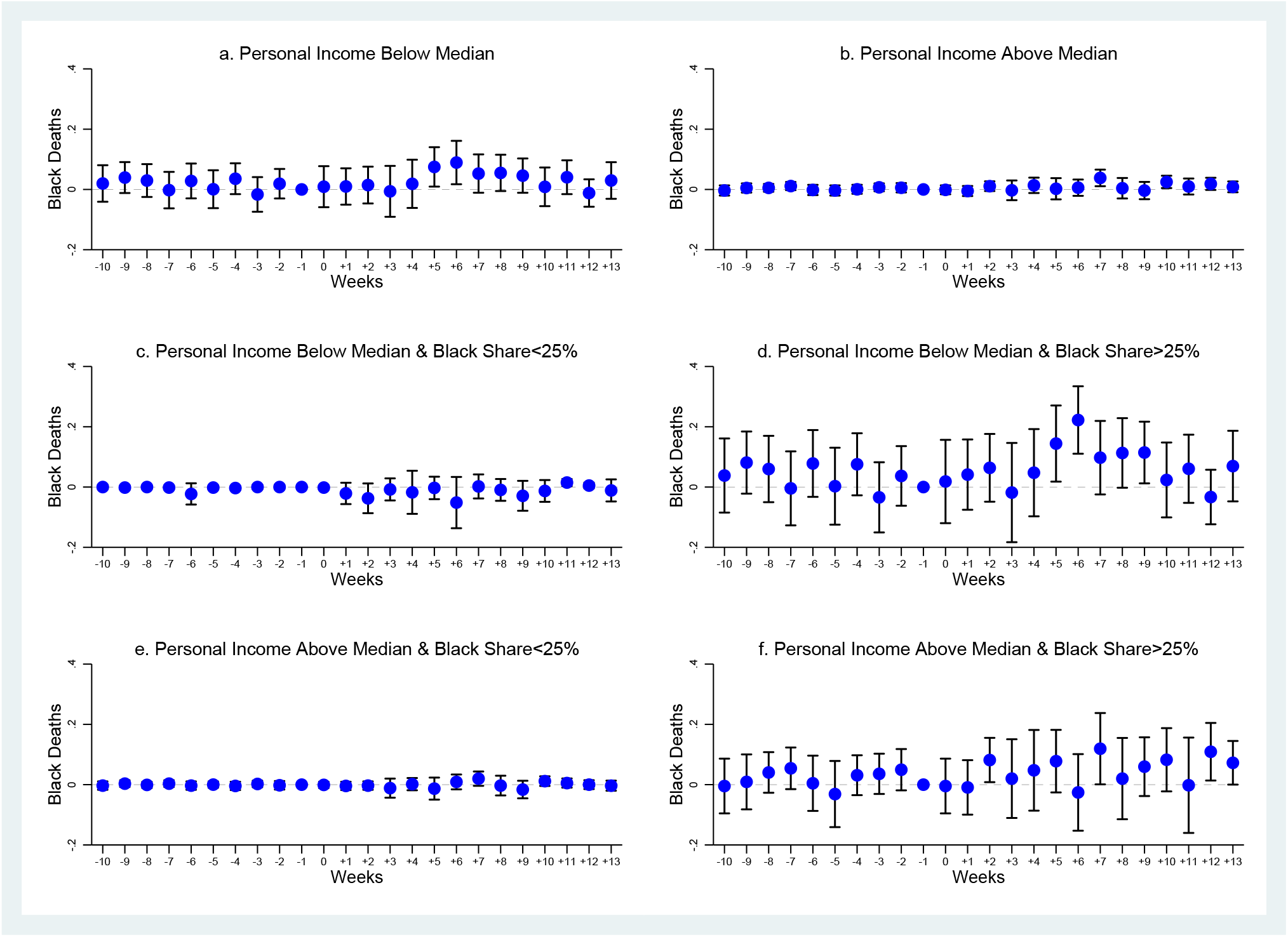
Heterogeneity by Income - Cook County, January 1-June 16, 2020. *Note:* The dependent variable is number of black deaths. The coefficients are least-squares estimates of the *β*_*k*_s over samples of block groups with personal income below median (Panel a), above median (Panel b), below median and with black share below 25 percent (Panel c), below median and with black share above 25 percent (Panel d), above median and with black share below 25 percent (Panel e), and above median and with black share above 25 percent (Panel f). Block group and week fixed effects are included. Vertical lines represent 95 percent confidence intervals based on standard errors clustered at at block group level. The omitted period *k* = *−*1, i.e., week 10.

**Figure A16:**
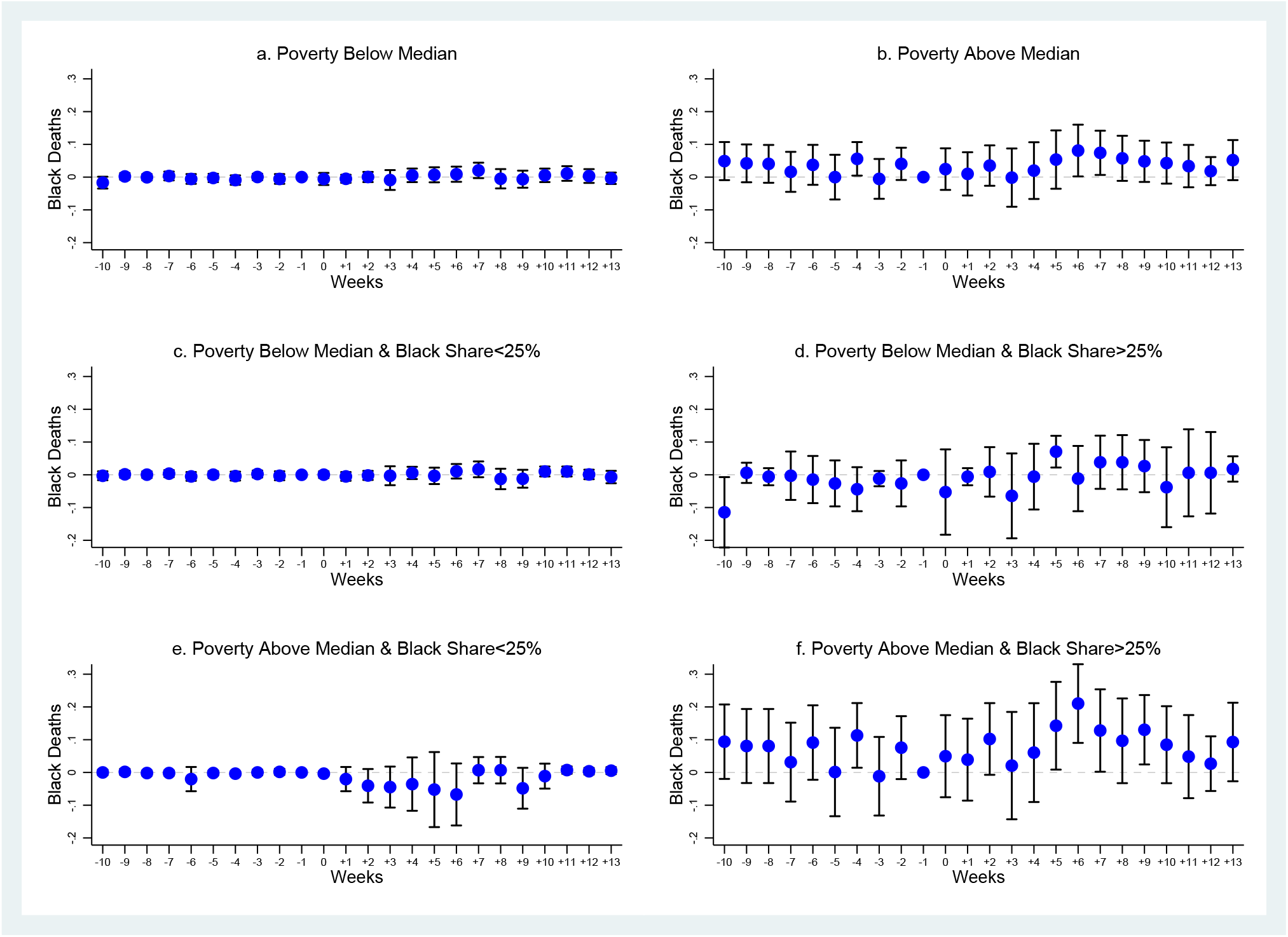
Heterogeneity by Poverty - Cook County, January 1-June 16, 2020. *Note:* The dependent variable is number of black deaths. The coefficients are least-squares estimates of the *β*_*k*_s over samples of block groups with population share below the poverty line below median (Panel a), above median (Panel b), below median and with black share below 25 percent (Panel c), below median and with black share above 25 percent (Panel d), above median and with black share below 25 percent (Panel e), and above median and with black share above 25 percent (Panel f). Block group and week fixed effects are included. Vertical lines represent 95 percent confidence intervals based on standard errors clustered at at block group level. The omitted period *k* = *−*1, i.e., week 10.

**Figure A17:**
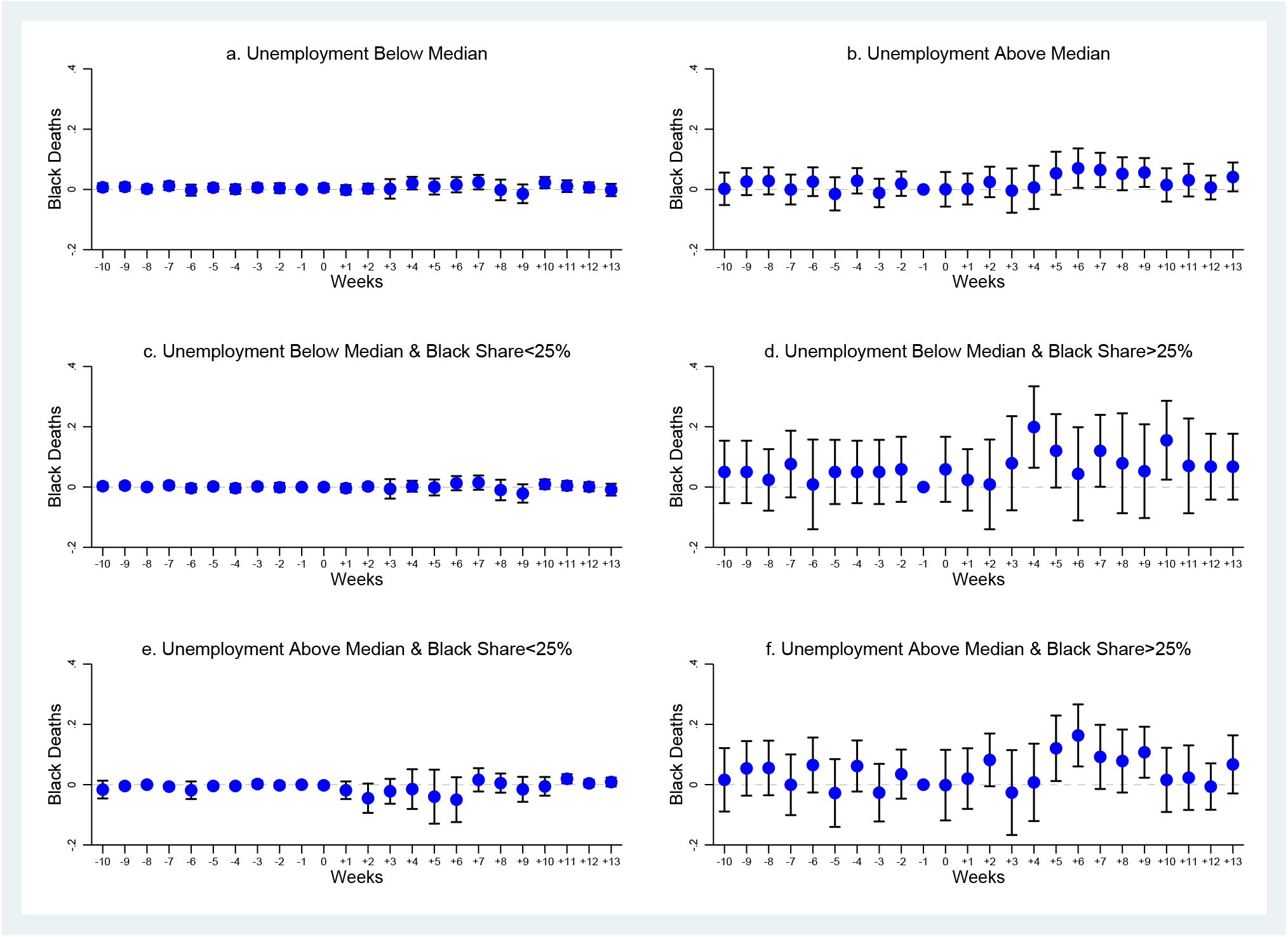
Heterogeneity by Unemployment - Cook County, January 1-June 16, 2020. *Note:* The dependent variable is number of black deaths. The coefficients are least-squares estimates of the *β*_*k*_s over samples of block groups with unemployment rate below median (Panel a), above median (Panel b), below median and with black share below 25 percent (Panel c), below median and with black share above 25 percent (Panel d), above median and with black share below 25 percent (Panel e), and above median and with black share above 25 percent (Panel f). Block group and week fixed effects are included. Vertical lines represent 95 percent confidence intervals based on standard errors clustered at at block group level. The omitted period *k* = *−*1, i.e., week 10.

**Figure A18:**
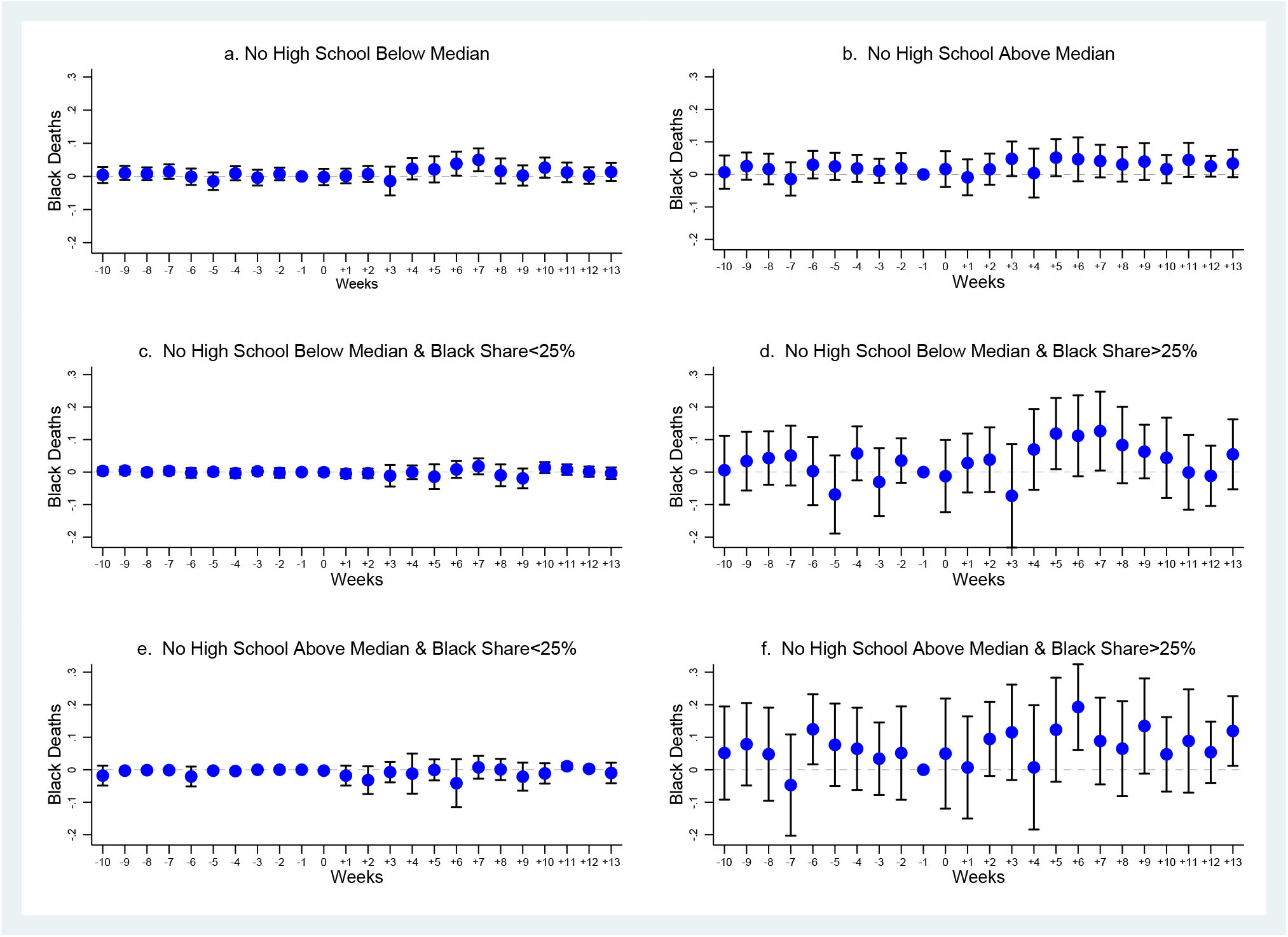
Heterogeneity by Education - Cook County, January 1-June 16, 2020. *Note:* The dependent variable is number of black deaths. The coefficients are least-squares estimates of the *β*_*k*_s over samples of block groups with population share with no high school below median (Panel a), above median (Panel b), below median and with black share below 25 percent (Panel c), below median and with black share above 25 percent (Panel d), above median and with black share below 25 percent (Panel e), and above median and with black share above 25 percent (Panel f). Block group and week fixed effects are included. Vertical lines represent 95 percent confidence intervals based on standard errors clustered at at block group level. The omitted period *k* = *−*1, i.e., week 10.

**Figure A19:**
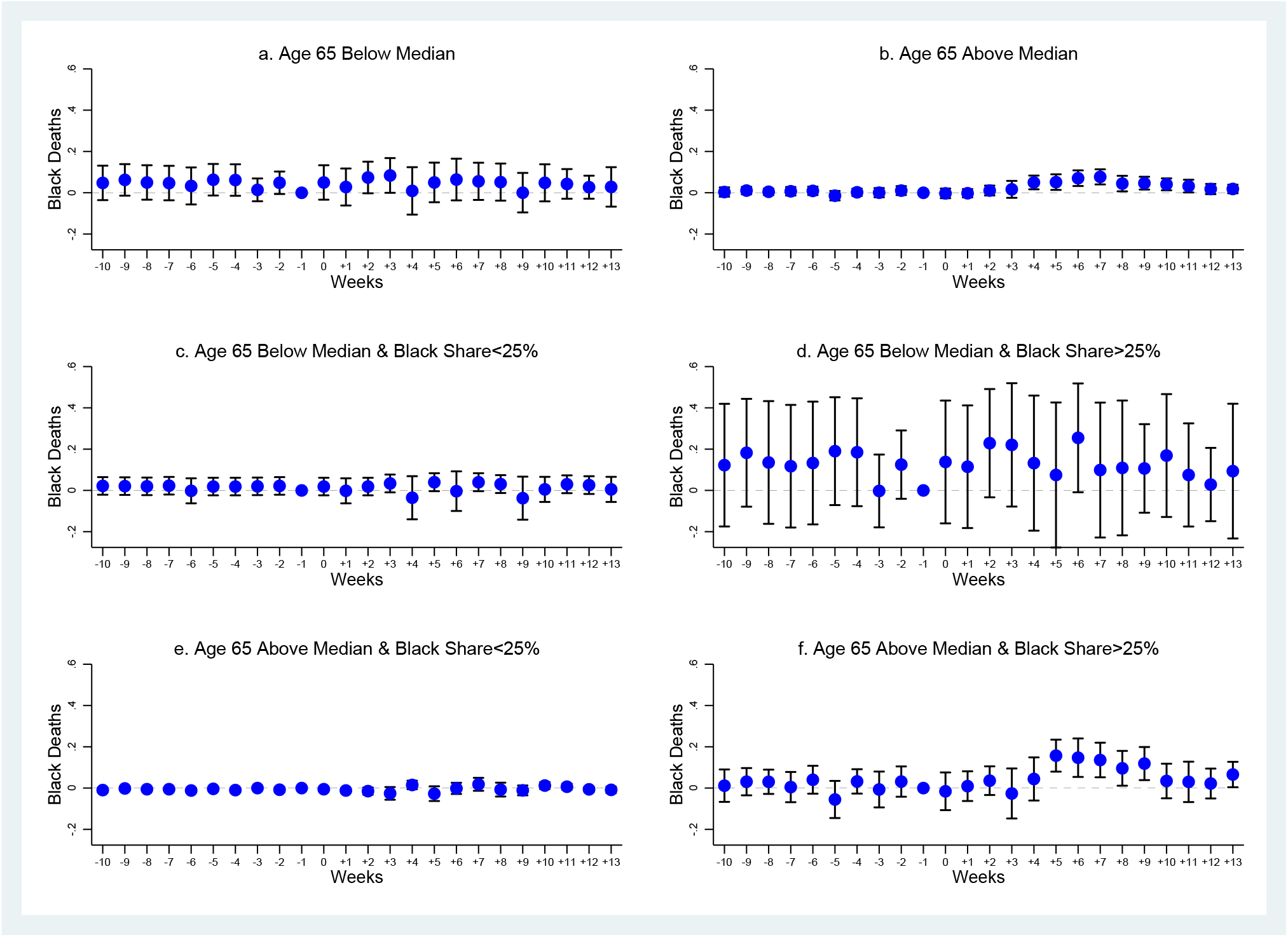
Heterogeneity by Age - Cook County, January 1-June 16, 2020. *Note:* The dependent variable is number of black deaths. The coefficients are least-squares estimates of the *β*_*k*_s over samples of block groups with population share aged 65+ below median (Panel a), above median (Panel b), below median and with black share below 25 percent (Panel c), below median and with black share above 25 percent (Panel d), above median and with black share below 25 percent (Panel e), and above median and with black share above 25 percent (Panel f). Block group and week fixed effects are included. Vertical lines represent 95 percent confidence intervals based on standard errors clustered at at block group level. The omitted period *k* = *−*1, i.e., week 10.

**Figure A20:**
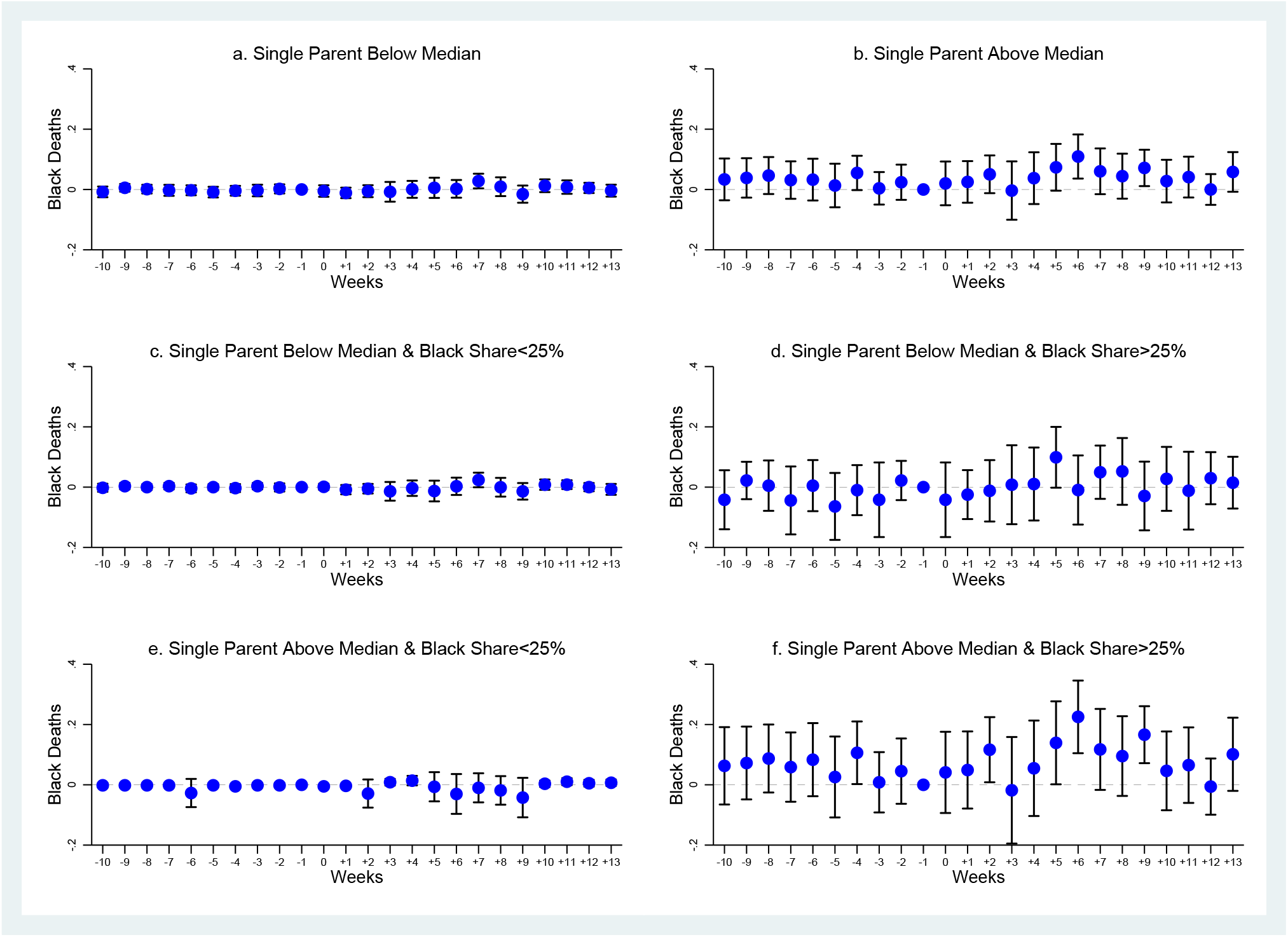
Heterogeneity by Single Parents - Cook County, January 1-June 16, 2020. *Note:* The dependent variable is number of black deaths. The coefficients are least-squares estimates of the *β*_*k*_s over samples of block groups with population share of single parents below median (Panel a), above median (Panel b), below median and with black share below 25 percent (Panel c), below median and with black share above 25 percent (Panel d), above median and with black share below 25 percent (Panel e), and above median and with black share above 25 percent (Panel f). Block group and week fixed effects are included. Vertical lines represent 95 percent confidence intervals based on standard errors clustered at at block group level. The omitted period *k* = *−*1, i.e., week 10.

**Figure A21:**
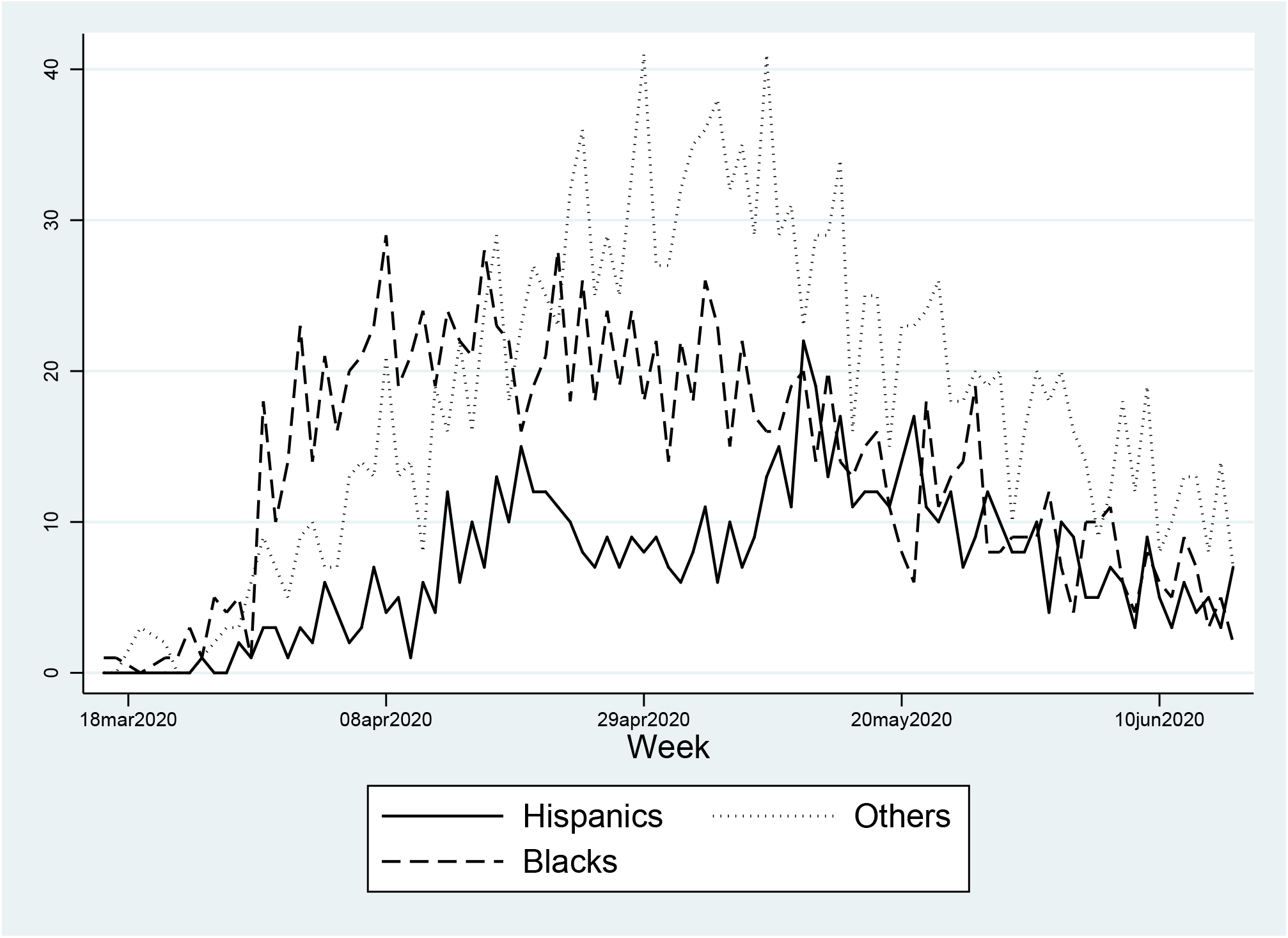
Hispanic, Black, and Other COVID-19 Deaths - Cook County, March 16-June 16, 2020. *Note:* The figure reports the number of COVID-19 related deaths by day, separately for Hispanics, blacks, and other groups.

**Figure A22:**
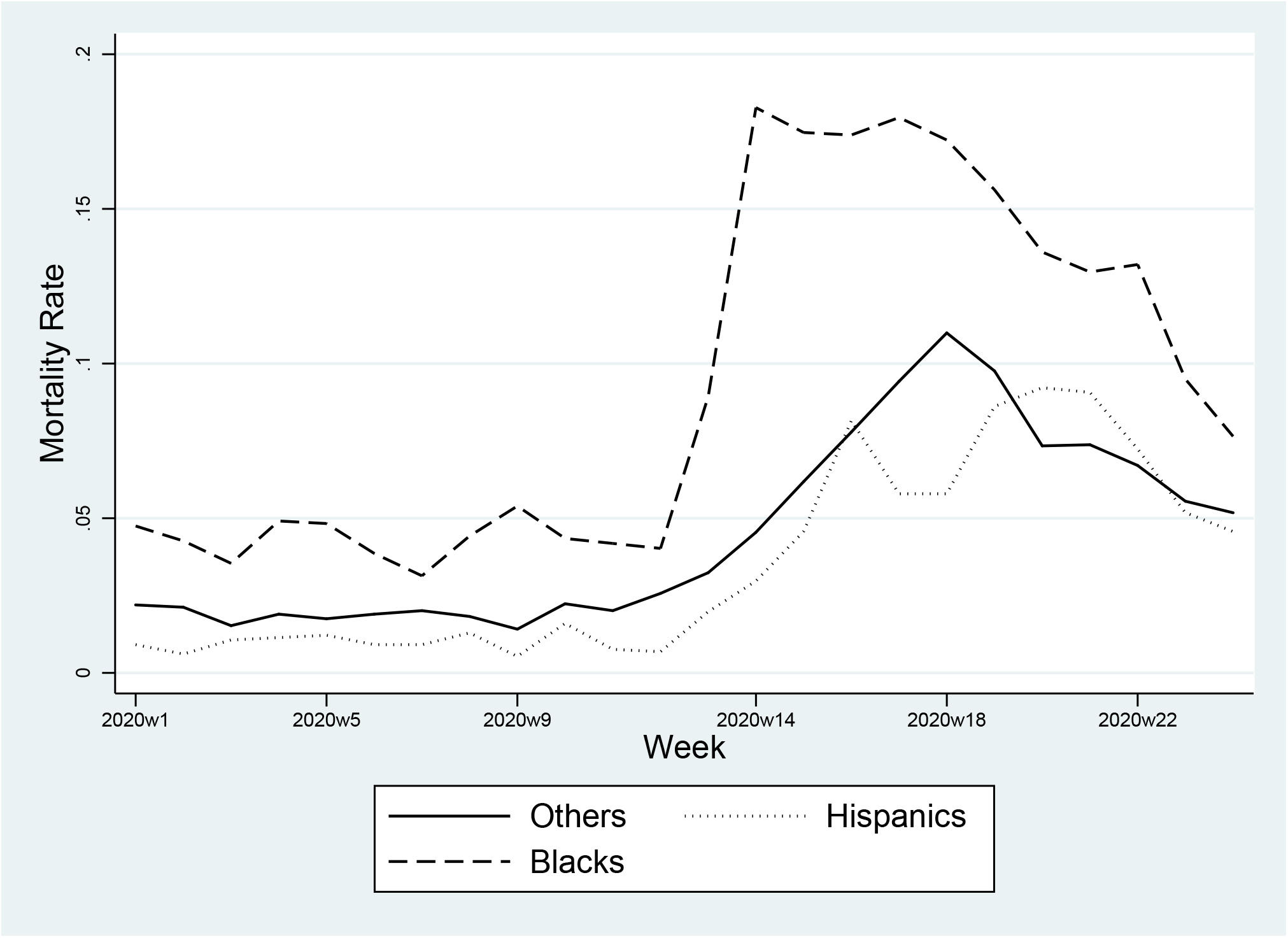
Mortality Rate, Hispanics, Blacks, and Others - Cook County, January 1-June 16, 2020. *Note:* Mortality rates from any cause of death by week, separately for Hispanics, blacks, and other groups.

**Figure A23:**
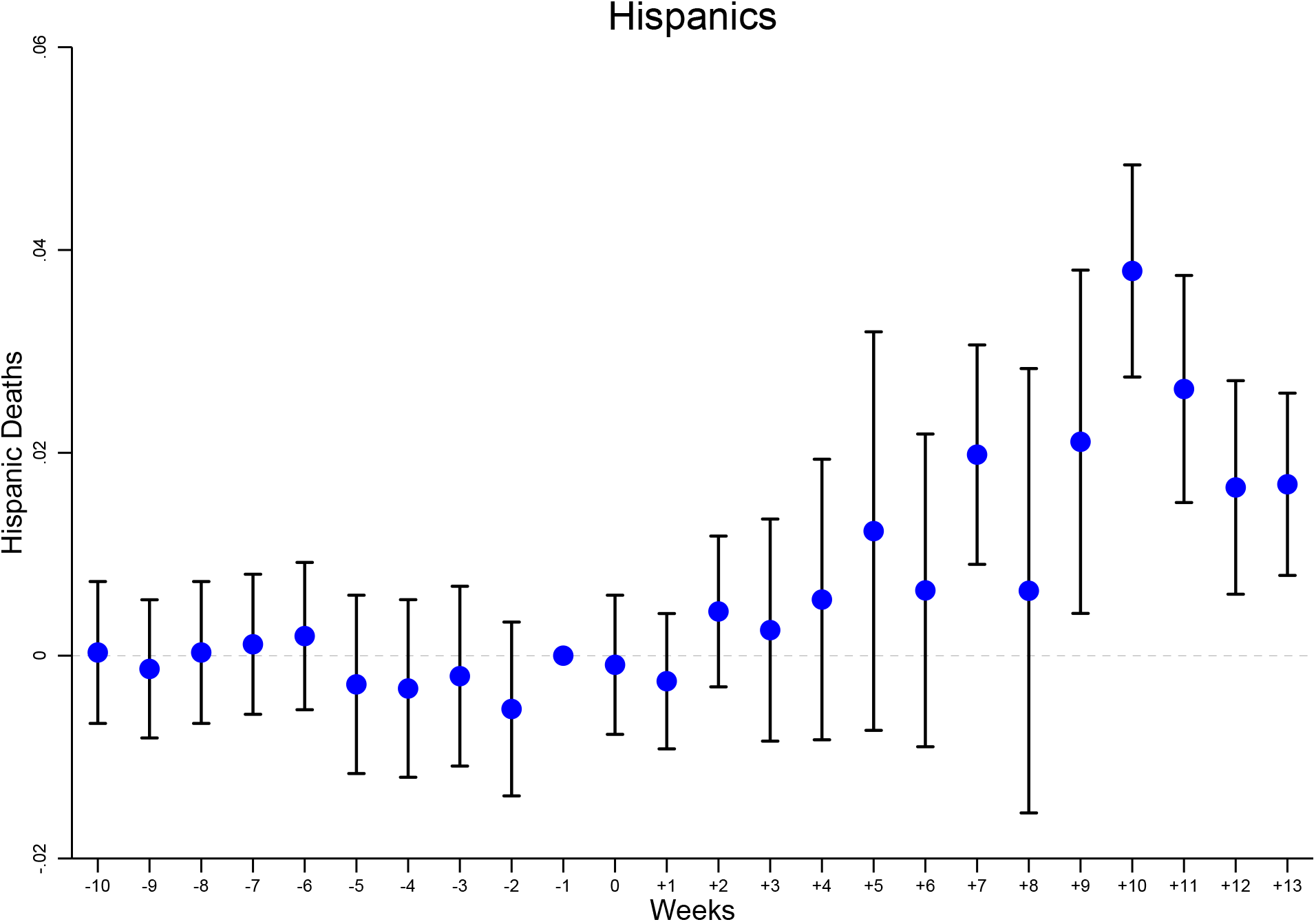
Dynamic Effect of the Treatment on Number of Hispanic Deaths - Cook County, January 1-June 16, 2020. *Note:* The dependent variable is number of Hispanic deaths. The coefficients are least-squares estimates of the *β*_*k*_s with 10*k*9. Block group and week fixed effects are included. Vertical lines represent 95 percent confidence intervals based on standard errors clustered at at block group level. The omitted period *k* = *−*1, i.e., week 10.

**Table A1:** Variable Definitions and Sources - Cross Section.

| Variable | Definition | Source |
| --- | --- | --- |
| Black COVID-19 Death | Dummy variable taking value one if an individual who died from COVID-19 is black, and zero otherwise | Cook County Medical Examiner's Officer |
| Hispanic COVID-19 Death | Dummy variable taking value one if an individual who died from COVID-19 is Hispanic, and zero otherwise | Cook County Medical Examiner's Officer |
| Other COVID-19 Death | Dummy variable taking value one if an individual who died from COVID-19 is other than black, and zero otherwise | Cook County Medical Examiner's Officer |
| HOLC A | Dummy variable taking value one if an individual who died from COVID-19 lived in HOLC Area A, and zero otherwise | University of Richmond Mapping Inequality: Redlining in New Deal America 1935-1940 |
| HOLC B | Dummy variable taking value one if an individual who died from COVID-19 lived in HOLC Area B, and zero otherwise | University of Richmond Mapping Inequality: Redlining in New Deal America 1935-1940 |
| HOLC C | Dummy variable taking value one if an individual who died from COVID-19 lived in HOLC Area C, and zero otherwise | University of Richmond Mapping Inequality: Redlining in New Deal America 1935-1940 |
| HOLC D | Dummy variable taking value one if an individual who died from COVID-19 lived in HOLC Area D, and zero otherwise | University of Richmond Mapping Inequality: Redlining in New Deal America 1935-1940 |
| Age Groups | Set of eight dummy variables taking value one if an individual who died from COVID-19 was respectively aged from 20-29 (tricenarian) up to 100+ (centenarian), and zero otherwise | Cook County Medical Examiner's Officer |
| Female | Dummy variable taking value one if an individual who died from COVID-19 was female | Cook County Medical Examiner's Officer |
| Comorbidities | Set of 14 dummy variables that take value one (and zero otherwise) when an individual who died from COVID-19 was respectively affected by diabetes and/or asthma, liver disease, cancer, hypertension, kidney disease, obesity, respiratory diseases (including cystic fibrosis, pulmonary and lung diseases), neuro-cardiac diseases (including cardiovascular disease, stroke, and dementia), neuro-respiratory diseases (including sclerosis, Parkinson, myasthenia, palsy, hemiplegia, quadriplegia, brain and cerebellum diseases), asplenia (including splenectomy, spleen and sickle cell disease), immunodeficiency (including HIV, immunosuppression, and anaemia), transplant, and heart diseases (including valve disease). | Cook County Medical Examiner's Officer |
| Distance From Hospital | Individual distance (in degrees) from hospital | Cook County Health and Hospitals Facilities |
| Distance From Nursing Home | Individual distance (in degrees) from nursing home | Medicare Nursing Home Compare |
| Population | 2014-2018 tract level population | American Community Survey through Cook County Government Open Data |
| Share Aged 18-64 | 2014-2018 tract level share of the population aged 18-64 | American Community Survey through Cook County Government Open Data |
| Share Aged 65+ | 2014-2018 tract level share of the population aged 65+ | American Community Survey through Cook County Government Open Data |
| Share With No High School | 2014-2018 tract level share of the population without a high school diploma | American Community Survey through Cook County Government Open Data |
| Black Share | 2014-2018 tract level share of the population who is black | American Community Survey through Cook County Government Open Data |
| Hispanic Share | 2014-2018 tract level share of the population who is Hispanic | American Community Survey through Cook County Government Open Data |

**Table A2:**
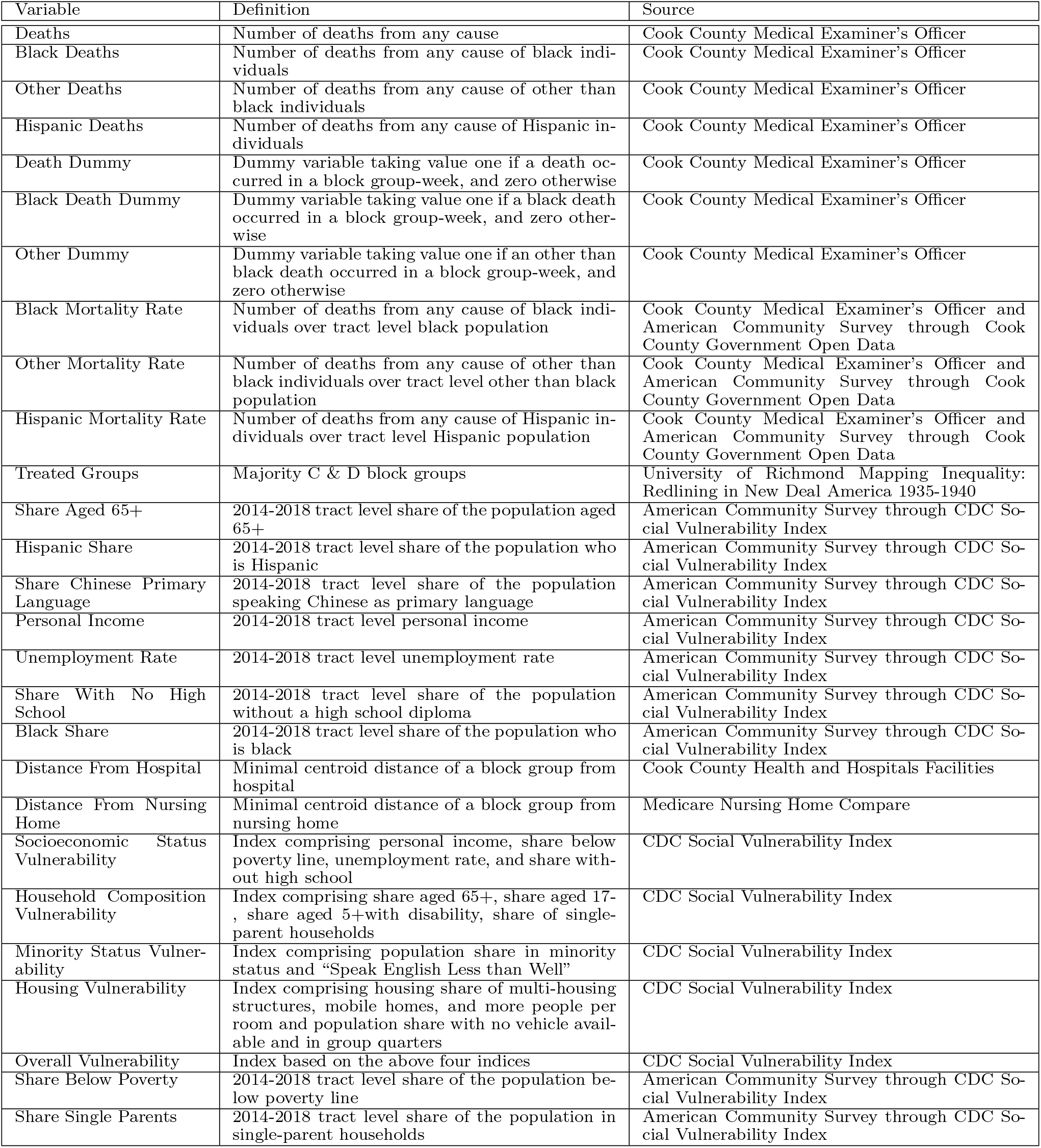
Variable Definitions and Sources - Panel.

**Table A3:**
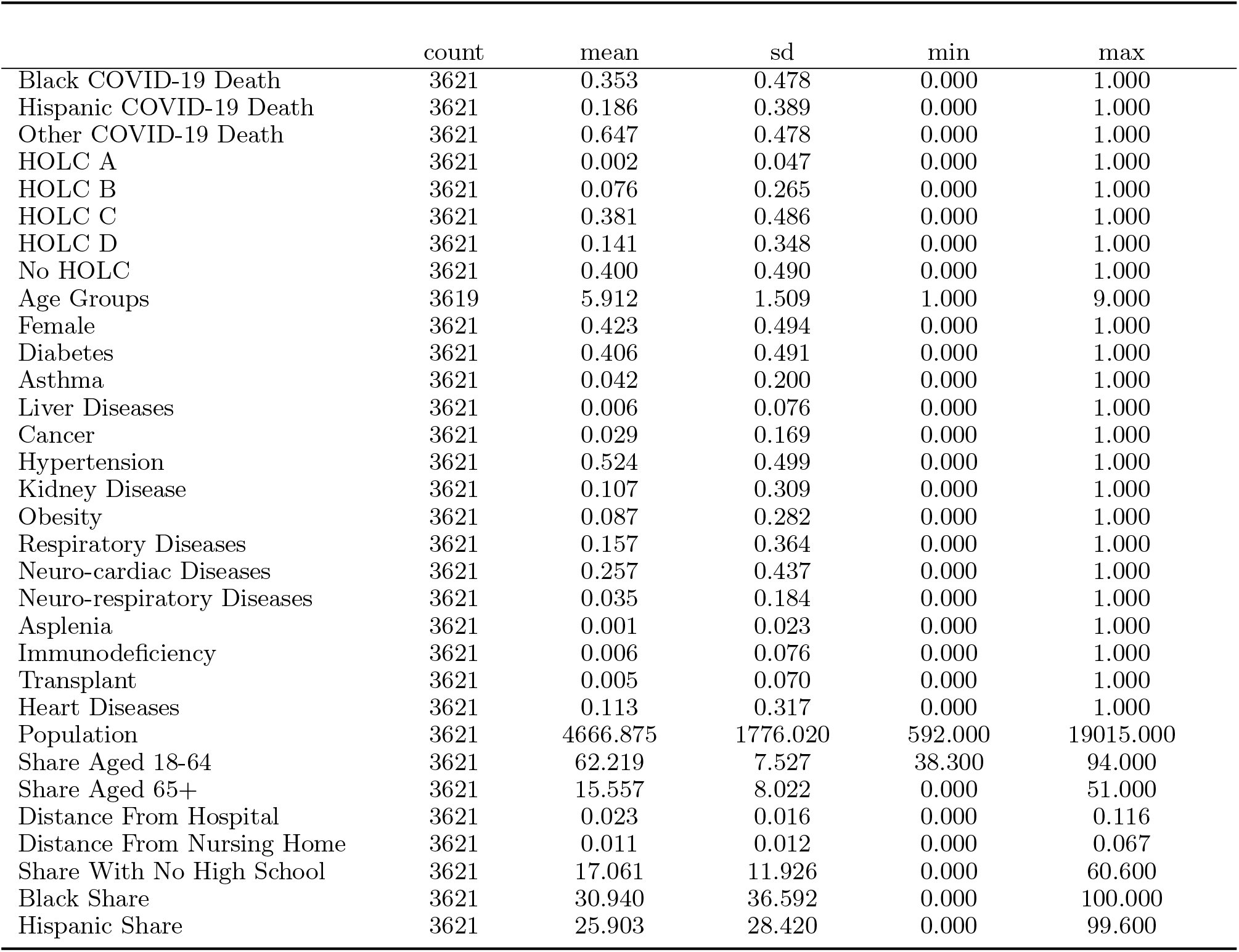
Summary Statistics, COVID-19 Deaths, Cross Section - Cook County, March 16-June 16, 2020.

|  | count | mean | sd | min | max |
| --- | --- | --- | --- | --- | --- |
| Black COVID-19 Death | 3621 | 0.353 | 0.478 | 0.000 | 1.000 |
| Hispanic COVID-19 Death | 3621 | 0.186 | 0.389 | 0.000 | 1.000 |
| Other COVID-19 Death | 3621 | 0.647 | 0.478 | 0.000 | 1.000 |
| HOLC A | 3621 | 0.002 | 0.047 | 0.000 | 1.000 |
| HOLC B | 3621 | 0.076 | 0.265 | 0.000 | 1.000 |
| HOLC C | 3621 | 0.381 | 0.486 | 0.000 | 1.000 |
| HOLC D | 3621 | 0.141 | 0.348 | 0.000 | 1.000 |
| No HOLC | 3621 | 0.400 | 0.490 | 0.000 | 1.000 |
| Age Groups | 3619 | 5.912 | 1.509 | 1.000 | 9.000 |
| Female | 3621 | 0.423 | 0.494 | 0.000 | 1.000 |
| Diabetes | 3621 | 0.406 | 0.491 | 0.000 | 1.000 |
| Asthma | 3621 | 0.042 | 0.200 | 0.000 | 1.000 |
| Liver Diseases | 3621 | 0.006 | 0.076 | 0.000 | 1.000 |
| Cancer | 3621 | 0.029 | 0.169 | 0.000 | 1.000 |
| Hypertension | 3621 | 0.524 | 0.499 | 0.000 | 1.000 |
| Kidney Disease | 3621 | 0.107 | 0.309 | 0.000 | 1.000 |
| Obesity | 3621 | 0.087 | 0.282 | 0.000 | 1.000 |
| Respiratory Diseases | 3621 | 0.157 | 0.364 | 0.000 | 1.000 |
| Neuro-cardiac Diseases | 3621 | 0.257 | 0.437 | 0.000 | 1.000 |
| Neuro-respiratory Diseases | 3621 | 0.035 | 0.184 | 0.000 | 1.000 |
| Asplenia | 3621 | 0.001 | 0.023 | 0.000 | 1.000 |
| Immunodeficiency | 3621 | 0.006 | 0.076 | 0.000 | 1.000 |
| Transplant | 3621 | 0.005 | 0.070 | 0.000 | 1.000 |
| Heart Diseases | 3621 | 0.113 | 0.317 | 0.000 | 1.000 |
| Population | 3621 | 4666.875 | 1776.020 | 592.000 | 19015.000 |
| Share Aged 18-64 | 3621 | 62.219 | 7.527 | 38.300 | 94.000 |
| Share Aged 65+ | 3621 | 15.557 | 8.022 | 0.000 | 51.000 |
| Distance From Hospital | 3621 | 0.023 | 0.016 | 0.000 | 0.116 |
| Distance From Nursing Home | 3621 | 0.011 | 0.012 | 0.000 | 0.067 |
| Share With No High School | 3621 | 17.061 | 11.926 | 0.000 | 60.600 |
| Black Share | 3621 | 30.940 | 36.592 | 0.000 | 100.000 |
| Hispanic Share | 3621 | 25.903 | 28.420 | 0.000 | 99.600 |

**Table A4:** Black COVID-19 Death, Cross-Sectional Results, All Coefficients - Cook County, March 16-June 16, 2020.

|  | Black COVID-19 Death |  |  |  |  |  |  |  |
| --- | --- | --- | --- | --- | --- | --- | --- | --- |
|  | (1) | (2) | (3) | (4) | (5) | (6) | (7) | (8) |
| HOLC A | -0.3808***<br>(0.0640) | -0.3652***<br>(0.0720) | -0.3244***<br>(0.0757) | -0.3261***<br>(0.0800) | -0.3247***<br>(0.0811) | -0.3636***<br>(0.0798) | -0.3514***<br>(0.0751) | -0.1250***<br>(0.0424) |
| HOLC B | 0.0389<br>(0.0329) | 0.0360<br>(0.0334) | 0.0399<br>(0.0327) | 0.0373<br>(0.0323) | 0.0355<br>(0.0327) | 0.0303<br>(0.0322) | 0.0072<br>(0.0316) | 0.0169<br>(0.0250) |
| HOLC C | 0.0800***<br>(0.0177) | 0.0817***<br>(0.0185) | 0.0830***<br>(0.0181) | 0.0677***<br>(0.0175) | 0.0716***<br>(0.0202) | 0.1176***<br>(0.0216) | 0.0848***<br>(0.0234) | -0.0146<br>(0.0182) |
| HOLC D | 0.1752***<br>(0.0233) | 0.1769***<br>(0.0238) | 0.1745***<br>(0.0231) | 0.0833***<br>(0.0255) | 0.0852***<br>(0.0266) | 0.1232***<br>(0.0281) | 0.0922***<br>(0.0297) | 0.0106<br>(0.0222) |
| Tricenarian |  | 0.1373<br>(0.1324) | 0.1069<br>(0.1333) | 0.0898<br>(0.1306) | 0.0934<br>(0.1318) | 0.1113<br>(0.1287) | 0.1244<br>(0.1278) | -0.0055<br>(0.0981) |
| Quadragenarian |  | 0.0593<br>(0.1262) | 0.0282<br>(0.1278) | 0.0278<br>(0.1282) | 0.0326<br>(0.1294) | 0.0600<br>(0.1255) | 0.0728<br>(0.1236) | 0.0233<br>(0.0961) |
| Quinquagenarian |  | 0.0559<br>(0.1225) | 0.0140<br>(0.1229) | 0.0198<br>(0.1230) | 0.0252<br>(0.1240) | 0.0480<br>(0.1211) | 0.0648<br>(0.1192) | 0.0121<br>(0.0895) |
| Sexagenarian |  | 0.1348<br>(0.1200) | 0.0778<br>(0.1216) | 0.0795<br>(0.1207) | 0.0857<br>(0.1220) | 0.0907<br>(0.1197) | 0.1144<br>(0.1178) | 0.0603<br>(0.0911) |
| Septuagenarian |  | 0.1149<br>(0.1171) | 0.0543<br>(0.1178) | 0.0619<br>(0.1177) | 0.0678<br>(0.1189) | 0.0672<br>(0.1168) | 0.0890<br>(0.1145) | 0.0288<br>(0.0892) |
| Octogenarian |  | 0.0768<br>(0.1188) | 0.0148<br>(0.1192) | 0.0155<br>(0.1200) | 0.0218<br>(0.1212) | 0.0140<br>(0.1196) | 0.0409<br>(0.1182) | 0.0180<br>(0.0891) |
| Nonagenarian |  | 0.0082<br>(0.1226) | -0.0498<br>(0.1225) | -0.0433<br>(0.1233) | -0.0374<br>(0.1248) | -0.0468<br>(0.1230) | -0.0102<br>(0.1203) | -0.0291<br>(0.0921) |
| Centenarian |  | 0.0986<br>(0.1350) | 0.0372<br>(0.1358) | 0.0399<br>(0.1337) | 0.0445<br>(0.1356) | 0.0270<br>(0.1338) | 0.0744<br>(0.1311) | 0.0061<br>(0.1046) |
| Female |  | 0.1036***<br>(0.0156) | 0.0977***<br>(0.0157) | 0.0961***<br>(0.0154) | 0.0955***<br>(0.0154) | 0.0886***<br>(0.0155) | 0.0902***<br>(0.0155) | 0.0504***<br>(0.0115) |
| Diabetes |  |  | 0.0040<br>(0.0156) | -0.0004<br>(0.0155) | -0.0003<br>(0.0154) | 0.0103<br>(0.0152) | 0.0092<br>(0.0149) | 0.0005<br>(0.0122) |
| Asthma |  |  | 0.0145<br>(0.0440) | 0.0155<br>(0.0438) | 0.0147<br>(0.0438) | 0.0085<br>(0.0427) | -0.0003<br>(0.0427) | -0.0034<br>(0.0314) |
| Liver Diseases |  |  | -0.1527<br>(0.0948) | -0.1488*<br>(0.0870) | -0.1498*<br>(0.0868) | -0.1516*<br>(0.0820) | -0.1496*<br>(0.0801) | 0.0089<br>(0.0771) |
| Cancer |  |  | 0.0722<br>(0.0478) | 0.0747<br>(0.0467) | 0.0753<br>(0.0474) | 0.0670<br>(0.0480) | 0.0580<br>(0.0477) | 0.0166<br>(0.0395) |
| Hypertension |  |  | 0.0939***<br>(0.0165) | 0.0936***<br>(0.0166) | 0.0938***<br>(0.0166) | 0.0908***<br>(0.0167) | 0.0881***<br>(0.0167) | 0.0545***<br>(0.0142) |
| Kidney Disease |  |  | 0.0782***<br>(0.0272) | 0.0806***<br>(0.0266) | 0.0802***<br>(0.0268) | 0.0694**<br>(0.0266) | 0.0702**<br>(0.0269) | 0.0659***<br>(0.0213) |
| Obesity |  |  | 0.0413<br>(0.0297) | 0.0305<br>(0.0287) | 0.0306<br>(0.0289) | 0.0290<br>(0.0281) | 0.0264<br>(0.0280) | 0.0118<br>(0.0234) |
| Respiratory Diseases |  |  | 0.0764***<br>(0.0214) | 0.0740***<br>(0.0209) | 0.0747***<br>(0.0209) | 0.0700***<br>(0.0203) | 0.0752***<br>(0.0201) | 0.0405**<br>(0.0175) |
| Neuro-cardiac Diseases |  |  | 0.0158<br>(0.0213) | 0.0166<br>(0.0214) | 0.0177<br>(0.0217) | 0.0196<br>(0.0217) | 0.0236<br>(0.0213) | 0.0453***<br>(0.0165) |
| Neuro-respiratory Diseases |  |  | -0.0997***<br>(0.0338) | -0.0951***<br>(0.0333) | -0.0943***<br>(0.0330) | -0.0986***<br>(0.0327) | -0.0843**<br>(0.0338) | -0.0739***<br>(0.0261) |
| Asplenia |  |  | 0.1501<br>(0.3863) | 0.1260<br>(0.3213) | 0.1394<br>(0.3237) | 0.1531<br>(0.3323) | 0.1704<br>(0.3184) | -0.1478<br>(0.1288) |
| Immunodeficiency |  |  | 0.4063***<br>(0.0899) | 0.3897***<br>(0.0934) | 0.3927***<br>(0.0935) | 0.3756***<br>(0.0883) | 0.3685***<br>(0.0877) | 0.2470**<br>(0.1020) |
| Transplant |  |  | 0.1204<br>(0.1186) | 0.1200<br>(0.1159) | 0.1177<br>(0.1166) | 0.0940<br>(0.1071) | 0.0805<br>(0.1071) | 0.0136<br>(0.1024) |
| Heart Diseases |  |  | 0.0256<br>(0.0267) | 0.0178<br>(0.0271) | 0.0180<br>(0.0271) | 0.0093<br>(0.0266) | 0.0157<br>(0.0259) | 0.0081<br>(0.0197) |
| Population (log) |  |  |  | -0.1734***<br>(0.0210) | -0.1725***<br>(0.0208) | -0.1780***<br>(0.0209) | -0.1568***<br>(0.0207) | -0.0260<br>(0.0178) |
| Share Aged 18-64 |  |  |  |  | -0.0016<br>(0.0014) | -0.0065***<br>(0.0018) | -0.0067***<br>(0.0017) | 0.0014<br>(0.0012) |
| Share Aged 65+ |  |  |  |  | -0.0007<br>(0.0013) | -0.0065***<br>(0.0015) | -0.0058***<br>(0.0016) | -0.0011<br>(0.0011) |
| Share With No High School |  |  |  |  |  | -0.0073***<br>(0.0009) | -0.0073***<br>(0.0008) | -0.0007<br>(0.0006) |
| Distance From Hospital |  |  |  |  |  |  | -2.9815***<br>(0.4945) | 0.4574<br>(0.4217) |
| Distance From Nursing Home |  |  |  |  |  |  | 3.5242***<br>(0.6538) | -1.5140***<br>(0.5396) |
| Black Share |  |  |  |  |  |  |  | 0.0086***<br>(0.0002) |
| Constant | 0.2957***<br>(0.0094) | 0.1637<br>(0.1185) | 0.1384<br>(0.1233) | 1.6084***<br>(0.2004) | 1.6999***<br>(0.2262) | 2.2492***<br>(0.2516) | 2.0984***<br>(0.2511) | 0.1585<br>(0.2039) |
| Adj.R-squared | 0.056 | 0.069 | 0.090 | 0.111 | 0.111 | 0.132 | 0.144 | 0.475 |
| Observations | 3618 | 3616 | 3616 | 3616 | 3616 | 3616 | 3616 | 3616 |

Note: The dependent variable is a dummy variable that takes value one if an individual who died from COVID-19 is black, an zero otherwise. The omitted HOLC area is A. The omitted age group is vicanarian. Robust standard errors clustered at a day level in parentheses: \*\*\* p<0.01, \*\* p<0.05, \* p<0.1.

**Table A5:**
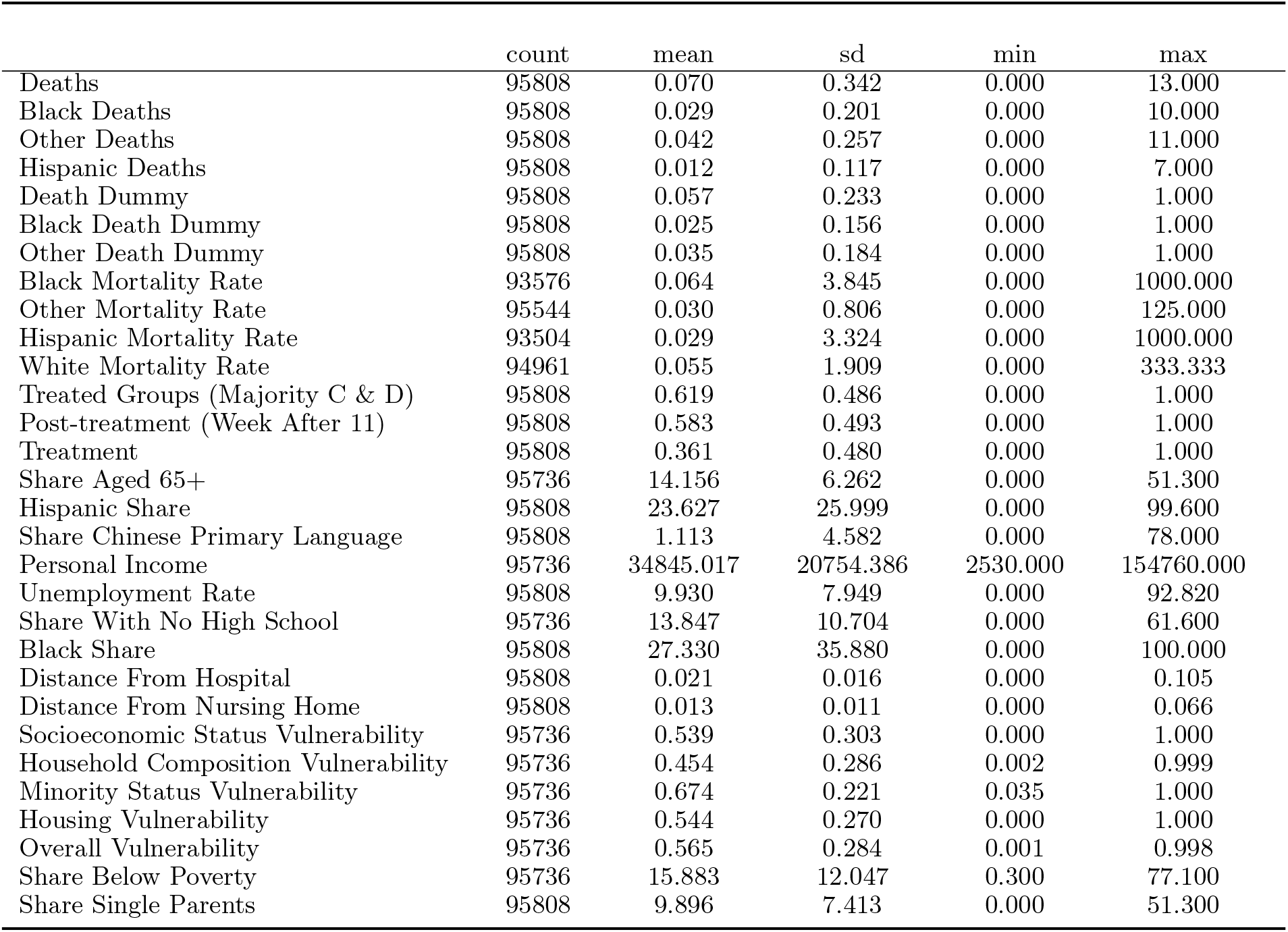
Summary Statistics, All Deaths, Panel - Cook County, January 1-June 16, 2020.

**Table A6:**
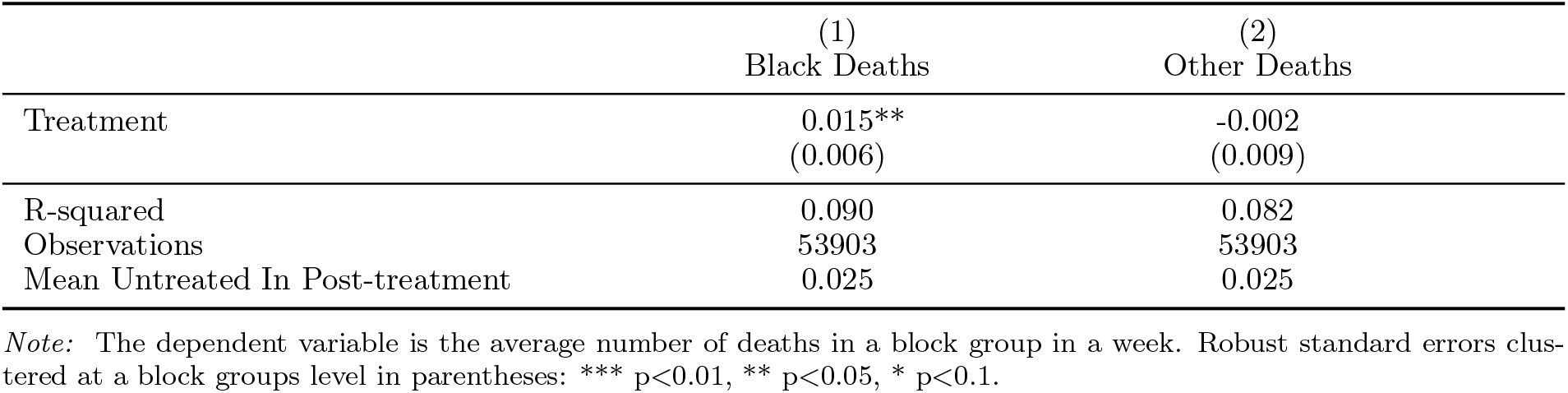
Average Treatment Effect.

**Table A7:**
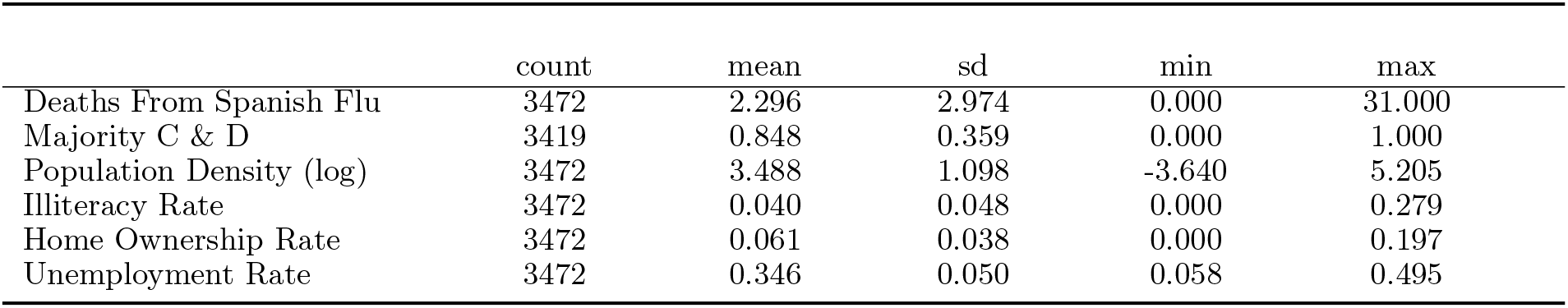
Summary Statistics, Spanish Flu Deaths, Chicago, 1918.

## Footnotes

1 Since April 15, after some states started to report data, the COVID Tracking Project at The Atlantic has been providing updated state-level race and ethnicity data on cases and deaths, with information limited to race and ethnicity. See https://covidtracking.com/race. On July 5, 2020 The New York Times (Oppel et al., 2020) reported data on 640,000 individual cases, by race, ethnicity and home county, collected through May 28. The data were acquired after filing a Freedom of Information Act suit against the Centers for Disease Control and Prevention.

2 See Eligon et al. (2020) in The New York Times.

3 *“They are the outcomes not just of the immediate moments in time, but they are the result of a long history of slavery and Jim Crow and redlining and institutionalized racism that too often have been the plague, the original sin, of our society.”* See https://www.youtube.com/watch?v=q_qB6SsErpA.

4 In the UK, the racial issue came to public attention when the first eleven doctors who died from COVID-19 were all reported to belong to black, Asian, and minority ethnic communities (Kirby, 2020).

5 For the much broader literature on racial discrimination in the US, see Lang and Kahn-Lang Spitzer (2020) for a survey and Nunn (2008), Bertocchi and Dimico (2012, 2014, 2020a), Bertocchi (2016), and references therein, on the legacy of slavery as its long-term determinant.

6 On residential segregation and cancer see also Landrine et al. (2017). An extensive literature confronts racial and ethnic disparities in health care and health outcomes. See Institute of Medicine (2003) and Orsi et al. (2010).

7 On the history of the HOLC, see Fishback et al. (2013). On the broader determinants, other than redlining, of race segregation in the metropolitan areas of the US, see Boustan (2011) and Rothstein (2017). On the influence of zoning policies, with a focus on 1923 Chicago, see Shertzer et al. (2016).

8 Subsequently, the activities of the HOLC were devoted to loan management and repayments. Operations were officially ceased in 1951 and termination was ordered in 1954.

9 See https://www.census.gov/quickfacts/fact/table/cookcountyillinois/PST120219.

10 Racial and ethnic data are based on self-identification. Reported figures are “alone”, that is, refer to those individuals that self-classify themselves as belonging to a single racial or ethnic group.

11 See https://coronavirus.jhu.edu/.

12 See Nelson et al. (2020) and https://dsl.richmond.edu/panorama/redlining/.

13 See Cook County Medical Examiner COVID-19 Related Deaths at https://datacatalog.cookcountyil.gov/Public-Safety/Medical-Examiner-Case-Archive-COVID-19-Related-Dea/3trz-enys.

14 Data refer to deaths for which COVID-19 is reported among either primary or secondary causes. Operationally, the Medical Examiner’s Officer looks for references to COVID-19 in any of these fields: Primary Cause, Primary Cause Line A, Primary Cause Line B, Primary Cause Line C, or Secondary Cause.

15 Namely, the Medical Examiner’s Office investigates any human death that falls within any or all of the following categories: criminal violence; suicide; accident; suddenly when in apparent good condition; unattended by a practicing licensed physician; suspicious or unusual circumstances; unlawful fetal death as provided in Public Act 101-0013 of the 101st General Assembly of Illinois; poisoning or attributable to an adverse reaction to drugs and/or alcohol; disease constituting a threat to public health; injury or toxic agent resulting from employment; during some medical diagnostic or therapeutic procedures; in any prison or penal institution; when involuntarily confined in jail prison hospitals or other institutions or in police custody; when any human body is to be cremated, dissected or buried at sea; when a dead body is brought into a new medico-legal jurisdiction without proper medical certification. Overall, each year, more than 16,000 deaths are reported to the Cook County Medical Examiner. See https://www.cookcountyil.gov/agency/medical-examiner.

16 Data may temporarily differ from those provided by the departments of public health because of time lags in notification.

17 Data are obtained from vital records, hospitals, and families.

18 The racial distribution remains very similar, whether or not unreferenced individuals are included.

19 On April 7, on the basis of the Medical Examiner’s data, the Chicago Tribune (Reys et al. 2020)—echoed by the Journal of the American Medical Association (Yancy, 2020) and the Lancet (Bhala, 2020)—reported that 68 percent of the dead in the City of Chicago involved African Americans, who represent about 30 percent of the city’s population. As of June 16, African Americans account for less than 42 percent of the deaths in Chicago. Thus, the same trend can be detected both for the City of Chicago and Cook County as a whole.

20 Only four deaths are reported below age 20, for three blacks aged 19, 18 and below 1, respectively, and a white aged 12. Figures are not normalized by the size of the population in each age group.

21 Again, figures are not normalized by the degree of feminization of the population.

22 Disease groupings followed those employed by OpenSAFELY Collaborative (2020). Groups are not mutually exclusive.

23 A block groups represents a combination of census blocks and a subdivision of a census tract. A block group is defined to contain between 600 and 3,000 individuals.

24 See Nelson et al. (2020) and https://dsl.richmond.edu/panorama/redlining/.

25 See Cook County Government Open Data at https://datacatalog.cookcountyil.gov/GIS-Maps/2010-U-S-Census-Mail-Return-Rates-and-Demographics/mpyu-4jqk. Data come from the American Community Survey estimates and are averages over the period 2014-2018 available by census tract

26 We take georeferenced hospital location from the Cook County Government (see Cook County Health and Hospitals Facilities at https://datacatalog.cookcountyil.gov/Economic-Development/Cook-County-Health-and-Hospitals-Facilities/jdix-z6uf) and georeferenced nursing home location from the Medicare Nursing Home Compare dataset (see https://data.medicare.gov/data/nursing-home-compare).

27 The Illinois Department of Public Health estimates that about 40 percent of the COVID-19 deaths in Cook County occurred in a long-term facility. See http://www.dph.illinois.gov/covid19/long-term-care-facility-outbreaks-covid-19.

28 See https://svi.cdc.gov/data-and-tools-download.html and Flanagan et al. (2011).

29 Some of the variables, e.g., personal income and unemployment, coincide with those obtained through the Cook County Government from the same source, i.e., the American Community Survey.

30 The reason why probabilities across the four HOLC areas do not sum up to one is that some neighborhoods of Cook County were not mapped by HOLC. 40 percent of the COVID-19 deaths occurred in ungraded neighborhoods of the county.

31 The similarity between the effects of red and yellowlining is consistent with the evidence presented by Aaronson et al. (2017) over a sample of US cities.

32 The merged dataset includes over 3,992 block groups and 1,318 census tracts.

33 This implies that between March 16 and June 16 there are more than 1,871 deaths in addition to COVID-19 related deaths, which is consistent with the 1,256 deaths reported before March 16.

34 As we examine how mortality evolves before and after the epidemic outbreak, we should keep in mind that, generally speaking, officially reported COVID-19 deaths do not match excess deaths, as measured by the gap between observed deaths after the epidemic outbreak and deaths observed during the same period in normal years. However, since our source is the Medical Examiner, the discrepancy between recorded COVID-19 and excess deaths is greatly alleviated, because of the specific nature of the deaths under his jurisdiction. Consequently, the deaths data we use in the cross-sectional analysis and those we use in the event study are directly comparable, provided that it remains possible that deaths from other causes under the Medical Examiner’s jurisdiction may also have risen or declined for various reasons (e.g., more individuals may have died because they did not receive care for other diseases, but fewer may have died for car accidents due to the lockdown). Indeed the Medical Examiner reports, to June 16, 2020, over 3,000 excess deaths compared with the same period in 2019, a figure that closely matches the COVID-19 deaths that were recorded.

35 To compute mortality rates, i.e., number of deaths over population, we first sum weekly deaths at a census tract level (because population data is at a tract level) and then we collapse by week.

36 The alternative would be to use the mortality rate as a dependent variable. However, we have racially-disaggregated data on population only at a census tract level rather than at block group level. In addition, block group fixed effects should absorb differences in population, given that the latter should remain constant over the span of only 24 weeks. In other words, using mortality data would amount to divide deaths by a constant, which should be picked up by the fixed effect.

37 A robustness check that includes ungraded neighborhoods actually leads to even sharper results, reported in Figure A11 that are omitted for brevity and available upon request.

38 Similar results obtain, as shown in Figure A12, if we replace the dependent variable with a dummy variable capturing the occurrence of a death. This is also addressing the issue raised in footnote 36, since the definition of the dependent variable as a dummy does not reflect the distinction between number of deaths and mortality rate. Other robustness checks which we do not report for brevity include slightly changing the definition of the treatment group (Majority C & D may include some A and B neighborhoods, so we also redefine the treatment group by excluding As, thus making the criterion more stringent) and using distributed and error lag models to respectively account for spillovers and serial correlation in the error.

39 This means that we drop block groups for which the majority of the surface area falls in grade B and C areas.

40 We choose this threshold because it is close to the average black share in the county. Furthermore, a higher threshold would greatly reduce the size of the sub-sample displaying a relatively higher black share.

41 See https://data.medicare.gov/data/nursing-home-compare.

42 See, for instance, Singh and Koran (2020) and Yancy (2020).

43 The Medical Examiner’s racial classifications are based on US Census Bureau categories, according to which Latino, or equivalently Hispanic, is defined as ethnicity, and can therefore belong to any racial group. This section focuses on white Hispanics, who in Cook County represent the vast majority of Hispanics. To June 16, only 13 deaths were reported for black Hispanics, i.e., 0.09 percent of the black deaths, and 1.6 percent of the Hispanic deaths.

44 Data for the years 2017-2019 are also provided by the Medical Examiner’s Office.

45 See http://www.ufiddynamics.org/data. The sources of the data are the 1920 Census and the 1920 annual report of City of Chicago Department of Health.

46 See Table A7 for summary statistics.

47 In a companion paper (Bertocchi and Dimico, 2020b) we examine COVID-19 outcomes along the gender and work dimensions.

## Notes

### Competing Interest Statement

The authors have declared no competing interest.

### Funding Statement

No external funding was received

## REFERENCES

Aaronson D., Hartley D., Mazumder B. (2017), The effects of the 1930s HOLC “redlining” maps. Federal Reserve Bank of Chicago Working Paper No. 2017-12.

Abraido-Lanza A.F., Dohrenwend B.P; Ng-Mak D.S., Turner J.B. (1999). The Latino mortality paradox: A test of the “salmon bias” and healthy migrant hypotheses. American Journal of Public Health, 89, (10): 15431548.

Alfani G., Murphy T. (2017), Plague and lethal epidemics in the pre-industrial World. Journal of Economic History, 77, 314–343.

Almond, D. (2006). Is the 1918 influenza pandemic over? Long-term effects of in utero influenza exposure in the post-1940 U.S. population. Journal of Political Economy, 114, 672–712.

Aassve A., Alfani G., Gandolfi F., Le Moglie M. (2020), Epidemics and trust: The case of the Spanish flu. IGIER Working Paper No. 661.

Almagro M., Orane-Hutchinson A. (2020), The determinants of the differential exposure to COVID-19 in New York City and their evolution over time. Covid Economics, 13, 31–50.

Anders J. (2019, The long run effects of de jure discrimination in the credit market: How redlining increased crime. Mimeo.

Anderson S. (2018), Legal origins and female HIV. American Economic Review, 108, 1407–1439.

Appel I., Nickerson J. (2016), Pockets of poverty: the long-term effects of redlining. Mimeo.

Bertocchi G. (2016), The legacies of slavery in and out of Africa. IZA Journal of Migration, 5, 1–19.

Bertocchi G., Dimico A. (2020a), Bitter sugar: Slavery and the black family. CEPR Discussion Paper No. 14837.

Bertocchi G., Dimico A. (2020a), COVID-19, gender, and work. Mimeo.

Bertocchi G., Dimico A. (2019), The long-term determinants of female HIV infection in Africa: The slave trade, polygyny, and sexual behavior. Journal of Development Economics, 140, 90–105.

Bertocchi G., Dimico A. (2014), Slavery, education, and inequality. European Economic Review, 70, 197–209.

Bertocchi G., Dimico A. (2012), The racial gap in education and the legacy of slavery. Journal of Comparative Economics, 40, 581–595.

Betancur J.J. (1996), The settlement experience of Latinos in Chicago: Segregation, speculation, and the ecology model. Source: Social Forces, 74, 1299–1324.

Bhala N., Curry G., Martineau A.R., Agyemang C., Bhopal R. (2020). Sharpening the global focus on ethnicity and race in the time of COVID-19. Lancet. Comment. May 8.

Borjas G.J. (2020), Demographics determinants of testing incidence and Covid-19 infections in New York City neighborhoods. NBER Working Paper 26952.

Boustan L.P. (2011), Racial residential segregation in American cities. In Brooks N., Donaghy K., Knaap G.-J. (eds.), Oxford Handbook of Urban Economics and Planning. New York: Oxford University Press, 318–339.

Cage J., Rueda V. (2020), Sex and the mission: The conflicting effects of early Christian investments on sub-Saharan Africa’s HIV epidemic. Journal of Demographic Economics, forthcoming.

Desmet K., Wacziarg R. (2020), Understanding spacial variation in COVID-19 across the United States. CEPR Discussion Paper No. 14842.

Douglas Commission [National Commission on Urban Problems] (1968), Building the American City. Washington: GPO.

Eligon J., Burch A.D.S., Searcey D., Oppel R.A.Jr. (2020), Black Americans face alarming rates of coronavirus infection in some states. The New York Times, April 7.

Couch K., Fairlie R.W., Xu H. (2020), The impacts of COVID-19 on minority unemployment: First evidence from April 2020 CPS microdata. IZA Discussion Paper No. 13264.

Federal Housing Administration (1938), Underwriting Manual: Underwriting and Valuation Procedure Under Title II of the National Housing Act. Washington: Federal Housing Administration.

Fishback P., Rose J., Snowden K. (eds.) (2013), Well Worth Saving: How the New Deal Safeguarded Home Ownership. Chicago: University of Chicago Press.

Flanagan B., Gregory E., Hallisey E., Heitgerd J., Lewis B. (2011), A social vulnerability index for disaster management. Journal of Homeland Security and Emergency Management, 8, 1–22.

Gamble V.N. (2010), “There Wasnt a Lot of Comforts in Those Days:” African Americans, Public Health, and the 1918 Influenza Epidemic. Public Health Reports, Supplement 3, 125, 114–121.

Greer J.L. (2014), Historic home mortgage redlining in Chicago. Journal of the Illinois State Historical Society, 107, 204–233.

Greer J.L. (2012), Race and mortgage redlining in the United States. Mimeo.

Guimbeau A., Menon N., Musacchio A. (2020), The Brazilian bombshell? The longterm impact of the 1918 influenza pandemic the South American Way. NBER Working Paper No. 26929.

Harriss C.L. (1951), History and Policies of the Home Owners’ Loan Corporation. New York: NBER.

Helgertz J., Bengtsson T. (2019), The long-lasting influenza: The impact of fetal stress during the 1918 influenza pandemic on socioeconomic attainment and health in Sweden, 1968-2012. Demography, 56, 1389–1425.

Hillier A. (2005), Residential Security Maps and neighborhood appraisals: The Home Owners Loan Corporation and the case of Philadelphia. Social Science History, 29, 207–233.

Hillier A. (2003), Redlining and the Home Owners Loan Corporation. Journal of Urban History, 29, 394–420.

Hoyt H. (1933), One Hundred Years of Land Values in Chicago: The Relationship of the Growth of Chicago to the Rise of Its Land Values, 1830-1933. Chicago: University of Chicago Press.

Hunt D.B. (2009), Blueprint for Disaster: The Unraveling of Chicago Public Housing. Chicago: University of Chicago Press.

Institute of Medicine (2003), Unequal Treatment: Confronting Racial and Ethnic Disparities in Health Care. Washington: National Academies Press.

Kendi, I.X. (2020), Stop looking away from the race of COVID-19 victims. The Atlantic, April 1.

Jackson K. (1985), Crabgrass Frontier: The Suburbanization of the United States. New York: Oxford University Press.

Jackson K. (1980), Race, ethnicity, and real estate appraisal: The Home Owners Loan Corporation and the Federal Housing Administration. Journal of Urban History, 6, 419–452.

Jedwab R., Johnson N.D., Koyama M. (2019), Negative shocks and mass persecutions: Evidence from the black death. Journal of Economic Growth, 24, 345–395.

Jedwab R., Johnson N.D., Koyama M. (2016), Bones, bacteria and break points: The heterogeneous spatial effects of the Black Death and long-run growth. GMU Working Paper in Economics No. 16–30.

Karlsson, M., Nilsson T., Pichler S. (2014), The impact of the 1918 Spanish flu epidemic on economic performance in Sweden: An investigation into the consequences of an extraordinary mortality shock. Journal of Health Economics, 36, 1–19.

Kirby T. (2020), Evidence mounts on the disproportionate effect of COVID-19 on ethnic minorities. Lancet Respiratory Medicine. News. May 8.

Krieger N., Wright E., Chen J.T., Waterman P.D., Huntley E.R., Arcaya M. (2020), Cancer stage at diagnosis, historical redlining, and current neighborhood characteristics: Breast, cervical, lung, and colorectal cancer, Massachusetts, 2001-2015. American Journal of Epidemiology, in press.

Krieger N., Van Wye G., Huynh M., Waterman P.D., Maduro G., Li W., Gwynn R.C., Barbot O., Bassett M.T. (2020), Structural racism, historical redlining, and risk of preterm birth in New York City, 20132017. American Journal of Public Health, in press.

Krimmel J. (2018), Persistence of prejudice: Estimating the long term effects of redlining. Mimeo.

Landrine H., Corral I., Lee J.G.L., Efird J.T., Hall M.B., Bess J.J. (2017), Residential segregation and racial cancer disparities: A systematic review. Journal of Racial and Ethnic Health Disparities, 4, 1195–1205.

Lang K., Kahn-Lang Spitzer A. (2020), Race discrimination: An economic perspective. Journal of Economic Perspectives, 34, 68–89.

Lin, M.-J., Liu e.M. (2014), Does in utero exposure to illness matter? The 1918 influenza epidemic in Taiwan as a natural experiment. Journal of Health Economics, 37, 152–163.

Loper J. (2019), Womens position in ancestral societies and female HIV: The long-term effect of matrilineality in sub-Saharan Africa. Mimeo.

McLaren J. (2020), Racial disparity in COVID-19 deaths: Seeking economic roots with census data. NBER Working Paper No. 27407.

Markides K.S:, Coreil J. (1986), The health of Hispanics in the southwestern United States: An epidemiologic paradox. Public Health Reports, 101, 253–265-

Nardone A., Casey J.A., Morello-Frosch R., Mujahid M., Balmes J.R., Thakur N. (2020), Associations between historical residential redlining and current age-adjusted rates of emergency department visits due to asthma across eight cities in California: An ecological study. Lancet Planet Health, 4, e24–31.

Nelson R.K., Winling L., Marciano R., Connolly N., et al. (2020), Mapping inequality: Redlining in New Deal America. In Nelson R.K., Ayers E.L. (eds.), American Panorama: An Atlas of United States History. University of Richmond: Digital Scholarship Lab.

Nier, C.L.III (1999), Perpetuation of segregation: Toward a new historical and legal interpretation of redlining under the Fair Housing Act. John Marshall Law Review, 32, 617–665.

Nunn N. (2008), Slavery, inequality, and economic development in the Americas: An examination of the Engerman-Sokoloff hypothesis. In Helpman E. (ed.), Institutions and Economic Performance. Cambridge: Harvard University Press, 48–180.

Okland H., Mamelund S.-E. (2019), Race and 1918 influenza Pandemic in the United States: A review of the literature. International Journal of Environmental Research and Public Health, 16, 1–18.

OpenSAFELY Collaborative (2020), OpenSAFELY: Factors associated with COVID-19-related hospital death in the linked electronic health records of 17 million adult NHS patients. medRxiv, May 7.

Oppel R.J.Jr., Gebeloff R., Lai K.K.R., Wright W., Smith M. (2020), The fullest look yet at the racial inequity of coronavirus. The New York Times, July 5.

Orsi J.M., Margellos-Anast H., Whitman S. (2010), Black-white health disparities in the United States and Chicago: A 15-year progress analysis. American Journal of Public Health, 100, 349–356.

Platt L., Warwick R. (2020), Are Some Ethnic Groups more Vulnerable to COVID-19 than Others? London: Institute for Fiscal Studies.

Price-Haywood E.G.,Burton J., Fort D., Seoane L. (2020), Hospitalization and mortality among black patients and white patients with Covid-19. The New England Journal of Medicine, 382, 2534–2543.

Reyes C., Husain N., Gutowski C., St. Clair S., Pratt G. (2020), Chicago’s coronavirus disparity: Black Chicagoans are dying at nearly six times the rate of white residents, data show. Chicago Tribune, April 6.

Richardson G., McBride M. (2009), Religion, longevity, and cooperation: The case of the craft guild. Journal of Economic Behavior & Organization, 71, 172–186.

Rothstein R. (2017), The Color of Law: A Forgotten History of How Our Government Segregated America. New York: Liveright.

Schill M.H., Wachter S.M. (1995), The spatial bias of federal housing law and policy: Concentrated poverty in urban America. University of Pennsylvania Law Review, 143, 1285–1342.

Schmitt-Grohe S., Teoh K., Uribe M. (2020), COVID-19: Testing inequality in New York City. NBER Working Paper No. 27019.

Shertzer A., Twinam T., Walsh R. (2016), Race, ethnicity, and discriminatory zoning. American Economic Journal: Applied Economics, 8, 217246.

Singh M., Koran M. (2020), ‘The virus doesn’t discriminate but governments do’: Latinos disproportionately hit by coronavirus. The Guardian, April 18.

Tuttle W.M.Jr. (1970), Race Riot: Chicago in the Summer of 1919. New York: Atheneum.

Voitglander N., Voth H. (2012), Persecution perpetuated: The medieval origins of anti-Semitic violence in Nazi Germany. Quarterly Journal of Economics, 127, 1339–1392.

White C., Nafilyan V. (2020). Coronavirus (COVID-19) related deaths by ethnic group, England and Wales: 2 March 2020 to 10 April 2020. Office for National Statistics.

Wiemers E.E., Abrahams S., AlFakhri M., Hotz V.J., Schoeni R.F., Seltzer J.A. (2020). Disparities in vulnerability to severe complications from Covid-19 in the United States. NBER Working Paper No. 27294.

Yancy C.W. (2020), COVID-19 and African Americans. Journal of the American Medical Association. Opinion. April 15.

Zenou Y., Boccard N. (2000), Racial discrimination and redlining in cities. Journal of Urban Economics, 48, 260–285.

